# Discovery of cardiac imaging biomarkers by training neural network models across diagnostic modalities

**DOI:** 10.1101/2021.02.07.21251025

**Authors:** Shinichi Goto, Andreas A. Werdich, Max Homilius, Jenine E. John, Li-Ming Gan, Calum A. MacRae, Marcelo F. DiCarli, Rahul C. Deo

## Abstract

Machines can be readily trained to automate medical image interpretation, with the primary goal of replicating human capabilities. Here, we propose an alternative role: using machine learning to discover pragmatic imaging-based biomarkers by interpreting one complex imaging modality via a second, more ubiquitous, lower-cost modality. We applied this strategy to train convolutional neural network models to estimate positron emission tomography (PET)-derived myocardial blood flow (MBF) at rest and with hyperemic stress, and their ratio, coronary flow reserve (CFR), using contemporaneous two-dimensional echocardiography videos as inputs. The resulting parameters, echoAI-restMBF, echoAI-stressMBF, and echoAI-CFR modestly approximated the original values. However, using echocardiograms of 5,393 (derivation) and 5,289 (external validation) patients, we show they sharply stratify individuals according to disease comorbidities and combined with baseline demographics, are strong predictors for heart failure hospitalization (C-statistic derivation: 0.79, 95% confidence interval 0.77-0.81; validation: 0.81, 0.79-0.82) and acute coronary syndrome (C-statistic derivation: 0.77, 0.73-0.80; validation: 0.75, 0.73-0.78). Using echocardiograms of 3,926 genotyped individuals, we estimate narrow-sense heritability of 9.2%, 20.4% and 6.5%, respectively for echoAI-restMBF, echoAI-stressMBF, and echoAI-CFR. MBF indices show inverse genetic correlation with impedance-derived body mass indices, such as fat-free body mass (e.g., ρ=−0.43, q=0.05 for echoAI-restMBF) and resolve conflicting historical data regarding body mass index and CFR. In terms of diseases, genetic association with ischemic heart disease is seen most prominently for echoAI-stressMBF (ρ=−0.37, q=2.4×10^−03^). We hypothesize that interpreting one imaging modality through another represents a type of “information bottleneck”, capturing latent features of the original physiologic measurements that have relevance across tissues. Thus, we propose a broader potential role for machine learning algorithms in developing scalable biomarkers that are anchored in known physiology, representative of latent biological factors, and are readily deployable in population health applications.

## Introduction

The automated interpretation of medical images, videos, and signals has been a popular application of machine learning algorithms for decades. Recent examples include detecting breast cancer in mammograms^1^, intracranial hemorrhage in computed tomography scans^2, 3^, and cardiac amyloidosis in electrocardiograms and echocardiograms^4–6^. These and other prevailing use cases primarily view medical data as a depiction of anatomical structures requiring segmentation or a collection of features enriched in specific disease states. However, with its high information content, medical imaging has the potential to provide a window into underlying latent disease processes, which may have prognostic utility or motivate more precise biologically grounded disease definitions.

For example, coronary flow reserve (CFR) is an intriguing physiological parameter defined as the proportional increase in myocardial blood flow during maximal vasodilatory stress^7–10^. It is the ratio of the stress myocardial blood flow (stressMBF) to the rest myocardial blood flow (restMBF), and lower values reflect abnormalities in the coronary epicardial vessels and microvasculature. These values have prognostic utility in predicting cardiovascular outcomes^11–14^ and are abnormal in several cardiac and systemic disease states, including dilated cardiomyopathy^15^, heart failure with preserved ejection fraction^16^, osteoporosis^17^, systemic lupus erythematosus^18^, rheumatoid arthritis^18^, and chronic obstructive pulmonary disease^19^. Coronary blood flow indices are unfortunately challenging to measure without invasive coronary angiography or indirectly through magnetic resonance imaging or positron emission tomography (PET), all of which have high associated costs.

CFR has a clear physiological basis, and its association with non-cardiac diseases implies that it reflects more global underlying biologic processes that may be captured by other potentially lower cost modalities. Beyond the pragmatism of developing a more scalable version of these original measurements, building a representation of one modality with another may provide a cleaner readout of the underlying signal free of technical artifacts, reminiscent of the concept of an information bottleneck in machine learning^20–22^. In this study, we exploited this strategy for biomarker discovery by training convolutional neural networks to estimate rest and stress MBF and CFR using only resting echocardiogram videos as inputs. We deployed the resulting models on thousands of echocardiograms, enabling development and external validation of prognostic models, and investigating their genetic basis.

## Results

### Convolutional neural network models using resting echocardiograms modestly estimate indices of PET-determined myocardial blood flow

An overview of our strategy is shown in **Figure 1**. We selected a total of 6,147 echocardiogram studies that occurred within one year of 3,448 corresponding PET studies and trained three-dimensional convolutional neural networks using resting apical 4-chamber (A4c) videos to estimate: 1) myocardial blood flow (MBF) at rest; (2) after vasodilatory stress; and 3) coronary flow reserve (**Figures S1 and S2**). We refer to the resulting parameter estimates as echoAI-restMBF, echoAI-stressMBF, and echoAI-CFR. The output of echo-AI models only had a modest correlation with the corresponding PET measurement: the Spearman’s rank correlation coefficients were 0.35, 0.42, and 0.29 for rest MBF, stress MBF, and CFR, respectively. The corresponding median absolute errors were 0.16 ml/g/min, 0.5 ml/g/min, and 0.4 (**Figure S3**).

**Figure 1.**
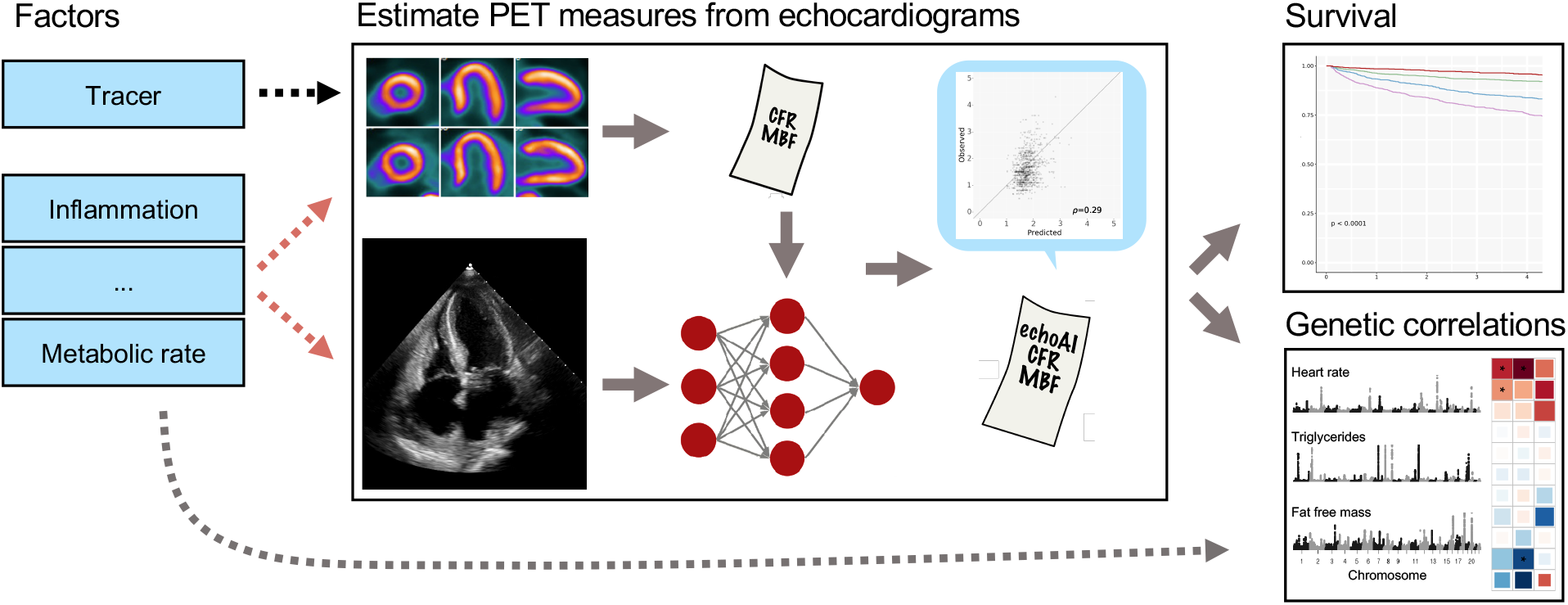
Graphical overview of the study. Myocardial blood flow (MBF), as measured by PET, is a powerful predictor of cardiovascular outcomes, but has restricted availability due to equipment cost, tracer availability, technical expertise required, and radiation risk. In contrast, two-dimensional B-mode echocardiography measured at rest is a widely available, scalable technology. We hypothesized that many biological factors that drive abnormalities in MBF (red dotted arrows) would also be reflected within echocardiograms whereas technical factors unique to PET (black dotted arrow) would not. We trained three-dimensional convolutional neural network-derived models to estimate PET-derived indices of MBF, including CFR, using contemporaneously measured echocardiogram videos as inputs. We deployed the resulting models on tens of thousands of echocardiogram videos and found that echoAI derived MBF parameters and CFR sharply stratify patients by comorbidities, predict cardiovascular outcomes, and have genetic correlation with a breadth of clinical traits, shedding light on underlying biological mechanisms.

### Echo-AI stress MBF and echo-AI CFR stratify patients by age and cardiac and metabolic disease status

Although the agreement with the original PET-derived estimates was modest, we nonetheless explored the utility of our echoAI-derived measures, which are scalable, and thus can be deployed on a large number of echocardiograms. We selected all patients at Brigham and Women’s Hospital (BWH) who had at least one echocardiogram within a 6-month period (June 2015-November 2015). We used the earliest echocardiogram for each patient, resulting in 5,393 echocardiograms, and we deployed all 3 models on all available A4c videos. Analysis of the baseline patient characteristics stratified by echoAI-restMBF, echoAI-stressMBF, and echoAI-CFR values (**Tables 1-3**) revealed a sharp separation of comorbidity status by quartiles of echoAI-stressMBF and echoAI-CFR. Patients with lower echoAI-stressMBF and echoAI-CFR were older, had worse cardiac disease status, including lower ejection fraction and higher proportions of heart failure, atrial fibrillation, and coronary artery disease (CAD; all p<0.001). Furthermore, strata defined by low echoAI-stressMBF and echoAI-CFR reflected key facets of metabolic disorders associated with microvascular dysfunction, including a greater proportion of patients with type 2 diabetes mellitus, hyperlipidemia, and renal dysfunction^12^ (**Tables 2 and 3**).

**Table 1.** Baseline demographic for EchoAI-restMBF quartiles (BWH cohort)

|  | Missing n (%) | Quartile 1 | Quartile 2 | Quartile 3 | Quartile 4 | p |
| --- | --- | --- | --- | --- | --- | --- |
| Number patients |  | 1,349 | 1,348 | 1,348 | 1,348 |  |
| **AI model** |  |  |  |  |  |  |
| Echo-AI Rest MBF, A.U. ± SD | 0 (0) | 0.7 ± 0.1 | 0.8 ± 0.0 | 0.9 ± 0.0 | 1.0 ± 0.1 | <0.001 |
| Echo-AI Stress MBF, A.U. ± SD | 0 (0) | 0.9 ± 0.4 | 1.2 ± 0.4 | 1.4 ± 0.4 | 1.6 ± 0.4 | <0.001 |
| Echo-AI CFR, A.U. ± SD | 0 (0) | 1.9 ± 0.4 | 2.0 ± 0.4 | 2.0 ± 0.4 | 2.0 ± 0.4 | <0.001 |
| **Demographics** |  |  |  |  |  |  |
| Age, years ± SD | 11 (0.2) | 62.3 ± 16.3 | 59.9 ± 16.5 | 60.7 ± 16.0 | 58.1 ± 16.2 | <0.001 |
| Gender, Female (%) | 11 (0.2) | 405 (30.1) | 679 (50.5) | 809 (60.1) | 963 (71.5) | <0.001 |
| Smoking | 0 (0) |  |  |  |  | <0.001 |
| Never, n (%) |  | 624 (46.3) | 681 (50.5) | 711 (52.7) | 745 (55.3) |  |
| Current, n (%) |  | 75 (5.6) | 89 (6.6) | 82 (6.1) | 77 (5.7) |  |
| Passive, n (%) |  | 1 (0.1) | 0 (0.0) | 3 (0.2) | 6 (0.4) |  |
| Quit, n (%) |  | 516 (38.3) | 441 (32.7) | 452 (33.5) | 418 (31.0) |  |
| Unknown, n (%) |  | 133 (9.9) | 137 (10.2) | 100 (7.4) | 102 (7.6) |  |
| Race | 0 (0) |  |  |  |  | 0.051 |
| White, n (%) |  | 1,099 (81.5) | 1,086 (80.6) | 1,098 (81.5) | 1,063 (78.9) |  |
| Black, n (%) |  | 112 (8.3) | 91 (6.8) | 93 (6.9) | 110 (8.2) |  |
| Hispanic, n (%) |  | 44 (3.3) | 31 (2.3) | 33 (2.4) | 44 (3.3) |  |
| Asian, n (%) |  | 21 (1.6) | 40 (3.0) | 29 (2.2) | 38 (2.8) |  |
| Others, n (%) |  | 27 (2.0) | 40 (3.0) | 49 (3.6) | 45 (3.3) |  |
| Unknown, n (%) |  | 46 (3.4) | 60 (4.5) | 46 (3.4) | 48 (3.6) |  |
| HR, bpm ± SD | 384 (7.1) | 74.3 ± 13.9 | 73.4 ± 13.0 | 75.1 ± 12.0 | 78.6 ± 11.9 | <0.001 |
| SBP, mmHg ± SD | 361 (6.7) | 126.8 ± 16.8 | 128.4 ± 14.8 | 128.9 ± 15.3 | 129.1 ± 15.3 | 0.001 |
| DBP, mmHg ± SD | 361 (6.7) | 73.8 ± 11.6 | 73.6 ± 9.9 | 73.7 ± 9.9 | 73.5 ± 9.6 | 0.957 |
| BMI, kg/m<sup>2</sup> ± SD | 607 (11.3) | 29.0 ± 7.6 | 28.6 ± 7.3 | 28.3 ± 6.9 | 27.6 ± 8.7 | <0.001 |
| eGFR, ml/min/1.73 m<sup>2</sup> ± SD | 561 (10.4) | 63.4 ± 27.7 | 79.3 ± 28.7 | 80.2 ± 28.1 | 87.6 ± 29.3 | <0.001 |
| EF, % ± SD | 411 (7.6) | 52.5 ± 14.0 | 59.5 ± 8.1 | 61.4 ± 6.7 | 62.6 ± 5.7 | <0.001 |
| **Disease history** |  |  |  |  |  |  |
| Heart failure, n (%) | 3 (0.0) | 456 (33.8) | 234 (17.4) | 143 (10.6) | 133 (9.9) | <0.001 |
| Type 2 DM, n (%) | 3 (0.0) | 271 (20.1) | 214 (15.9) | 220 (16.3) | 191 (14.2) | <0.001 |
| Hypertension, n (%) | 3 (0.0) | 733 (54.4) | 714 (53.0) | 733 (54.4) | 668 (49.6) | 0.041 |
| Atrial fibrillation, n (%) | 3 (0.0) | 483 (35.8) | 215 (16.0) | 127 (9.4) | 90 (6.7) | <0.001 |
| Hyperlipidemia, n (%) | 3 (0.0) | 501 (37.2) | 486 (36.1) | 497 (36.9) | 412 (30.6) | 0.001 |
| CAD, n (%) | 3 (0.0) | 369 (27.4) | 270 (20.0) | 191 (14.2) | 138 (10.2) | <0.001 |
| Stroke, n (%) | 3 (0.0) | 118 (8.8) | 91 (6.8) | 78 (5.8) | 69 (5.1) | 0.001 |
| PAD, n (%) | 3 (0.0) | 139 (10.3) | 125 (9.3) | 119 (8.8) | 99 (7.3) | 0.057 |
| COPD, n (%) | 3 (0.0) | 76 (5.6) | 88 (6.5) | 100 (7.4) | 81 (6.0) | 0.260 |
| Aortic stenosis | 3 (0.0) | 82 (6.1) | 66 (4.9) | 80 (5.9) | 57 (4.2) | 0.101 |
| Aortic regurgitation | 3 (0.0) | 22 (1.6) | 31 (2.3) | 28 (2.1) | 20 (1.5) | 0.365 |
| Mitral stenosis | 3 (0.0) | 13 (1.0) | 6 (0.4) | 5 (0.4) | 4 (0.3) | 0.067 |
| Mitral regurgitation | 3 (0.0) | 101 (7.5) | 67 (5.0) | 64 (4.7) | 48 (3.6) | <0.001 |
| **Medication** |  |  |  |  |  |  |
| ASA, n (%) | 3 (0.0) | 679 (50.4) | 628 (46.6) | 578 (42.9) | 505 (37.5) | <0.001 |
| Statin | 3 (0.0) | 702 (62.1) | 586 (43.5) | 583 (43.2) | 448 (33.3) | <0.001 |
| ACEi, n (%) | 3 (0.0) | 442 (32.8) | 354 (26.3) | 330 (24.5) | 295 (21.9) | <0.001 |
| ARB, n (%) | 3 (0.0) | 220 (16.3) | 189 (14.0) | 204 (15.1) | 174 (12.9) | 0.075 |
| Beta blocker, n (%) | 3 (0.0) | 792 (58.8) | 616 (45.7) | 562 (41.7) | 432 (32.1) | <0.001 |
| Thiazide, n (%) | 3 (0.0) | 180 (13.4) | 154 (11.4) | 168 (12.5) | 161 (12.0) | 0.474 |
| Loop diuretic, n (%) | 3 (0.0) | 504 (37.4) | 272 (20.2) | 204 (15.1) | 162 (12.0) | <0.001 |
| MRA, n (%) | 3 (0.0) | 160 (11.9) | 86 (6.4) | 54 (4.0) | 48 (3.6) | <0.001 |

Abbreviations. BWH: Brigham and Women's Hospital, CFR: coronary flow reserve, MBF: myocardial blood flow, A.U.: arbitrary unit, HR: heart rate, SBP: systolic blood pressure, DBP: diastolic blood pressure, BMI: body mass index, eGFR: estimated glomerular filtration rate, EF: ejection fraction, LV: left ventricle, DM: diabetes mellitus, CAD: coronary heart disease, PAD: peripheral artery disease, COPD: chronic obstructive pulmonary disease, ASA: acetyl-salicylic acid, ACEi: angiotensin-converting enzyme inhibitor, ARB: angiotensin II receptor blocker, MRA: mineralocorticoid receptor antagonist

**Table 2.** Baseline demographic for EchoAI-stressMBF quartiles (BWH cohort)

|  | Missing n (%) | Quartile 1 | Quartile 2 | Quartile 3 | Quartile 4 | p |
| --- | --- | --- | --- | --- | --- | --- |
| Number patients |  | 1,349 | 1,348 | 1,348 | 1,348 |  |
| **AI model** |  |  |  |  |  |  |
| Echo-AI Rest MBF, A.U. ± SD | 0 (0) | 0.7 ± 0.1 | 0.8 ± 0.1 | 0.9 ± 0.1 | 0.9 ± 0.1 | <0.001 |
| Echo-AI Stress MBF, A.U. ± SD | 0 (0) | 0.7 ± 0.2 | 1.1 ± 0.1 | 1.4 ± 0.1 | 1.9 ± 0.2 | <0.001 |
| Echo-AI CFR, A.U. ± SD | 0 (0) | 1.7 ± 0.2 | 1.9 ± 0.3 | 2.0 ± 0.3 | 2.3 ± 0.4 | <0.001 |
| **Demographics** |  |  |  |  |  |  |
| Age, years ± SD | 11 (0.2) | 68.3 ± 13.8 | 63.0 ± 15.2 | 58.9 ± 15.0 | 50.8 ± 15.9 | <0.001 |
| Gender, Female (%) | 11 (0.2) | 419 (31.2) | 620 (46.1) | 788 (58.5) | 1029 (76.6) | <0.001 |
| Smoking | 0 (0) |  |  |  |  | <0.001 |
| Never, n (%) |  | 575 (42.6) | 663 (49.2) | 705 (52.3) | 818 (60.7) |  |
| Current, n (%) |  | 68 (5.0) | 83 (6.2) | 88 (6.5) | 84 (6.2) |  |
| Passive, n (%) |  | 2(0.1) | 1 (0.1) | 3 (0.2) | 4 (0.3) |  |
| Quit, n (%) |  | 579 (42.9) | 484 (35.9) | 444 (32.9) | 320 (23.7) |  |
| Unknown, n (%) |  | 125 (9.3) | 117 (8.7) | 108 (8.0) | 122 (9.1) |  |
| Race | 0 (0) |  |  |  |  | 0.008 |
| White, n (%) |  | 1,100 (81.5) | 1,091 (80.9) | 1,102 (81.8) | 1,053 (78.1) |  |
| Black, n (%) |  | 108 (8.0) | 107 (7.9) | 91 (6.8) | 100 (7.4) |  |
| Hispanic, n (%) |  | 26 (1.9) | 41 (3.0) | 42 (3.1) | 43 (3.2) |  |
| Asian, n (%) |  | 24 (1.8) | 28 (2.1) | 24 (1.8) | 52 (3.9) |  |
| Others, n (%) |  | 35 (2.6) | 34 (2.5) | 40 (3.0) | 52 (3.9) |  |
| Unknown, n (%) |  | 56 (4.2) | 47 (3.5) | 49 (3.6) | 48 (3.6) |  |
| HR, bpm ± SD | 384 (7.1) | 75.4 ± 12.9 | 72.9 ± 13.3 | 73.1 ± 13.0 | 76.1 ± 12.1 | <0.001 |
| SBP, mmHg ± SD | 361 (6.7) | 128.8 ± 17.1 | 130.2 ± 15.5 | 128.8 ± 14.5 | 125.4 ± 14.8 | <0.001 |
| DBP, mmHg ± SD | 361 (6.7) | 72.3 ± 11.6 | 73.8 ± 10.2 | 74.5 ± 9.7 | 73.9 ± 9.4 | <0.001 |
| BMI, kg/m<sup>2</sup> ± SD | 607 (11.3) | 29.6 ± 8.3 | 29.3 ± 8.8 | 28.3 ± 6.3 | 26.5 ± 6.6 | <0.001 |
| eGFR, ml/min/1.73 m<sup>2</sup> ± SD | 561 (10.4) | 63.3 ± 25.9 | 74.5 ± 26.6 | 83.3 ± 27.0 | 95.7 ± 27.1 | <0.001 |
| EF, % ± SD | 411 (7.6) | 51.8 ± 14.4 | 60.0 ± 7.8 | 61.7 ± 5.7 | 62.5 ± 5.2 | <0.001 |
| **Disease history** |  |  |  |  |  |  |
| Heart failure, n (%) | 3 (0.0) | 542 (40.2) | 226 (16.8) | 142 (10.5) | 56 (4.2) | <0.001 |
| Type 2 DM, n (%) | 3 (0.0) | 345 (25.6) | 263 (19.5) | 181 (13.4) | 107 (7.9) | <0.001 |
| Hypertension, n (%) | 3 (0.0) | 875 (64.9) | 822 (61.1) | 681 (50.5) | 470 (34.9) | <0.001 |
| Atrial fibrillation, n (%) | 3 (0.0) | 539 (40.0) | 236 (17.5) | 84 (6.2) | 56 (4.2) | <0.001 |
| Hyperlipidemia, n (%) | 3 (0.0) | 591 (43.8) | 551 (40.9) | 447 (33.2) | 307 (22.8) | <0.001 |
| CAD, n (%) | 3 (0.0) | 493 (36.6) | 267 (19.8) | 156 (11.6) | 52 (3.9) | <0.001 |
| Stroke, n (%) | 3 (0.0) | 141 (10.5) | 88 (6.5) | 84 (6.2) | 43 (3.2) | <0.001 |
| PAD, n (%) | 3 (0.0) | 168 (12.5) | 139 (10.3) | 118 (8.8) | 57 (4.2) | <0.001 |
| COPD, n (%) | 3 (0.0) | 110 (8.2) | 99 (7.4) | 80 (5.9) | 56 (4.2) | <0.001 |
| Aortic stenosis | 3 (0.0) | 129 (9.6) | 103 (7.7) | 37 (2.7) | 16 (1.2) | <0.001 |
| Aortic regurgitation | 3 (0.0) | 31 (2.3) | 30 (2.2) | 27 (2.0) | 13 (1.0) | 0.038 |
| Mitral stenosis | 3 (0.0) | 16 (1.2) | 8 (0.6) | 4 (0.3) | 0 (0.0) | <0.001 |
| Mitral regurgitation | 3 (0.0) | 133 (9.9) | 85 (6.3) | 39 (2.9) | 23 (1.7) | <0.001 |
| **Medication** |  |  |  |  |  |  |
| ASA, n (%) | 3 (0.0) | 819 (60.8) | 693 (51.5) | 543 (40.3) | 335 (24.9) | <0.001 |
| Statin | 3 (0.0) | 845 (62.7) | 682 (50.7) | 514 (38.1) | 278 (20.6) | <0.001 |
| ACEi, n (%) | 3 (0.0) | 487 (36.1) | 387 (28.8) | 323 (24.0) | 224 (16.6) | <0.001 |
| ARB, n (%) | 3 (0.0) | 275 (20.4) | 228 (16.9) | 182 (13.5) | 102 (7.6) | <0.001 |
| Beta blocker, n (%) | 3 (0.0) | 953 (70.7) | 728 (54.1) | 472 (35.0) | 249 (18.5) | <0.001 |
| Thiazide, n (%) | 3 (0.0) | 198 (14.7) | 179 (13.3) | 173 (12.8) | 113 (8.4) | <0.001 |
| Loop diuretic, n (%) | 3 (0.0) | 636 (47.2) | 256 (19.0) | 164 (12.2) | 86 (6.4) | <0.001 |
| MRA, n (%) | 3 (0.0) | 192 (14.2) | 80 (5.9) | 40 (3.0) | 36 (2.7) | <0.001 |

Abbreviations. BWH: Brigham and Women's Hospital, CFR: coronary flow reserve, MBF: myocardial blood flow, A.U.: arbitrary unit, HR: heart rate, SBP: systolic blood pressure, DBP: diastolic blood pressure, BMI: body mass index, eGFR: estimated glomerular filtration rate, EF: ejection fraction, LV: left ventricle, DM: diabetes mellitus, CAD: coronary heart disease, PAD: peripheral artery disease, COPD: chronic obstructive pulmonary disease, ASA: acetyl-salicylic acid, ACEi: angiotensin-converting enzyme inhibitor, ARB: angiotensin II receptor blocker, MRA: mineralocorticoid receptor antagonist

**Table 3.**
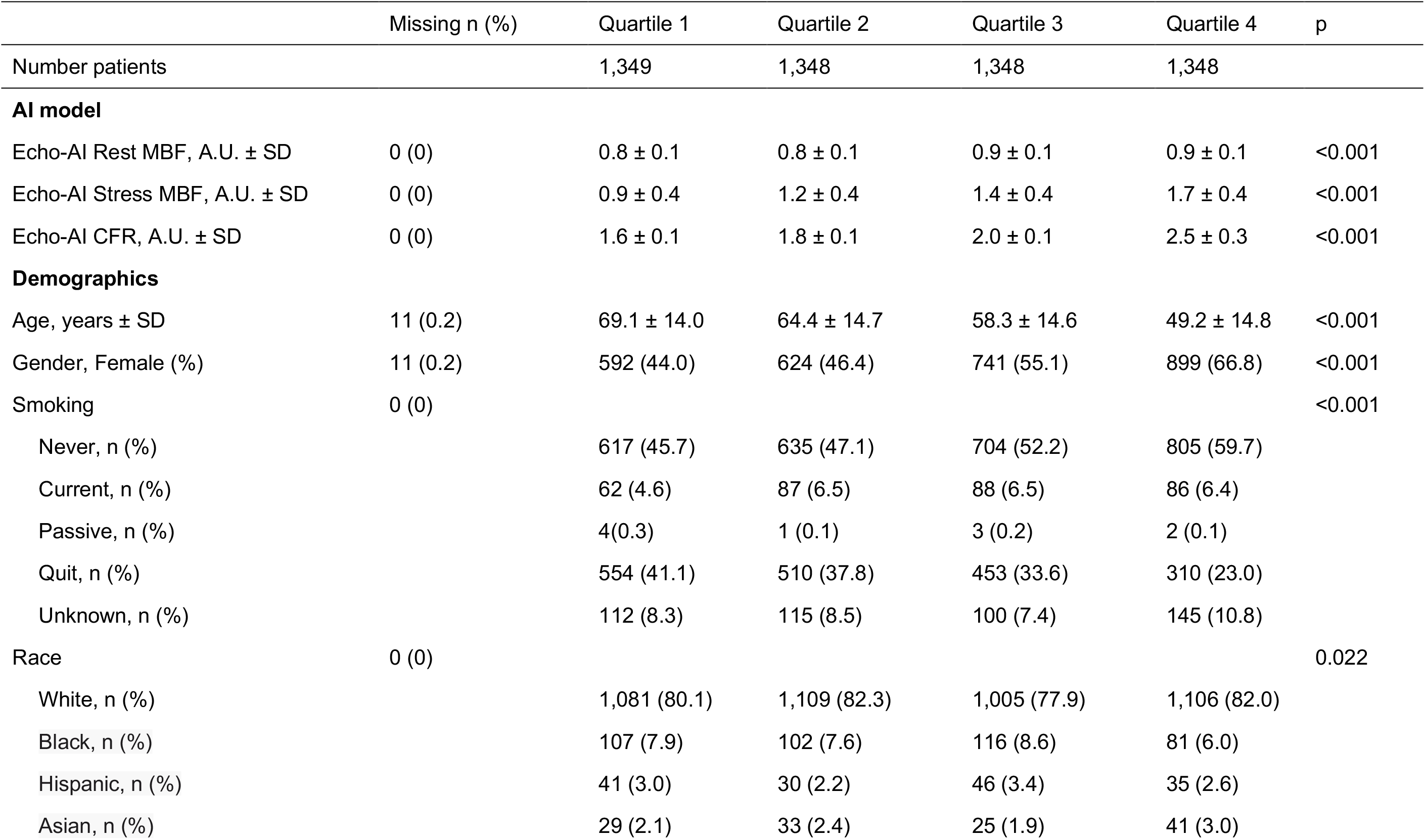
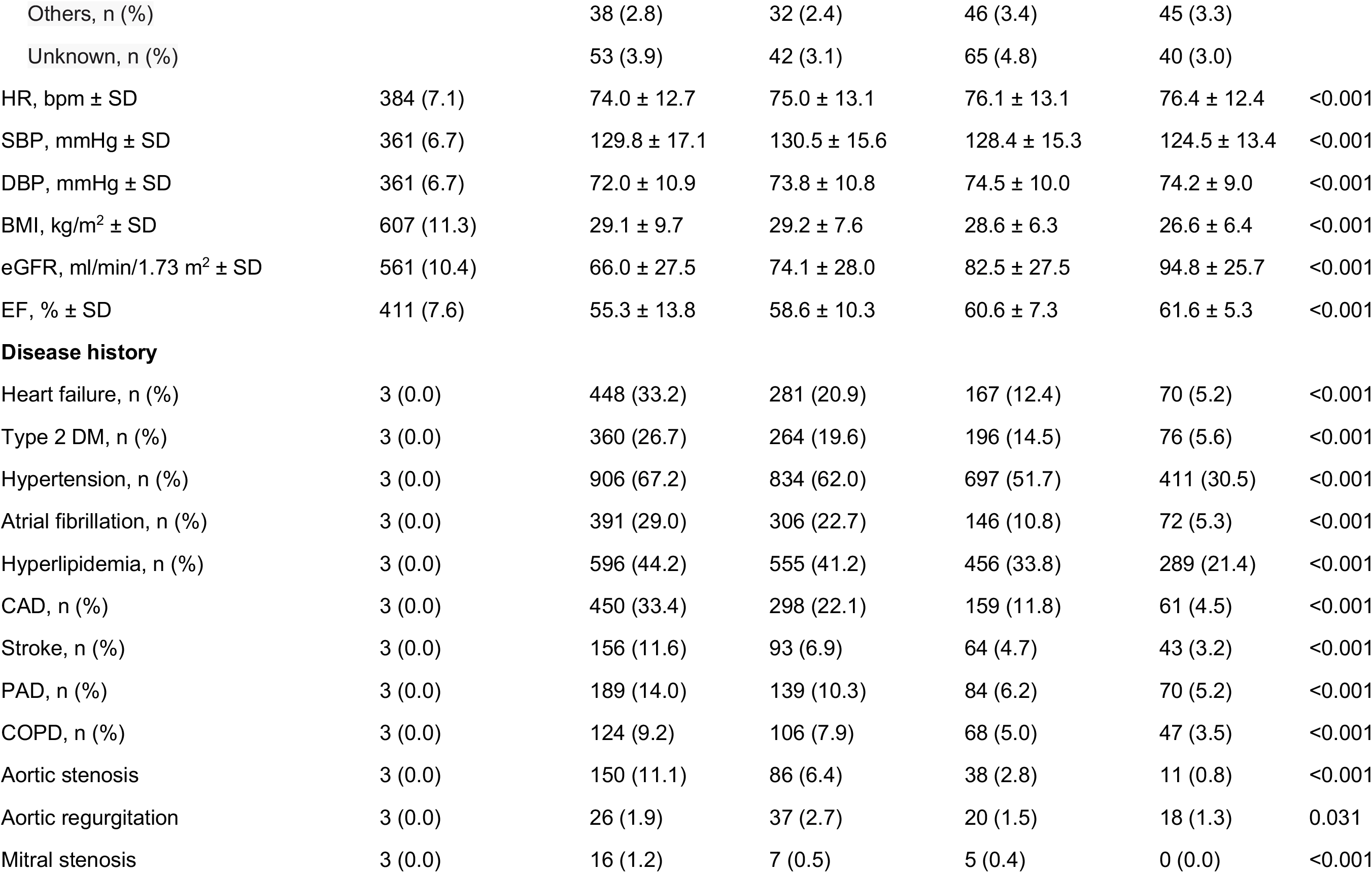
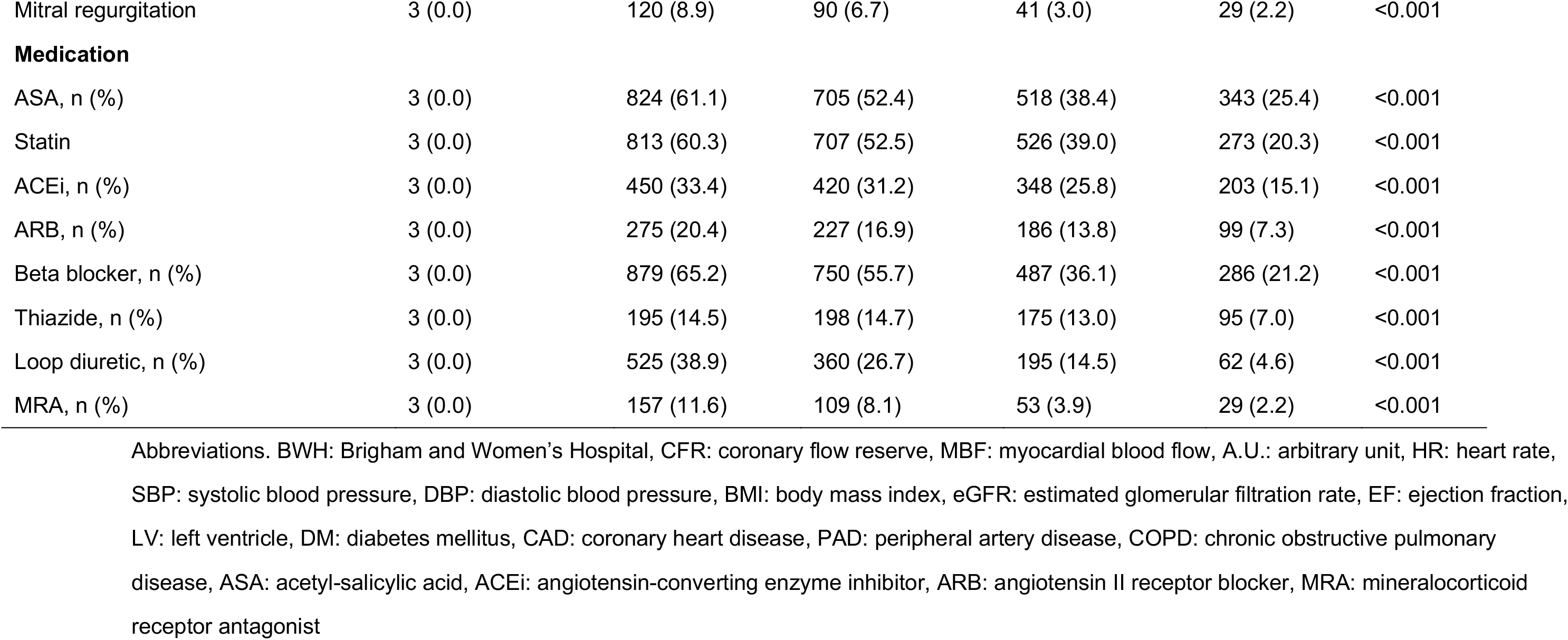
Baseline demographic for EchoAI-CFR quartiles (BWH cohort)

The echoAI-restMBF values similarly separated patients according to these baseline attributes but with less sharp differences in the prevalence of comorbidities and indices of diminished cardiac and renal performance (**Table 1**). We further validated these results using an external cohort consisting of 5,289 patients from Massachusetts General Hospital (MGH) who had at least one echocardiogram within Feb 2016-Nov 2016 and observed a similar stratification of comorbidities with echoAI measures (**Tables S1-S3**).

### EchoAI-CFR and echoAI-stressMBF predict cardiovascular outcomes, independent of known risk markers

To assess the ability of echoAI measures to predict adverse cardiovascular outcomes, we performed a time-to-event analysis on the same 5,393 patients from BWH (**Figure S4, Tables S4-S9**) for the outcomes of heart failure (HF) hospitalization and acute coronary syndrome (ACS), adjudicated by a machine learning algorithm as described previously^19^. The median follow-up length for this cohort was 4.1 (interquartile range (IQR): 1.7-4.4) years for heart failure and 4.2 (2.1-4.4) years for ACS. In univariate analysis, the echoAI measures predicted HF hospitalization with AUC 0.60 (95% CI 0.57-0.63), 0.70 (0.68-0.73) and 0.71 (0.69-0.73) and ACS with AUC 0.58 (0.54-0.62), 0.67 (0.63-0.72), and 0.66 (0.63-0.70) for echoAI-restMBF, echoAI-stressMBF and echo-AI-CFR respectively (**Figure 2**, **Figure S5 and S6**). Importantly, the predictive accuracy of echoAI-stressMBF and echoAI-CFR was higher than that of known HF predictors. For example, for HF hospitalization, the AUC for age alone was 0.67, and that of HF history alone was 0.66. For ACS, these echoAI measures’ predictive accuracy was similar to that of known strong predictors such as age and CAD history. We further performed a multivariable Cox proportional hazard model analysis with 9 parameters (the 3 echoAI parameters and 6 baseline characteristics including sex, age, ejection fraction, heart rate, history of CAD, and history of heart failure), which revealed that echoAI-stressMBF and echoAI-CFR were associated with HF hospitalization (hazard ratio per standard deviation change: echoAI-stressMBF=0.78, 95% CI 0.68-0.90, echoAI-CFR=0.71 95% CI 0.63-0.81) and echoAI-stressMBF was associated with ACS (HR: echoAI-stressMBF=0.73, 95% CI 0.56-0.95), independent of other covariates (**Figure 3**). The ROC analysis of the output of this 9-parameter Cox proportional hazard model predicted HF hospitalization (C-statistic 0.79 95%CI 0.77-0.88) and ACS (C-statistic 0.77, 0.73-0.80) with high discriminative accuracy (**Figure S5B and S6B)** and excellent calibration (**Figure S5C and S6C)**.

**Figure 2.**
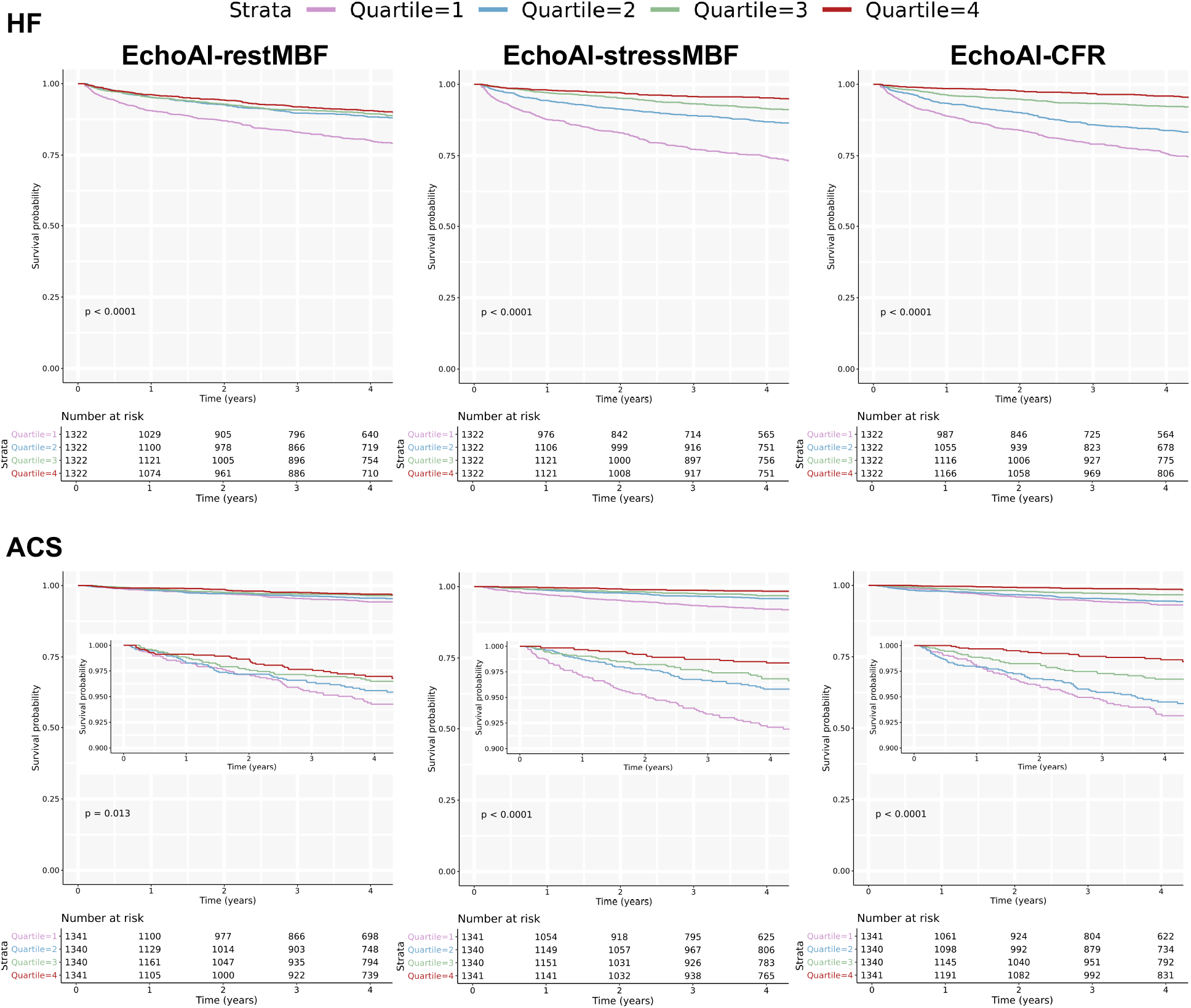
Cardiovascular event free survival for echo-AI measurements quartiles. Kaplan-Meier plots for HF hospitalization and ACS free survival for echoAI measurement quartiles.

**Figure 3.**
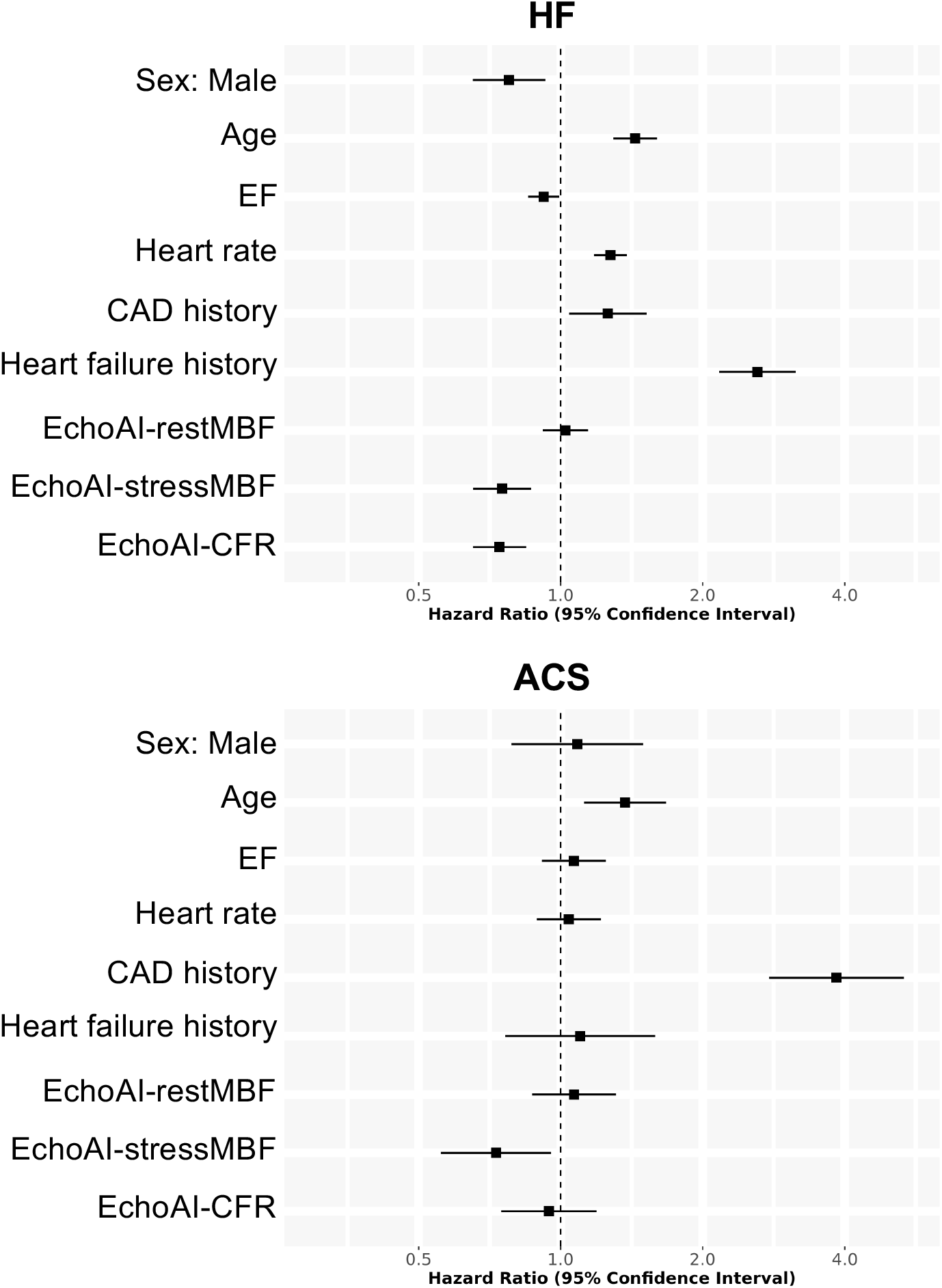
Hazard ratios of echoAI measurements and various patient characteristics for the multivariable Cox proportional hazard models. Forest plot showing the hazard ratio and 95% confidence intervals for each parameter in the Cox proportional hazard models for heart failure hospitalization and ACS. Hazard ratios are plotted as per standard deviation. Abbreviations, ACS: acute coronary syndrome, EF: ejection fraction, CAD: coronary artery disease, MBF: myocardial blood flow, CFR: coronary flow reserve,

We validated this result using the MGH external validation cohort (**Figure S7, Tables S10-S15**). This cohort’s median follow-up time was 3.3 (IQR: 1.4-3.6) years and 3.3 (0.9-3.6) years for heart failure hospitalization and ACS, respectively. As with the BWH cohort, echoAI-stressMBF and echoAI-CFR accurately stratified patients by risk of HF hospitalization and ACS in this cohort (**Figure S8**). Multivariable Cox proportional hazard models, trained *de novo* on MGH data, yielded results consistent with findings from the BWH cohort: echoAI-stressMBF and echoAI-CFR both associated with HF hospitalization (HR per standard deviation change: 0.87 95%CI 0.77-0.98 and 0.76 95%CI 0.68-0.84, respectively) and echoAI-stressMBF associated with ACS (HR per standard deviation change: 0.82 95%CI 0.68-0.99) independent of other baseline characteristics (**Figures S9**). We further tested the predictive accuracy of the individual echoAI measurements (along with the 6 baseline features) and the combined 9-parameter model trained on BWH data. In the univariate analysis, predictive accuracy assessed by C-statistics was similar for all 9 parameters (**Figures S10A and S11A**). The 9-parameter model performed similarly to the derivation BWH cohort for heart failure hospitalization (**Figure S10B**, C-statistic 0.81 95% CI 0.79-0.82) and ACS (**Figure S11B**, C-statistic 0.75 95% CI 0.73-0.78). Furthermore, the models had excellent calibration on the MGH cohort without any adjustment (**Figures S10C and S11C**)

### Genetic correlation of echoAI flow parameters with conventional traits

Given that our echoAI models have a modest input requirement - an echocardiogram with an apical 4-chamber view – they can enable investigations that have been historically challenging because of limited data availability. For example, there have been no systematic large-scale studies to understand the genetic determinants of myocardial blood flow, primarily because the imaging modalities used are invasive or require specialized expertise to acquire. We deployed the echoAI-restMBF, echoAI-stressMBF, and echoAI-CFR on echocardiograms of 3,926 participants from the Mass General Brigham Biobank, a repository of biomolecular samples combined with clinical data. We used the resulting model outputs as continuous phenotypes and performed a genomewide association study to identify genetic determinants. Although the top SNP for all traits fell below conventional genomewide significance thresholds, we used summary statistics to estimate narrow-sense trait heritability and explore genetic correlations with a diversity of phenotypes, some of which have previously been associated with MBF or CFR. We estimated heritability for echoAI-restMBF, echoAI-stressMBF and echoAI-CFR as 9.2%, 20.4%, and 6.5% respectively and found the MBF traits to be modestly correlated with one another (restMBF/stressMBF ρ=0.26).

We next focused on understanding whether the genetic determinants of myocardial blood flow indices are shared with any other trait, using summary statistics from the UK Biobank as a source for comparison. Genetic correlations are useful for parameters where phenotypic correlations with the underlying trait of interest may not be feasible, either because the trait has not been measured in the population or it has been modified by treatment. The strongest genetic correlations were seen for echoAI-stressMBF, which also had the highest estimated heritability. echoAI-stressMBF had an inverse genetic correlation (**Figure 4**) with several traits reflective of impedance-estimated body mass, including whole-body fat-free mass (ρ=−0.32, q=5.1×10^−05^) and basal metabolic rate (ρ=−0.33, q=5.1 ×10^−05^). echoAI-stressMBF also had negative correlation with ischemic heart disease (ρ=−0.37, q=2.4 ×10^−03^) and C-reactive protein (ρ=−0.21, q=0.09). Finally, echoAI-stressMBF had a weak positive genetic correlation with heart rate (ρ=0.16, q=0.10).

**Figure 4.**
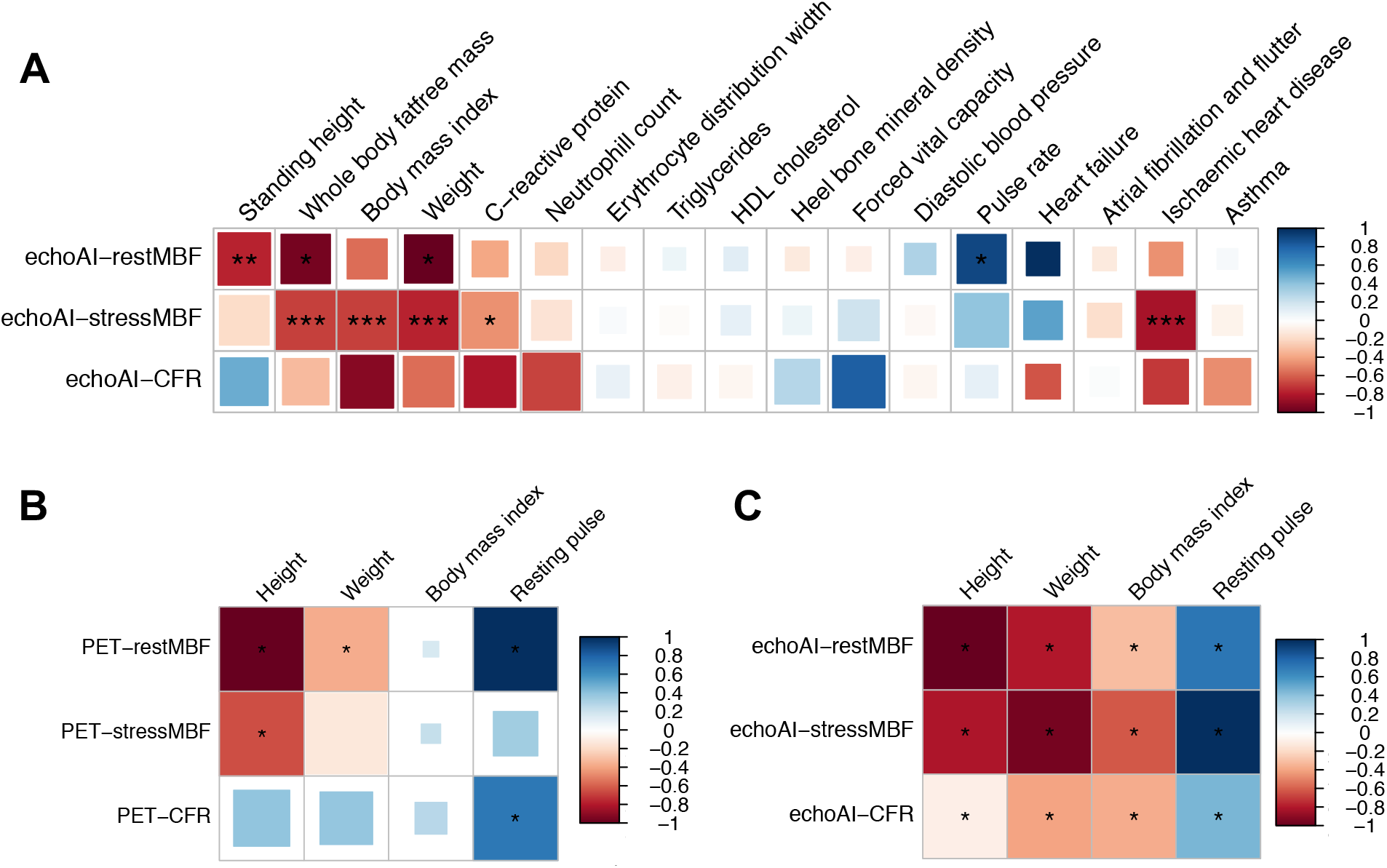
Genetic and phenotypic correlation of echoAI-CFR, echoAI-restMBF and echoAI-stressMBF with traits previously associated with indices of coronary blood flow. (A) Heatmap representing the genetic correlation coefficient values between echo-AI GWAS summary statistics and multiple traits in UK Biobank. Significant genetic correlations at FDR thresholds 0.01, 0.05, 0.1 are indicated by ***, **, *. (B) Correlation of PET measures with anthropomorphic and physiologic traits. (C) Correlation of echo-AI estimates with anthropomorphic and physiologic traits. Significant phenotypic correlations with Bonferroni-corrected p-values < 0.05 are indicated by *.

We observed similar genetic correlations with echoAI-restMBF, including an even stronger correlation with whole body fat-free mass (ρ=−0.43, q=0.05) and heart rate (ρ=−0.38, q=0.06). As expected, echoAI-restMBF had a weaker correlation with ischemic heart disease (ρ=−0.22, q=0.26). echoAI-restMBF also had a surprising inverse genetic correlation with standing height (ρ=−0.35, q=0.04).

Compared with the MBF indices, echoAI-CFR had a weaker correlation with whole body fat-free mass (ρ=−0.15, q=0.27) but interestingly a stronger negative correlation with body mass index (ρ=−0.41, q=0.19) and waist circumference (ρ=−0.37, q=0.20). echoAI-CFR had a negative but non-significant correlation with ischemic heart disease (ρ=−0.33, q=0.30), a negative correlation with neutrophil count (ρ=−0.31, q=0.14), a negative correlation with C-reactive protein (ρ=−0.37, q=0.19), and positive correlation with forced vital capacity (ρ=0.34, q=0.19). Larger sample sizes will be needed to estimate these parameters more precisely.

We confirmed several anthropomorphic and physiologic associations at the phenotypic level, using both PET-derived and echoAI indices (**Figure 4**). For height, Spearman correlations were −0.30 −0.26 and 0.0 for PET-restMBF, PET-stressMBF, and PET-CFR and for weight they were −0.14, −0.08, and 0.04, respectively. BMI had no apparent correlation with any of the PET blood flow indices. Resting heart rate was significantly correlated only with PET-restMBF (ρ=0.20) and PET-CFR (ρ=0.13). For the echoAI measures, the Spearman correlations with height were −0.30, −0.25, and −0.13 for echoAI-restMBF, echoAI-stressMBF, and echoAI-CFR, respectively. For weight, Spearman correlations were −0.24, −0.29, and −0.14; for BMI −0.12, −0.20, and −0.13; and for heart rate, −0.15 −0.22, and −0.08, respectively (**Figure 4**).

## Discussion

The primary, altogether unexpected result of this work is the fact that relatively modest estimators of complex physiologic parameters can be 1) strongly associated with specific comorbidities associated with abnormal coronary flow; 2) powerful predictors of cardiovascular outcomes independent of known risk markers; and 3) have a defined genetic basis that, through genetic correlation analyses, can shed light on latent biological processes that underlie these metrics.

By mapping PET-derived coronary blood flow indices to information derived from a single view of a 2D echocardiogram, we markedly increased the scale of downstream analyses, including longitudinal time-to-event analyses and genetic association analysis. From a workflow standpoint, this mapping of the output of a costly imaging modality to a more readily available one (i.e., echocardiography without Doppler) should also enable broader clinical deployment.

The rationale for this work is a belief in common biological factors or pathways underlying a broader set of disease manifestations. In the case of CFR, the strong association of a coronary blood flow parameter with a diverse group of measurements and disease states, including indices of myocardial function^23^, systemic inflammatory markers^19^, pulmonary disease^19^, and bone disease ^17^ suggest the existence of common latent processes. A parsimonious explanation is that vascular beds across a wide range of tissues have a partially shared susceptibility to internal and external stress and that dysfunction in one bed is reflected in others^24, 25^.

In addition to the pragmatic utility of developing more readily deployable biomarkers, we propose that our strategy of representing one imaging modality’s output via another may help estimate such a shared susceptibility. We describe our method as a type of “information bottleneck”, consistent with information-theoretic work by Tishby and colleagues, formulated two decades ago^20^, but enjoying recent attention as a possible explanation for the success of deep learning methods^21, 22^. Their work interprets supervised learning as a deliberate process of mapping a complex input signal into a compressed representation but still preserving relevant information about a target. Our application here goes one step further: by placing two modalities (PET and echocardiography) in series, we are driving the underlying physiologic process (i.e., abnormality of coronary blood flow under vasodilatory stress) through an information bottleneck to extract a specific underlying factor that may be partially obscured in the original measurement.

Our genetic correlation work also sheds light on distinctions between stress myocardial blood flow and coronary flow reserve. Previous work has demonstrated independent contributions of these two parameters to cardiovascular outcomes^12^. We found that echoAI-stressMBF had the strongest genetic correlation with ischemic heart disease and a negative correlation with C-reactive protein, a marker of systemic inflammation, in support of prior phenotypic correlation^19, 26^. Among other correlations, the inverse association with rest and stress myocardial blood flow with impedance-estimated fat-free body mass sheds light on previous difficult to reconcile findings. Prior phenotypic association of body mass index with coronary flow reserve and hyperemic blood flow in small groups of patients has been interpreted as the deleterious effects of obesity on endothelial cell function^26^, although body mass index does not unambiguously distinguish between adiposity and muscle mass. Our observed genetic correlation of rest myocardial blood flow with fat-free body mass and standing height (which was further supported by phenotypic data) suggests a complementary hypothesis – there may be competition for blood flow between the heart and remainder of the body: those with greater amounts of metabolically active body mass may have reduced coronary blood flow, independent of any adverse effects of adipose tissue on coronary microcirculation. This observation is in keeping with the previously observed non-monotonic relationship between BMI and CFR^27^, where CFR increases with BMI in the non-obese range but decreases in the obese range. Our results would indicate potential utility in correcting MBF parameters for lean body mass and height to isolate adverse effects of adipose tissue.

The primary limitations of this study are that the PET data were all derived from a single center. However, the utility of the model for predicting cardiovascular events is validated in an external cohort, which showed good discrimination and calibration, suggesting that the model is generalizable to a different institution. A second limitation is that the sample size for genetic association and correlation analysis, though large enough for multiple insights, is also relatively small to discover trait-associated loci.

In summary, we describe the development of scalable imaging-based biomarkers with surprisingly good discriminative properties for cardiovascular outcomes, and more broadly, a generalizable strategy for the discovery of measures of latent biological factors that may extend across multiple tissues.

## Methods

### Selection of temporally coincident PET-CFR and echocardiography cases

Consecutive clinically indicated rest/stress myocardial perfusion PET studies performed at Brigham and Women’s Hospital (Boston, MA) between January 1, 2006 and November 30, 2017, with a clinically indicated echocardiogram within 365 days were included. Patients who had undergone coronary artery bypass grafting (CABG) or cardiac transplantation prior to the PET or the echocardiogram study, were excluded. PET studies were performed with either ^82^Rubidium or ^13^N-ammonia, using computed tomography (CT) for attenuation correction as described previously^28^. Absolute global myocardial blood flow (MBF) was quantified in mL/min/g at rest and at peak hyperemia by a single operator blinded to patient data using a validated two-compartment tracer kinetic model as described previously^29^. Coronary flow reserve (CFR) was calculated as ratio between MBF at stress and MBF at rest. For contemporaneous echocardiograms, we selected all two-dimensional resting echocardiograms that occurred within 365 days of the corresponding PET study.

### Echocardiography pre-processing

The video processing involved 3 steps: (1) frame scaling, (2) frame rate adjustment and (3) square padding. For each echocardiography video, frames were scaled in each dimension by the original pixel spacing factors (in mm/pixel) that were extracted from the DICOM headers to account for variations in resolution. All images were subsequently scaled by a universal scaling factor of 1.177 which was chosen so that the largest dimension of all videos used for training, testing and validation was smaller than the square model input size of 299 pixels. This universal scaling made sure that the model input size of 299 by 299 pixels could be achieved for all videos by simple zero padding while keeping the correct feature sizes. Given that dynamic information, such as velocity of the heart walls, are dependent on frame rate we then set the model input frame rate to a uniform value of 30 frames per second (fps) for all the videos. We excluded all the videos with framerates of that were smaller than 30 fps or had lengths of less than 1.33s (40 frames × 30 fps). All qualifying videos were subsampled by the formula: P_n_=O_n·fps/30_, where P_n_ is the n-th frame of the processed video and O_n_ is the n-th frame of the original video. When the n·fps/30 term was a non-integer value, it was rounded to the closest integer. Videos were saved together with the corresponding labels (CFR and MBF values) as 3-D arrays in the TensorFlow TFRecord serialized format to be used by input pipeline. During training, images and labels were re-shuffled before each epoch, distributed across all 8 GPUs and finally zero-padded to the required square model input size of 299 by 299 pixels without further scaling.

We used TensorFlow (version 2.2.0) for implementation and deployment of the neural network models for myocardial blood flow indices. Given that the echocardiograms were time-series of multiple frames, we constructed a 3D-CNN model treating the temporal axis as the 3rd dimension, to maximize the ability of the model to use dynamic features. It consisted of 3 layers of 3D-CNN followed by 12 layers with Multi-3D-CNN-modules, which was constructed by 3 parallel multilayer 3D-CNNs and a max pooling operation concatenated at the end of the module (schematic shown in **Figure S12**). The model was trained using data from the training cohort (**Figure S1**). The echocardiography videos were labeled with the PET-measured CFR or MBF value on the study level and the model was trained to minimize the mean-squared-error between model prediction and the label using an RMSprop optimizer with an initial learning rate of 0.0001. The model was trained for 300 epochs with a batch size of 64 videos distributed across 8 NVIDIA Tesla V100 SXM2 32GB GPUs. At the end of each epoch, the Spearman correlation coefficient was calculated using the validation set. The final models were chosen as the models with the highest correlation coefficient on the validation cohort across all 300 epochs. The code to implement the models along with instructions for installation/usage are provided in a public repository https://github.com/obi-ml-public/echoAI-PET-measurements.

### Cohort selection for event analysis

The Massachusetts General Brigham Electronic Data Warehouse (EDW), which includes notes from >20 provider locations across a large integrated delivery network, was the source of notes and structured clinical data used in this study. All information in the electronic health record in the EDW system was gathered including free text medical notes and discharge summaries, and outcome events were adjudicated using a machine learning based model using the discharge summaries as input as described previously^30^. Briefly, after manually labeling 1,372 training, 592 validation and 1,003 test set discharge summaries, we trained neural network models for event adjudication for heart failure hospitalization (test set AUROC 0.97) and ACS (test set AUROC 0.98). We selected classification thresholds to maximize sensitivity at specificity over 0.90 (HF: cutoff 0.12, sensitivity 0.94, specificity 0.91. ACS: cutoff 0.04, sensitivity 0.91, specificity 0.91). The discharge summary classification models were deployed on 44,150 notes and a binary event status was determined by each model for each note.

To test if the echoAI measurements can predict clinical outcomes, survival analysis was performed using patients who had echocardiography within Jun 2015 and Nov 2015 at BWH. The first echocardiography study for each patient was identified and the echoAI models were ran on those videos to obtain the corresponding measurements. Follow up for each patient was determined using hospital and clinic encounters within the system. Patients were excluded if they had (1) no notes in the system, (2) less than 30 days follow up or (3) had an event during the first 30 days. To avoid the possible bias caused by the influence of COVID-19, all the patients were censored at Feb 2019. An external validation cohort was selected with similar procedure from patients who had at least one echo cardiogram from MGH (**Figure S7**). Since MGH started to enter notes from Feb 2016, the inclusion period was set to Feb 2016 to Nov 2016 for MGH.

### Analysis of predictive accuracy of echo-AI rest MBF, echo-AI stress MBF and echo AI CFR for heart failure hospitalization

Receiver operating characteristics (ROC) curves for the echoAI measurements along with ejection fractions, heart rate and 4 patient demographic variables (age, sex, heart failure history and CAD history) were calculated using timeROC package version 0.4. A multivariable Cox proportional hazard (PH) model including all 9 parameters were trained using survival package (version 3.2.3) and the ROC curve for the model output was calculated with the timeROC package. The individual hazard ratios for the multivariable Cox PH model were analyzed to assess the adjusted association of each parameter to the outcome. For external validation, two separate Cox PH models were used. To assess the ability of the 9-parameter model to predict outcomes, the original Cox PH models trained on BWH data were used without any weight adjustment. However, to analyze the individual adjusted hazard ratios for the echoAI measures along with 6 baseline characteristics information, we trained separate Cox PH models with the same input parameters on MGH data.

### Genetic association analysis

Genomewide SNP array data is available for over 36,000 individuals enrolled in the Massachusetts General Brigham Biobank^31^, a collection of plasma, serum, and DNA samples of consented subjects with associated linkage to their electronic health records. We downloaded echocardiograms for 4,400 of these patients and were able to compute echoAI-CFR and echoAI-stressMBF on 4,145. To minimize confounding by population structure, we focused our genetic association analyses on 3,925 white study subjects. We performed a genomewide association study using age, sex, and the first 10 principal components as covariates using PLINK 2.0^32^. Summary statistics were used to estimate narrow-sense heritability with the help of the R *HDL* package^33^. Genetic correlation with a prioritized selection of traits from the UK Biobank was also performed using summary statistics provided by the Neale Lab (http://www.nealelab.is/uk-biobank/). Genetic correlation p-values for UK Biobank traits were adjusted for multiple hypothesis testing using the R *qvalue* package^34^. We set a false-discovery threshold for “significance” at 0.20, though any such threshold is arbitrary and should be driven by downstream decisions related to the corresponding finding.

### Statistical analysis

Data were collected and stored using Numpy package version 1.19.2 with Python 3.7.3. The scatter plots and ROC curves are plotted using the *ggplot2*^35^ package (R 3.6.1). Continuous values are presented as mean ± standard deviation (SD) and categorical values are presented as numbers and percentages if not otherwise specified.

### Ethics statement

This study complies with all ethical regulations and guidelines. The study protocol was approved by local institutional review board (IRB) of Mass General Brigham (2019P002651). This study had minimal patient risk: it collected data retrospectively, there was no direct contact with patients, and data were collected after medical care was completed. Thus, and to recruit an unbiased and representative cohort of patients, data were collected under a waiver of informed consent, which was approved by the IRB.

### Data availability

The data that support the findings of this study are available on request from the corresponding author R.C.D. upon approval of the data sharing committees. The data are not publicly available due to the presence of information that could compromise research participant privacy.

### Code availability

The code to run the models along with instructions for installation/usage are provided in a public repository https://github.com/obi-ml-public/echoAI-PET-measurements. The model weights may contain patient personal information and thus, could not be shared. We provide a web-interface to run our model and generate predictions at http://onebraveideaml.org

## Supporting information

Genetic Correlation Data

## Data Availability

https://github.com/obi-ml-public/echoAI-PET-measurements

http://onebraveideaml.org

## Acknowledgement

This work was supported by One Brave Idea, co-founded by the American Heart Association and Verily with significant support from AstraZeneca and pillar support from Quest Diagnostics (to CAM and RCD), NIH/NHLBI HL140731 (to RCD) SENSHIN Medical Research Foundation (to SG), the Kanae foundation for the promotion of medical science (to SG), the Uehara Memorial Foundation (to SG), SG acknowledge support from Mower fellowship.

## Supplementary Figures

**Figure S1.**
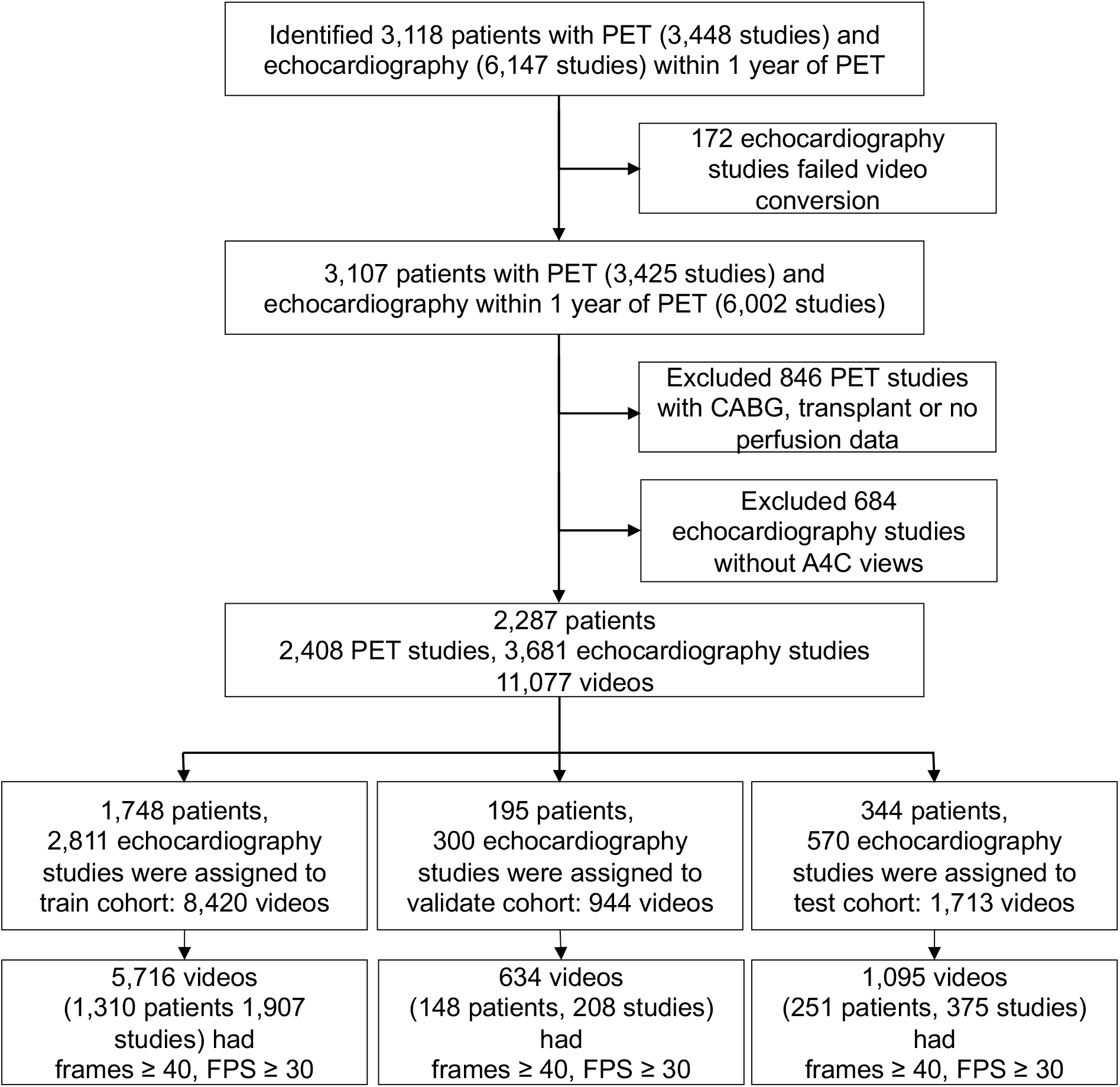
Consort diagram of echo selections for echoAI-CFR model training and testing.

**Figure S2.**
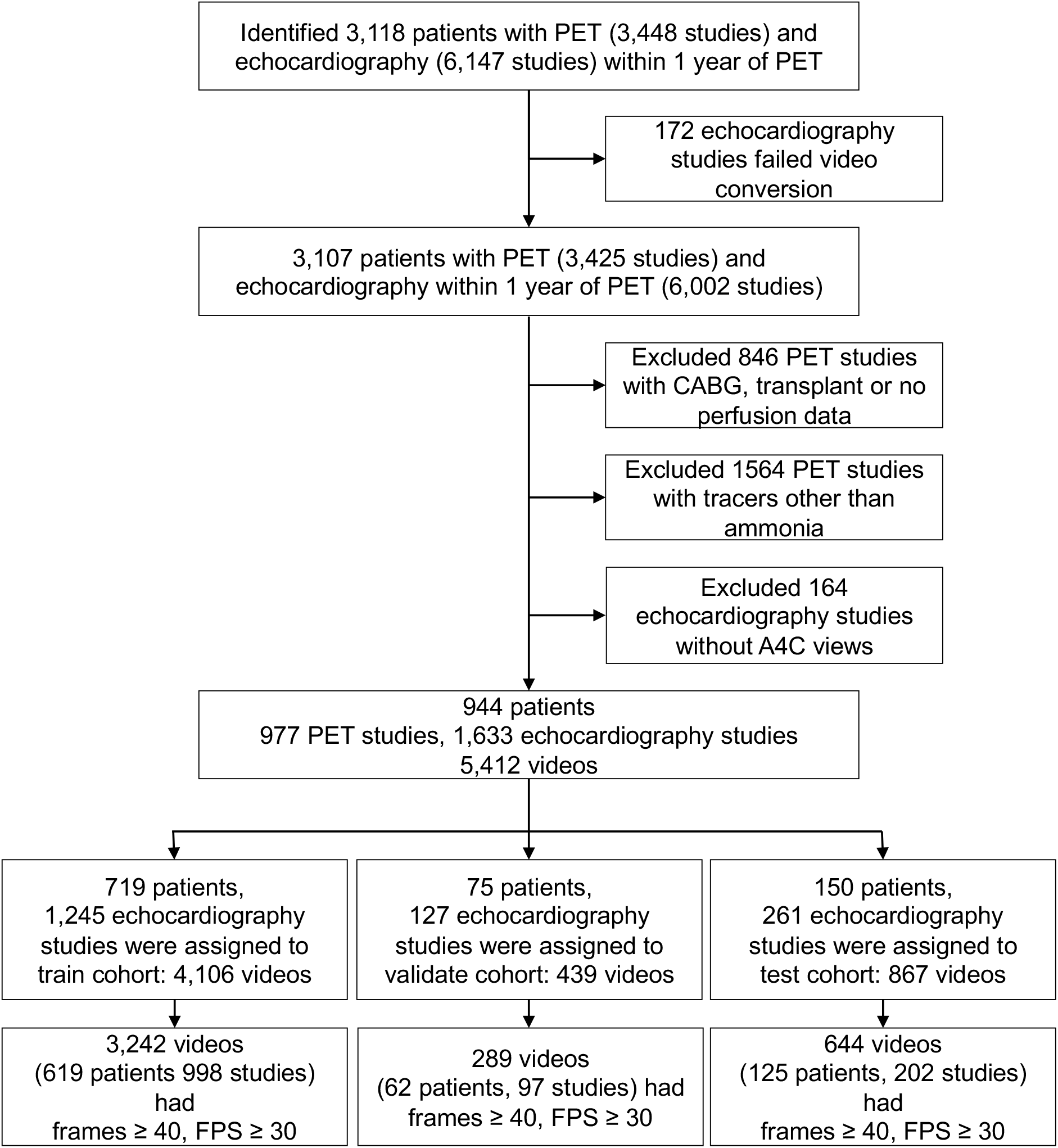
Consort diagram of echo selections for echoAI-restMBF and echoAI-stressMBF model training and testing.

**Figure S3.**
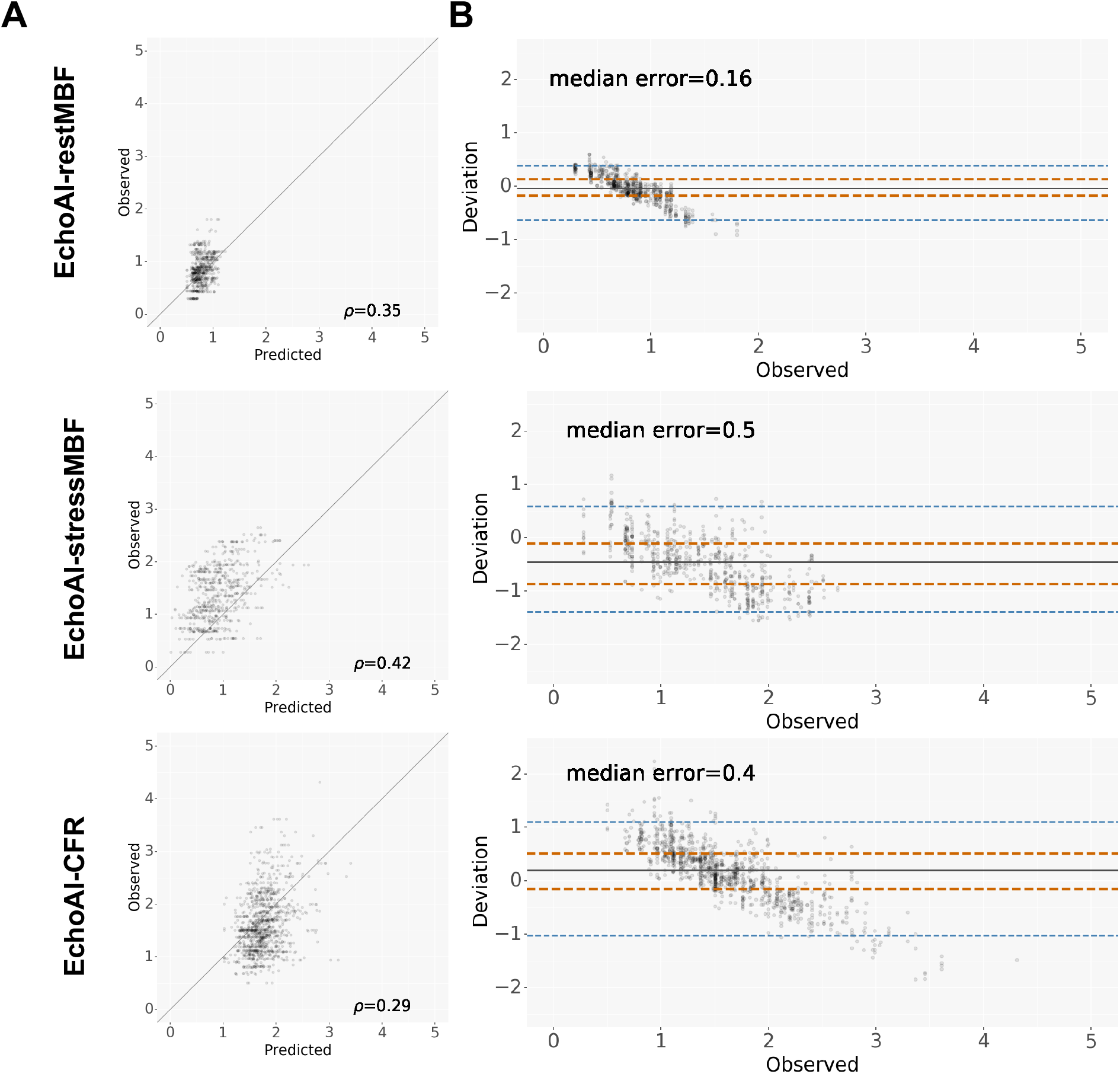
Agreement of EchoAI measurements with the PET measurements. (A) Scatter plots and (B) Bland-Altman plot for stress MBF, rest MBF and CFR. Observed represents the PET measured value and Predicted represents the echo-AI predicted value. Abbreviations, PET: positron emission tomography, MBF: myocardial blood flow, CFR: coronary flow reserve.

**Figure S4.**
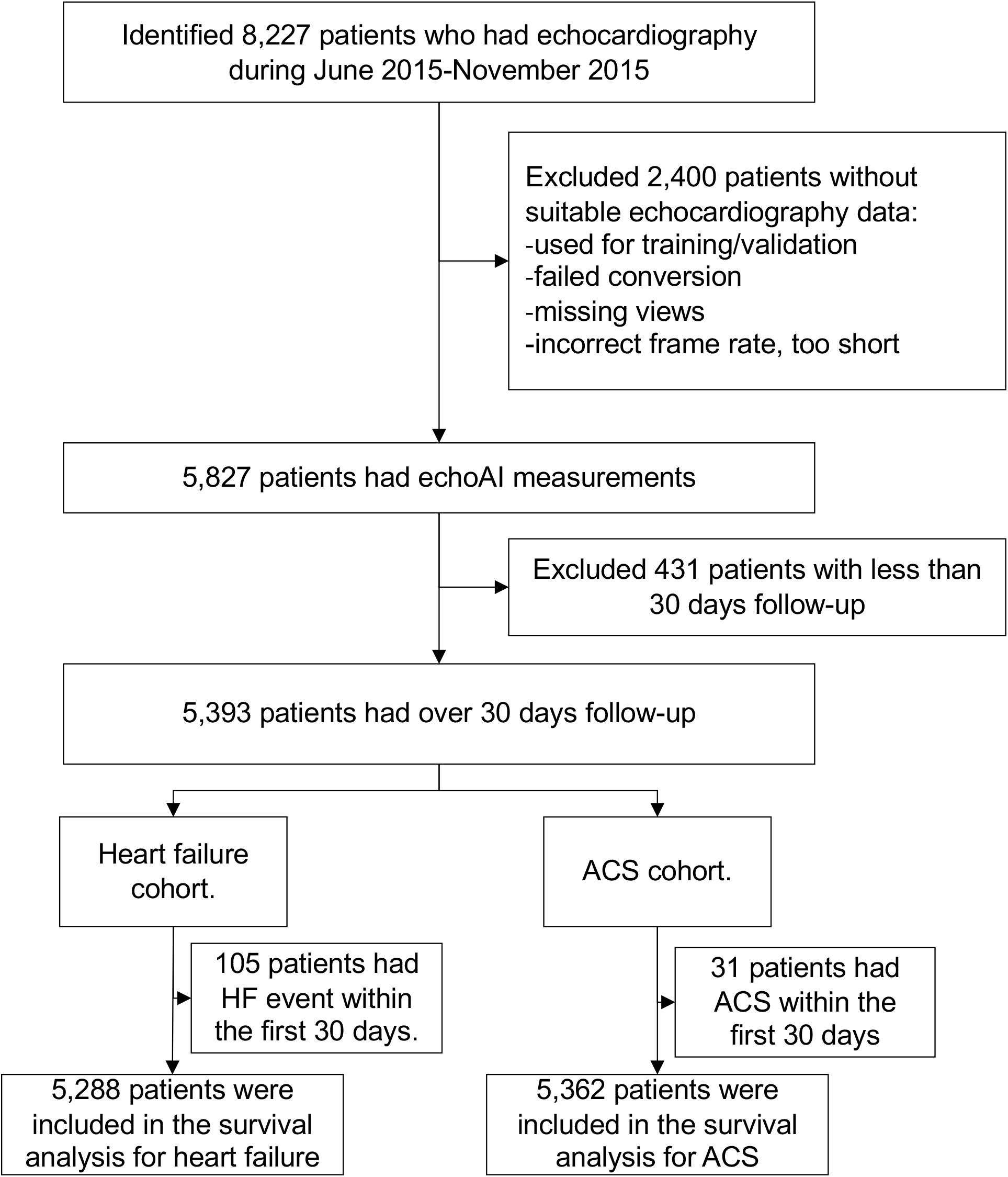
Consort diagram of patient selection for survival analysis.

**Figure S5.**
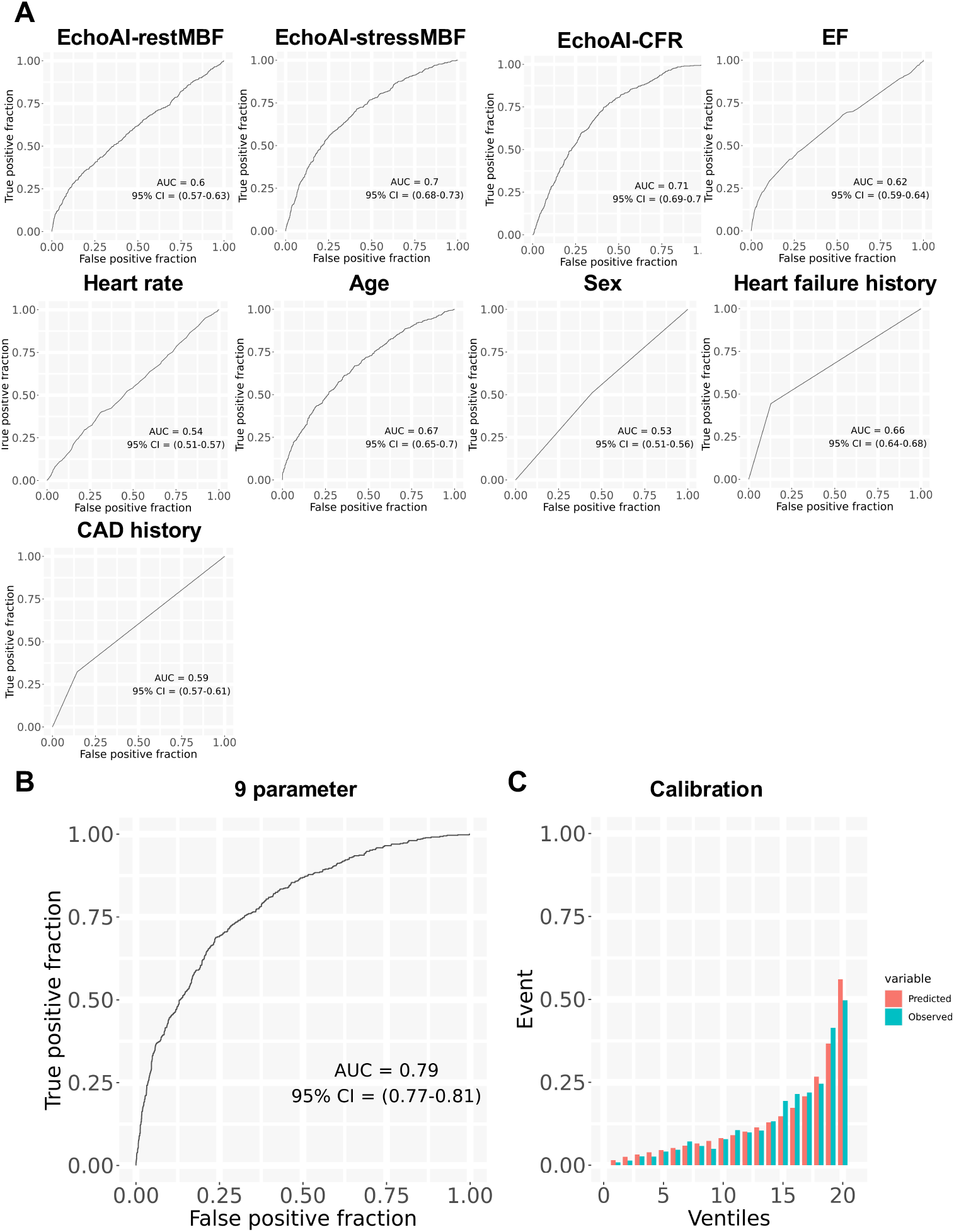
Predictive accuracy of echoAI measurements and various patient characteristics for heart failure hospitalization. (A) Receiver operating characteristics curves for the echoAI parameters, ejection fraction, heart rate and other patients’ demographics information to predict heart failure hospitalization. (B) Receiver operating characteristics curve and (C) calibration plot for the 9-parameter combined model to predict heart failure hospitalization. Abbreviations, EF: ejection fraction, CAD: coronary artery disease, MBF: myocardial blood flow, CFR: coronary flow reserve.

**Figure S6.**
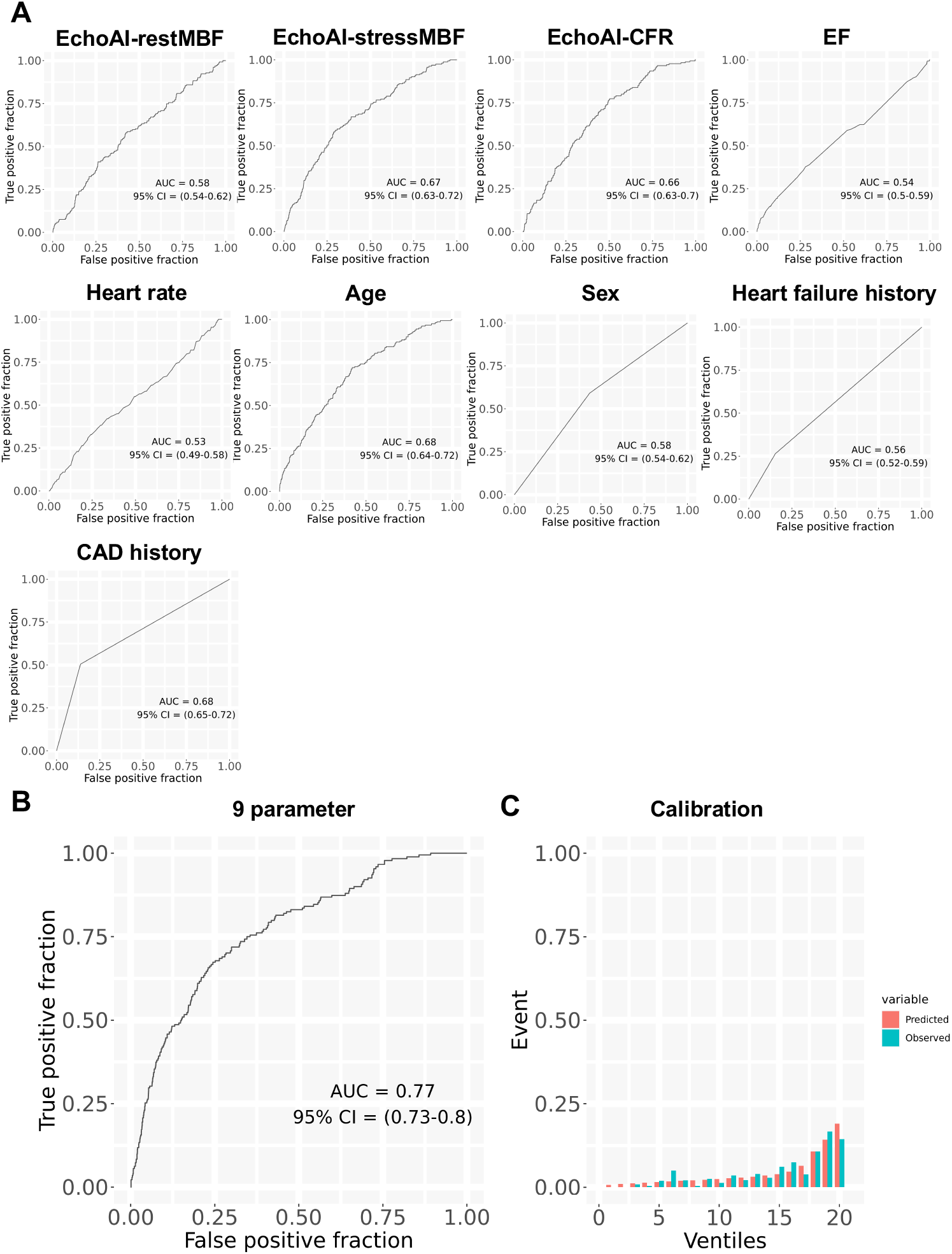
Predictive accuracy of echoAI measurements and various patient characteristics for ACS. (A) Receiver operating characteristics curves for the echoAI parameters, ejection fraction, heart rate and other patients’ demographics information to predict ACS. (B) Receiver operating characteristics curve and (C) calibration plot for the 9 parameters combined model to predict ACS. Abbreviations, ACS: acute coronary syndrome, EF: ejection fraction, CAD: coronary artery disease, MBF: myocardial blood flow, CFR: coronary flow reserve.

**Figure S7.**
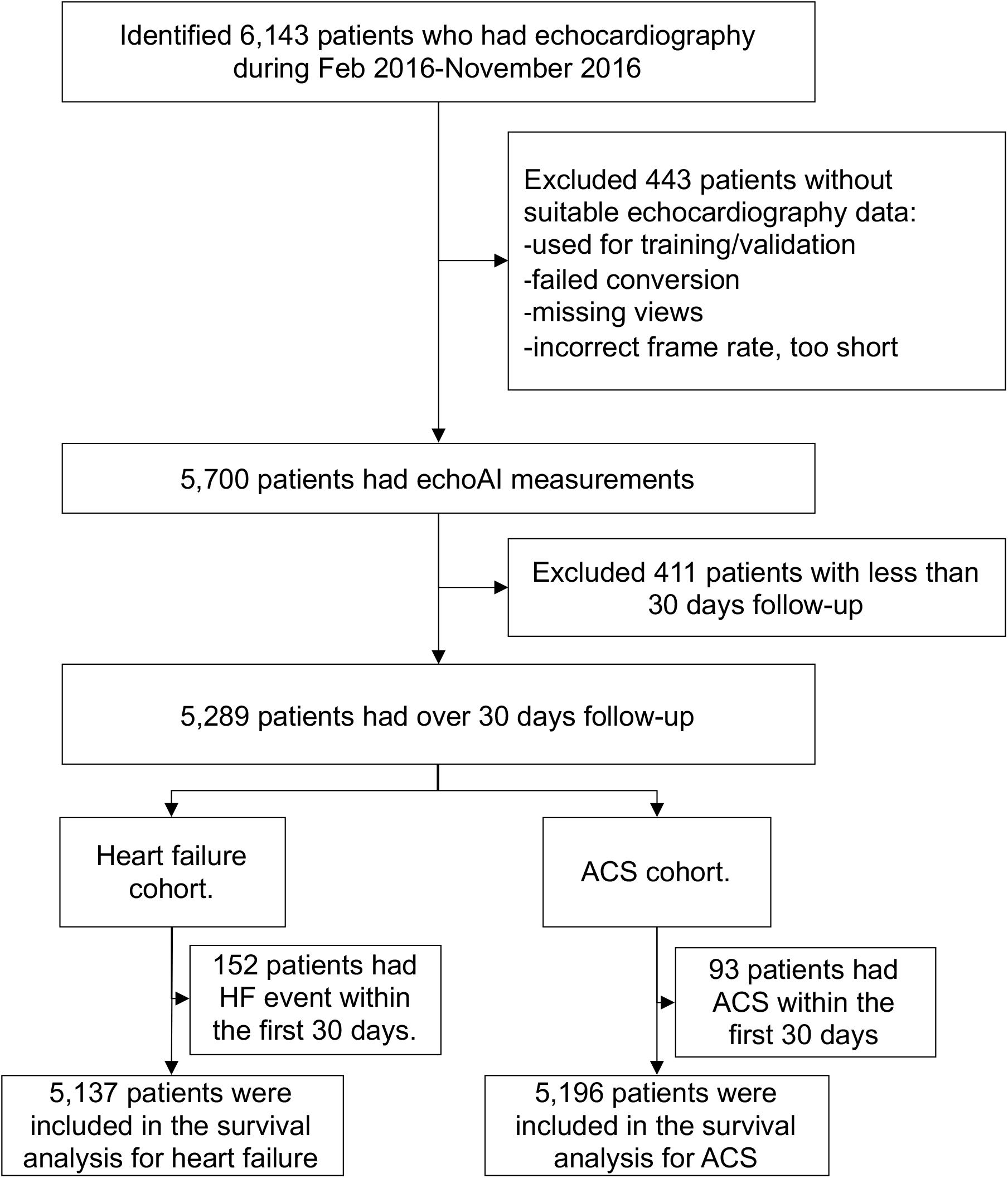
Consort diagram of patient selection for survival analysis in external validation cohort from MGH.

**Figure S8.**
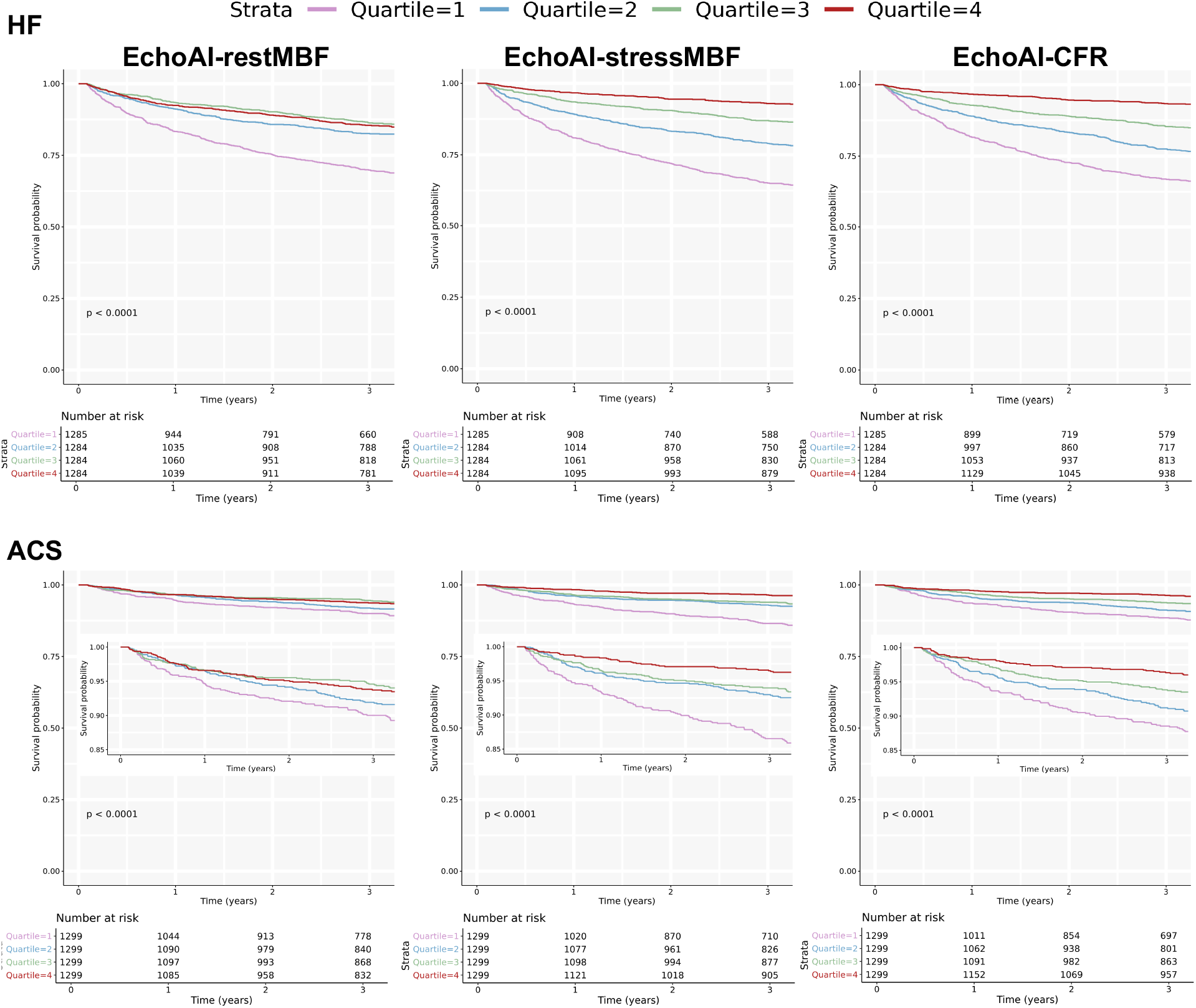
Cardiovascular event free survival for echo-AI measurements quartiles in external validation cohort from MGH. Kaplan-Meier plots for HF hospitalization and ACS free survival for echoAI measurement quartiles. MGH: Massachusetts General Hospital.

**Figure S9.**
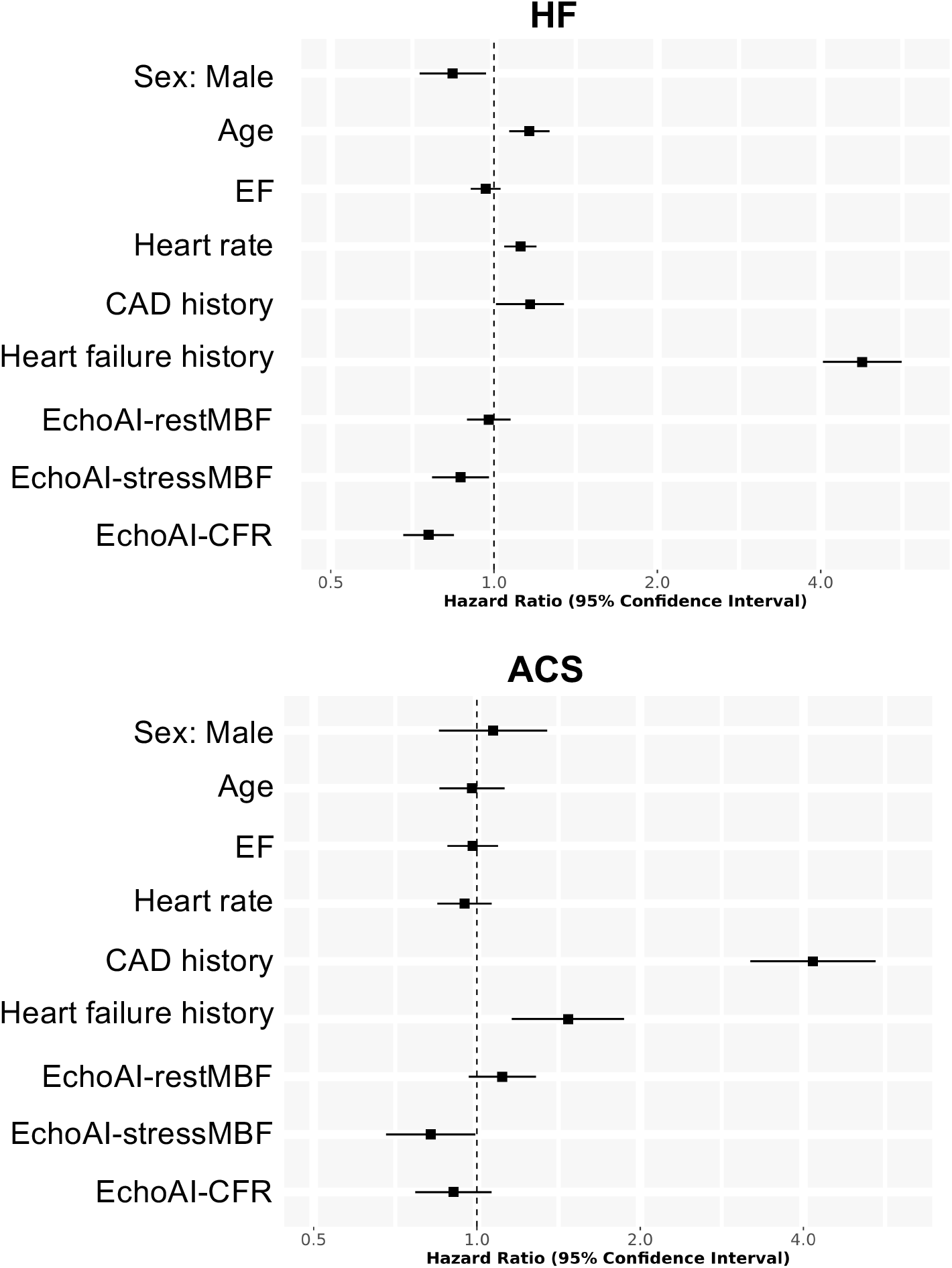
Hazard ratios of echoAI measurements and various patient characteristics for the multivariable Cox proportional hazard models in external validation cohort from MGH. Forest plot showing the hazard ratio and 95% confidence intervals for each parameter in the Cox proportional hazard models for heart failure hospitalization and ACS. Hazard ratios are plotted as per standard deviation. Abbreviations, ACS: acute coronary syndrome, EF: ejection fraction, CAD: coronary artery disease, MBF: myocardial blood flow, CFR: coronary flow reserve, MGH: Massachusetts General Hospital.

**Figure S10.**
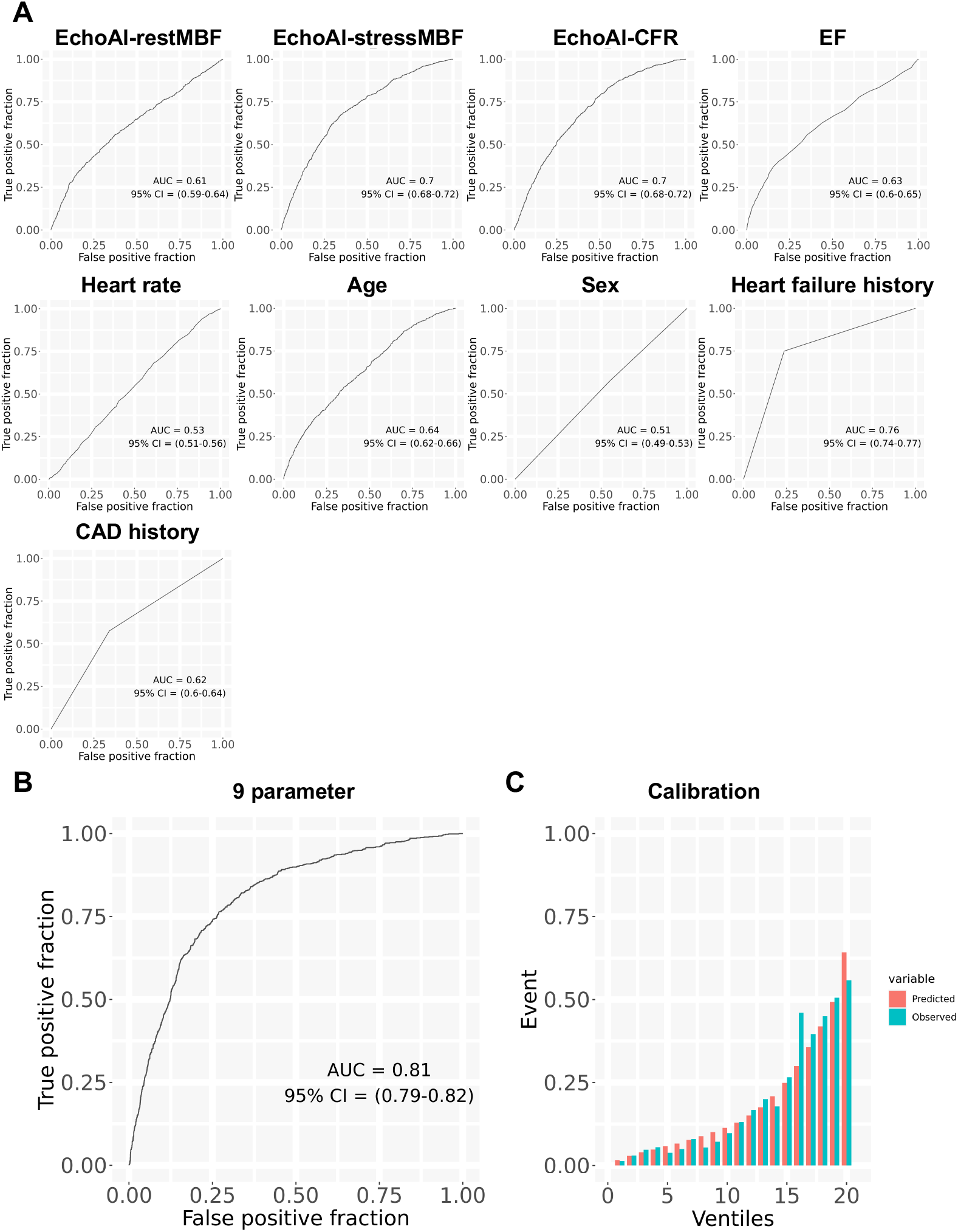
Predictive accuracy of echoAI measurements and various patient characteristics for heart failure hospitalization in external validation cohort from MGH. (A) Receiver operating characteristics curves for the echoAI parameters, ejection fraction, heart rate and other patients’ demographics information to predict heart failure hospitalization. (B) Receiver operating characteristics curve and (C) calibration plot for the 9 parameters combined model to predict heart failure hospitalization. Abbreviations, EF: ejection fraction, CAD: coronary artery disease, MBF: myocardial blood flow, CFR: coronary flow reserve.

**Figure S11.**
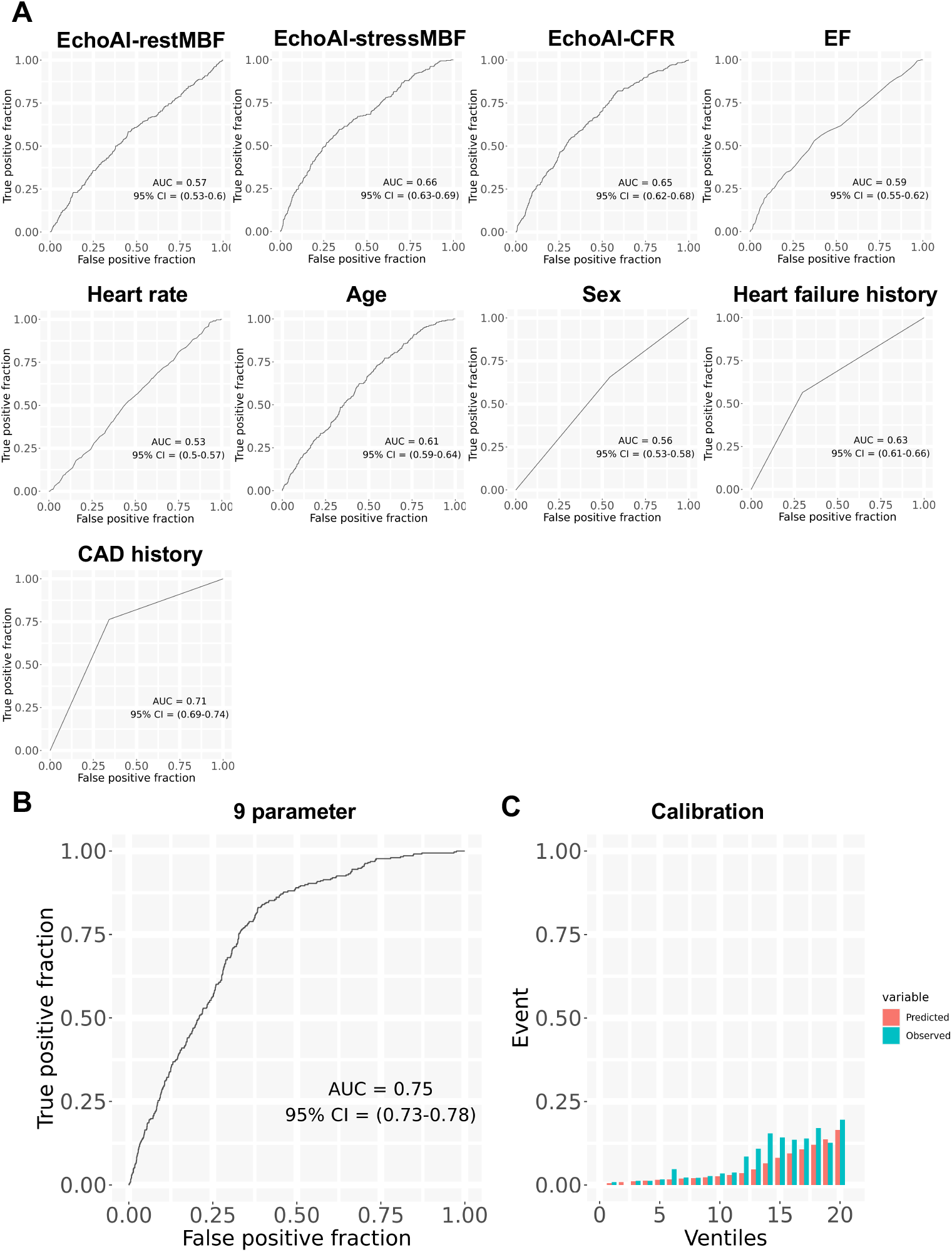
Predictive accuracy of echoAI measurements and various patient characteristics for ACS in external validation cohort from MGH. (A) Receiver operating characteristics curves for the echoAI parameters, ejection fraction, heart rate and other patients’ demographics information to predict ACS. (B) Receiver operating characteristics curve and (C) calibration plot for the 9 parameters combined model to predict ACS. Abbreviations, ACS: acute coronary syndrome, EF: ejection fraction, CAD: coronary artery disease, MBF: myocardial blood flow, CFR: coronary flow reserve.

**Figure S12.**
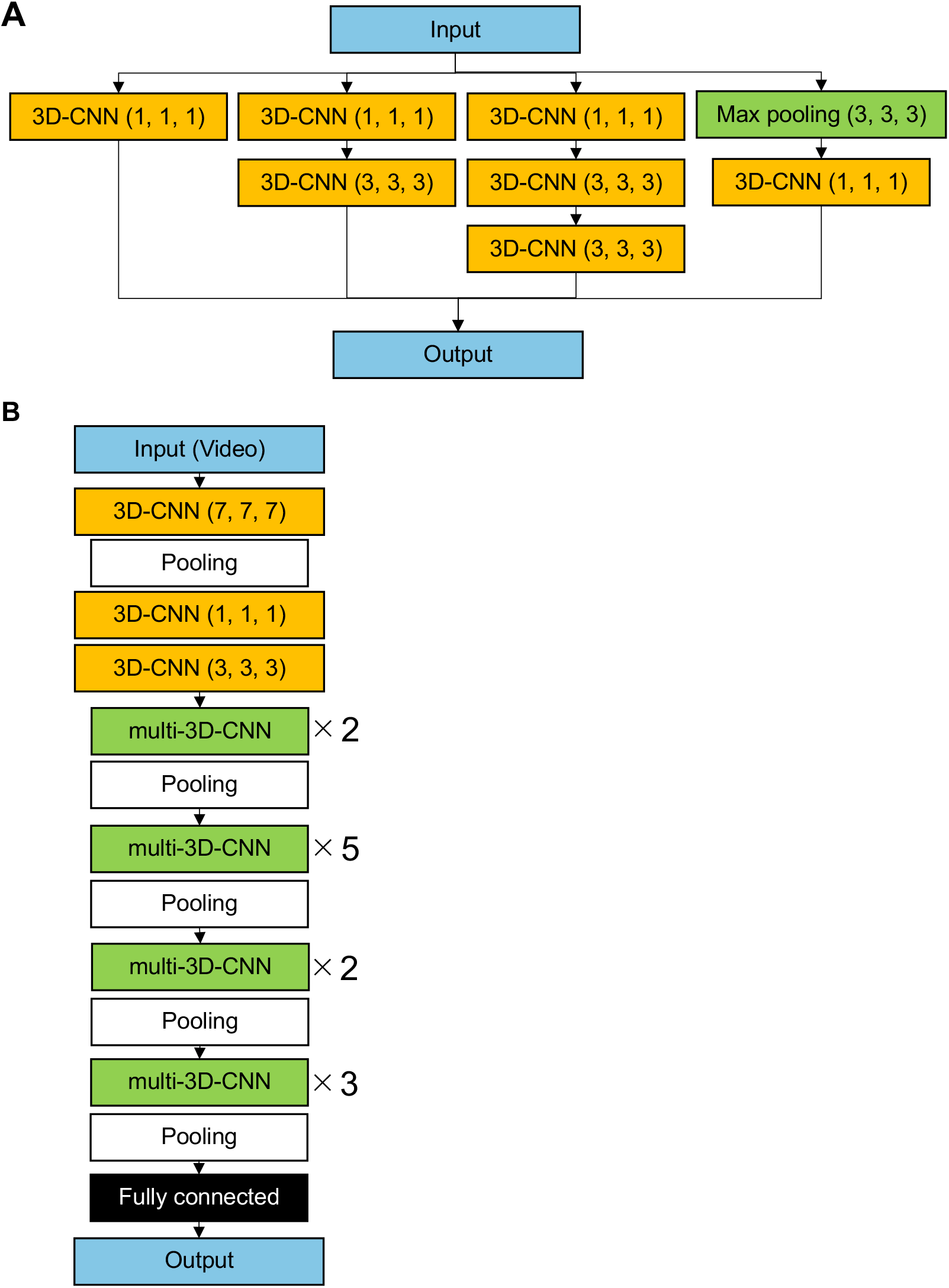
Schematic illustration of the model architecture. The model architecture for the (A) multi-CNN module and the (B) overall model. Abbreviations, CNN: convolutional neural network

## Supplementary Tables

**Table S1.**
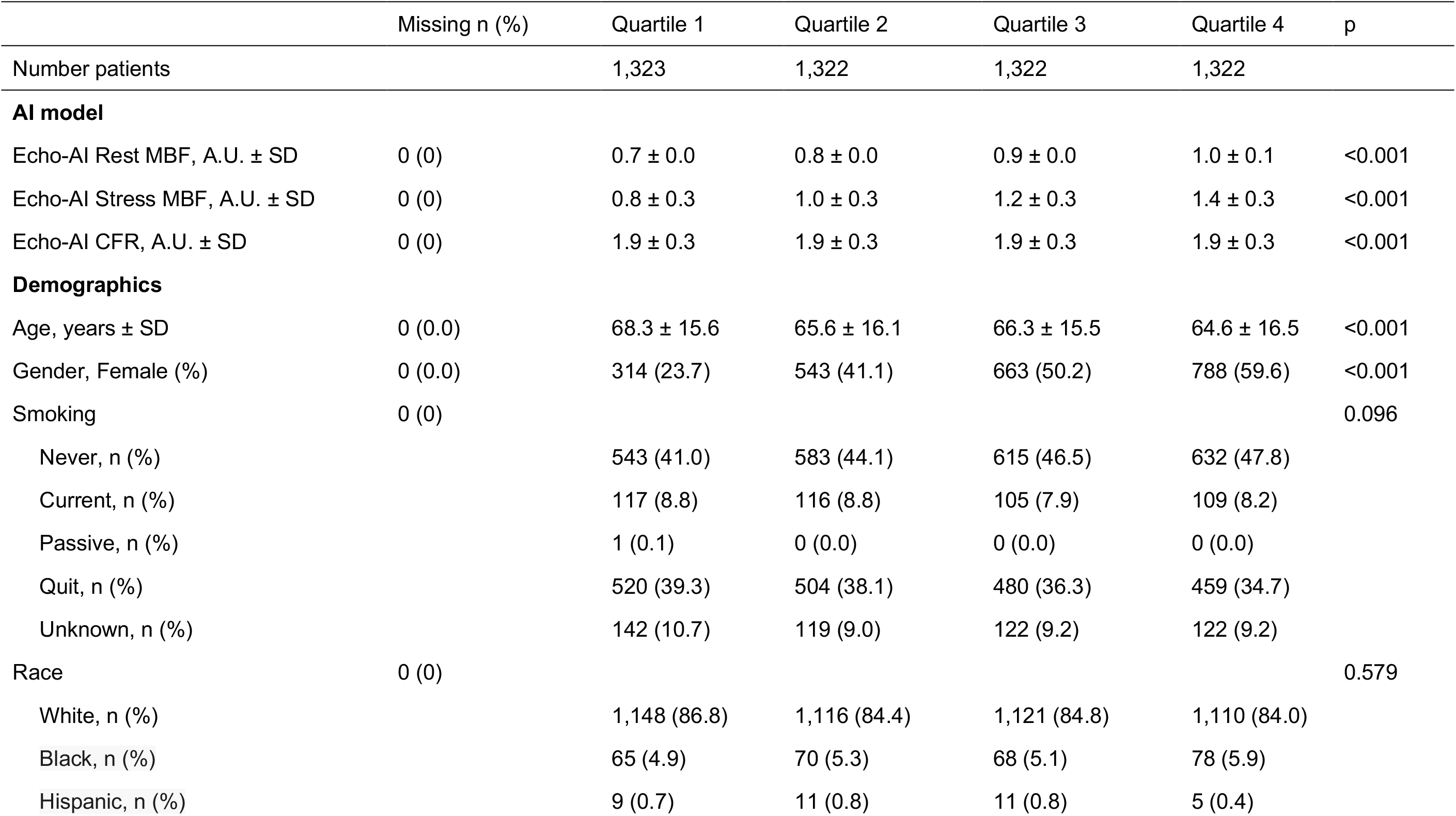
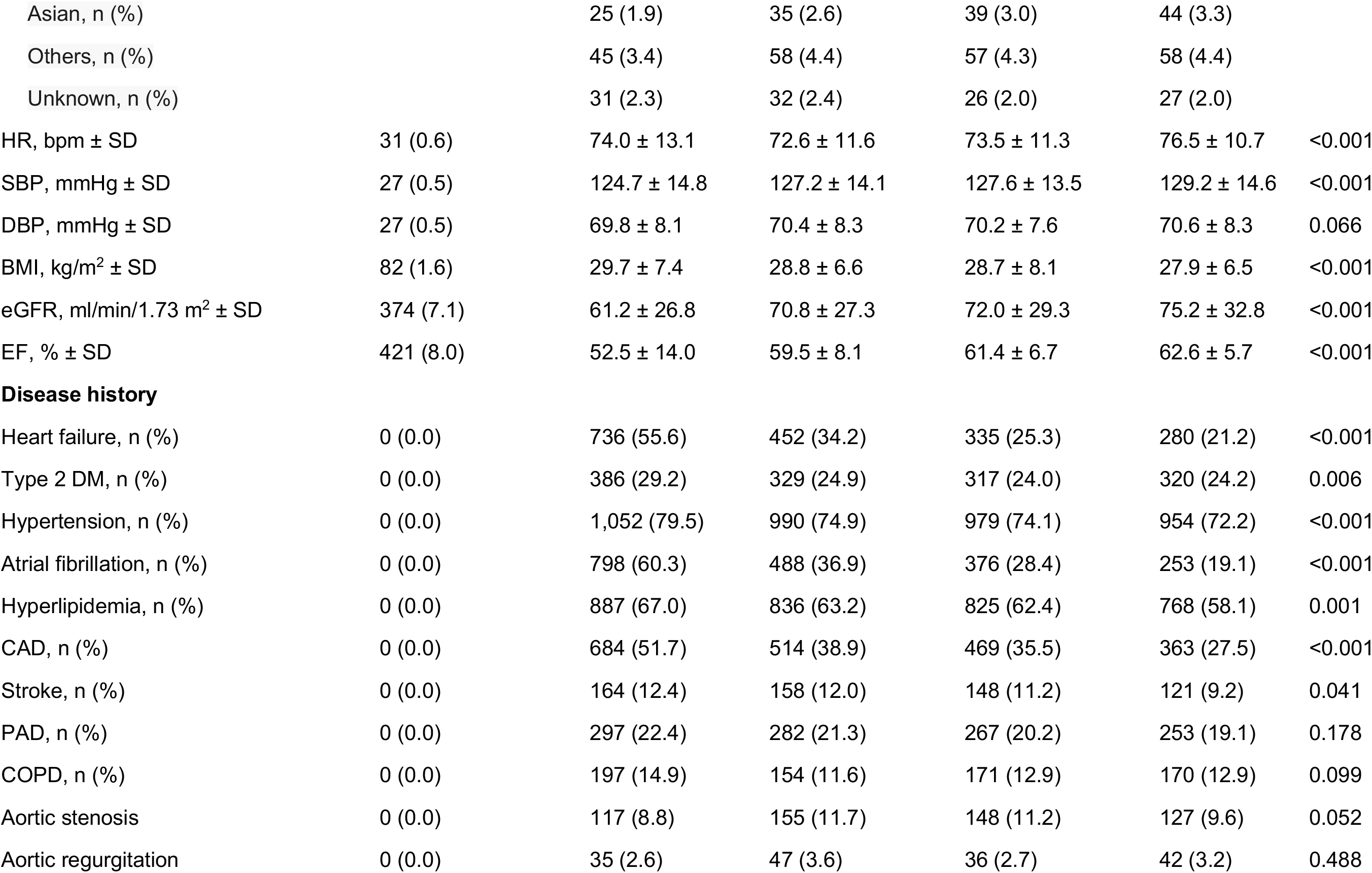
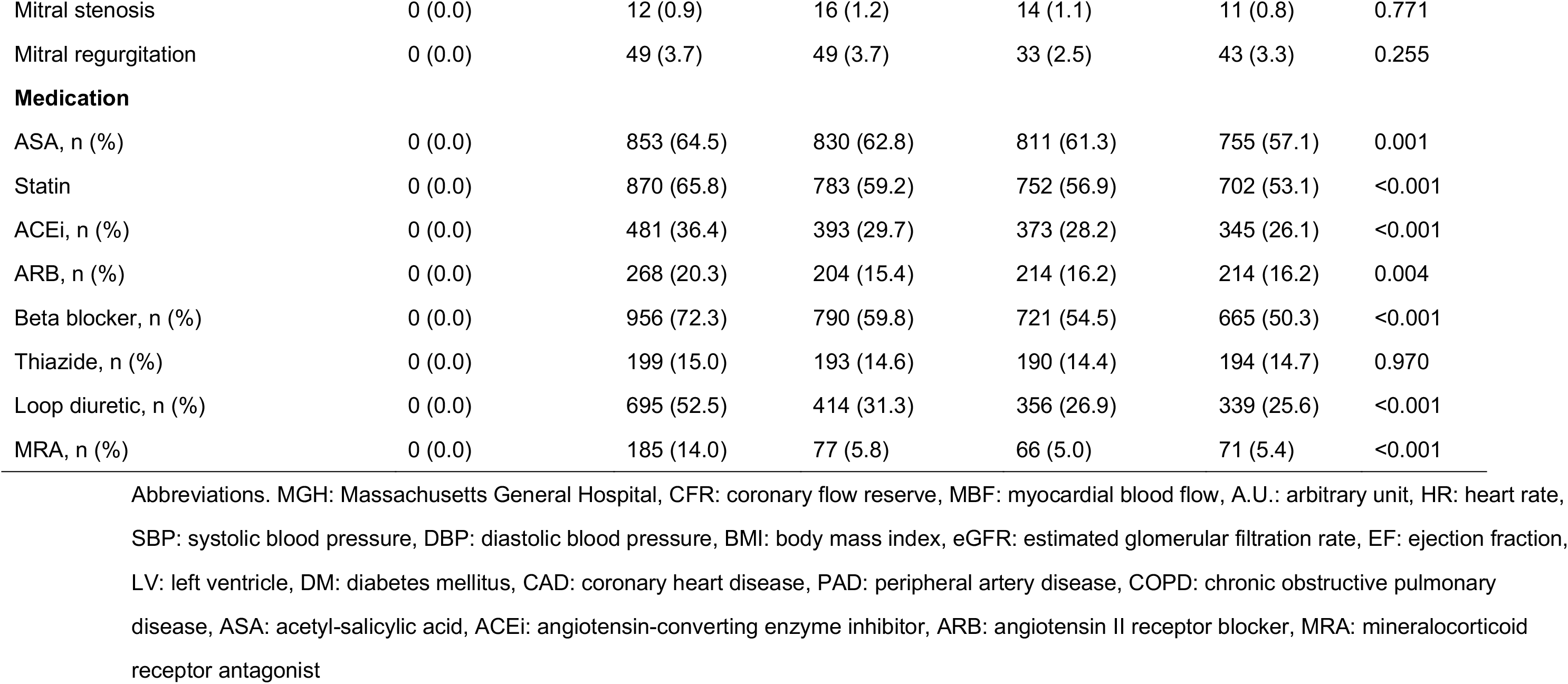
Baseline demographic for EchoAI-restMBF quartiles (MGH cohort)

**Table S2.** Baseline demographic for EchoAI-stressMBF quartiles (MGH cohort)

|  | Missing n (%) | Quartile 1 | Quartile 2 | Quartile 3 | Quartile 4 | p |
| --- | --- | --- | --- | --- | --- | --- |
| Number patients |  | 1,323 | 1,322 | 1,322 | 1,322 |  |
| **AI model** |  |  |  |  |  |  |
| Echo-AI Rest MBF, A.U. ± SD | 0 (0) | 0.7 ± 0.1 | 0.8 ± 0.1 | 0.9 ± 0.1 | 0.9 ± 0.1 | <0.001 |
| Echo-AI Stress MBF, A.U. ± SD | 0 (0) | 0.6 ± 0.1 | 0.9 ± 0.1 | 1.2 ± 0.1 | 1.6 ± 0.2 | <0.001 |
| Echo-AI CFR, A.U. ± SD | 0 (0) | 1.7 ± 0.2 | 1.8 ± 0.2 | 2.0 ± 0.2 | 2.2 ± 0.3 | <0.001 |
| **Demographics** |  |  |  |  |  |  |
| Age, years ± SD | 0 (0.0) | 71.8 ± 13.5 | 69.6 ± 14.2 | 65.7 ± 15.4 | 57.7 ± 17.0 | <0.001 |
| Gender, Female (%) | 0 (0.0) | 353 (26.7) | 542 (41.0) | 602 (45.5) | 811 (61.3) | <0.001 |
| Smoking | 0 (0) |  |  |  |  | <0.001 |
| Never, n (%) |  | 525 (39.7) | 578 (43.7) | 602 (45.5) | 668 (50.5) |  |
| Current, n (%) |  | 87 (6.6) | 102 (7.7) | 117 (8.9) | 141 (10.7) |  |
| Passive, n (%) |  | 0 (0.1) | 1 (0.1) | 0 (0.0) | 0 (0.0) |  |
| Quit, n (%) |  | 567 (42.9) | 514 (38.9) | 478 (36.2) | 404 (30.6) |  |
| Unknown, n (%) |  | 144 (10.9) | 127 (9.6) | 125 (9.5) | 109 (8.2) |  |
| Race | 0 (0) |  |  |  |  | 0.013 |
| White, n (%) |  | 1,152 (87.1) | 1,148 (86.8) | 1,105 (83.6) | 1,090 (82.5) |  |
| Black, n (%) |  | 71 (5.4) | 57 (4.3) | 74 (5.6) | 79 (6.0) |  |
| Hispanic, n (%) |  | 7 (0.5) | 9 (0.7) | 8 (0.6) | 12 (0.9) |  |
| Asian, n (%) |  | 21 (1.6) | 34 (2.6) | 37 (2.8) | 51 (3.9) |  |
| Others, n (%) |  | 47 (3.6) | 42 (3.2) | 66 (5.0) | 63 (4.8) |  |
| Unknown, n (%) |  | 25 (1.9) | 32 (2.4) | 32 (2.4) | 27 (2.0) |  |
| HR, bpm ± SD | 31 (0.6) | 72.9 ± 11.9 | 72.4 ± 11.6 | 73.3 ± 11.5 | 78.2 ± 11.3 | <0.001 |
| SBP, mmHg ± SD | 27 (0.5) | 126.9 ± 15.5 | 128.4 ± 14.2 | 127.5 ± 13.7 | 125.9 ± 13.9 | <0.001 |
| DBP, mmHg ± SD | 27 (0.5) | 68.9 ± 8.2 | 69.9 ± 7.7 | 70.9 ± 8.0 | 71.3 ± 8.3 | <0.001 |
| BMI, kg/m<sup>2</sup> ± SD | 82 (1.6) | 29.8 ± 8.3 | 29.3 ± 7.3 | 28.6 ± 6.4 | 27.2 ± 7.0 | <0.001 |
| eGFR, ml/min/1.73 m<sup>2</sup> ± SD | 374 (7.1) | 56.2 ± 25.7 | 64.7 ± 25.9 | 72.8 ± 28.7 | 85.4 ± 29.9 | <0.001 |
| EF, % ± SD | 421 (8.0) | 53.2 ± 16.9 | 63.0 ± 11.7 | 66.1 ± 8.8 | 67.4 ± 7.1 | <0.001 |
| **Disease history** |  |  |  |  |  |  |
| Heart failure, n (%) | 0 (0.0) | 834 (63.0) | 536 (40.5) | 310 (23.4) | 123 (9.3) | <0.001 |
| Type 2 DM, n (%) | 0 (0.0) | 498 (37.6) | 352 (26.6) | 293 (22.2) | 209 (15.8) | <0.001 |
| Hypertension, n (%) | 0 (0.0) | 1,145 (86.5) | 1,064 (80.5) | 985 (74.5) | 781 (59.1) | <0.001 |
| Atrial fibrillation, n (%) | 0 (0.0) | 805 (60.8) | 597 (45.2) | 358 (27.1) | 155 (11.7) | <0.001 |
| Hyperlipidemia, n (%) | 0 (0.0) | 967 (73.1) | 910 (68.8) | 830 (62.8) | 609 (46.1) | <0.001 |
| CAD, n (%) | 0 (0.0) | 810 (61.2) | 577 (43.6) | 443 (33.5) | 200 (15.1) | <0.001 |
| Stroke, n (%) | 0 (0.0) | 198 (15.0) | 150 (11.3) | 135 (10.2) | 108 (8.2) | <0.001 |
| PAD, n (%) | 0 (0.0) | 341 (25.8) | 333 (25.2) | 237 (17.9) | 188 (14.2) | <0.001 |
| COPD, n (%) | 0 (0.0) | 207 (15.6) | 177 (13.4) | 162 (12.3) | 146 (11.0) | 0.004 |
| Aortic stenosis | 0 (0.0) | 176 (13.3) | 162 (12.3) | 142 (10.7) | 67 (5.1) | <0.001 |
| Aortic regurgitation | 0 (0.0) | 49 (3.7) | 50 (3.8) | 36 (2.7) | 25 (1.9) | 0.012 |
| Mitral stenosis | 0 (0.0) | 16 (1.2) | 18 (1.4) | 10 (0.8) | 9 (0.7) | 0.215 |
| Mitral regurgitation | 0 (0.0) | 61 (4.6) | 48 (3.6) | 44 (3.3) | 21 (1.6) | <0.001 |
| **Medication** |  |  |  |  |  |  |
| ASA, n (%) | 0 (0.0) | 970 (73.3) | 882 (66.7) | 838 (63.4) | 559 (42.3) | <0.001 |
| Statin | 0 (0.0) | 992 (75.0) | 855 (64.7) | 765 (57.9) | 495 (37.4) | <0.001 |
| ACEi, n (%) | 0 (0.0) | 520 (39.3) | 407 (30.8) | 373 (28.2) | 292 (22.1) | <0.001 |
| ARB, n (%) | 0 (0.0) | 284 (21.5) | 238 (18.0) | 225 (17.0) | 153 (11.6) | <0.001 |
| Beta blocker, n (%) | 0 (0.0) | 1,052 (79.5) | 906 (68.5) | 715 (54.1) | 459 (34.7) | <0.001 |
| Thiazide, n (%) | 0 (0.0) | 222 (16.8) | 214 (16.2) | 196 (14.8) | 144 (10.9) | <0.001 |
| Loop diuretic, n (%) | 0 (0.0) | 779 (58.9) | 514 (38.9) | 310 (23.4) | 201 (15.2) | <0.001 |
| MRA, n (%) | 0 (0.0) | 184 (13.9) | 89 (6.7) | 72 (5.4) | 54 (4.1) | <0.001 |

Abbreviations. MGH: Massachusetts General Hospital, CFR: coronary flow reserve, MBF: myocardial blood flow, A.U.: arbitrary unit, HR: heart rate, SBP: systolic blood pressure, DBP: diastolic blood pressure, BMI: body mass index, eGFR: estimated glomerular filtration rate, EF: ejection fraction, LV: left ventricle, DM: diabetes mellitus, CAD: coronary heart disease, PAD: peripheral artery disease, COPD: chronic obstructive pulmonary disease, ASA: acetyl-salicylic acid, ACEi: angiotensin-converting enzyme inhibitor, ARB: angiotensin II receptor blocker, MRA: mineralocorticoid receptor antagonist

**Table S3.** Baseline demographic for EchoAI-CFR quartiles (MGH cohort)

|  | Missing n (%) | Quartile 1 | Quartile 2 | Quartile 3 | Quartile 4 | p |
| --- | --- | --- | --- | --- | --- | --- |
| Number patients |  | 1,323 | 1,322 | 1,322 | 1,322 |  |
| **AI model** |  |  |  |  |  |  |
| Echo-AI Rest MBF, A.U. ± SD | 0 (0) | 0.8 ± 0.1 | 0.8 ± 0.1 | 0.8 ± 0.1 | 0.8 ± 0.1 | <0.001 |
| Echo-AI Stress MBF, A.U. ± SD | 0 (0) | 0.8 ± 0.3 | 1.0 ± 0.3 | 1.1 ± 0.3 | 1.4 ± 0.3 | <0.001 |
| Echo-AI CFR, A.U. ± SD | 0 (0) | 1.6 ± 0.1 | 1.8 ± 0.0 | 2.0 ± 0.1 | 2.3 ± 0.2 | <0.001 |
| **Demographics** |  |  |  |  |  |  |
| Age, years ± SD | 0 (0.0) | 72.8 ± 13.1 | 70.4 ± 13.7 | 65.8 ± 14.6 | 55.8 ± 16.7 | <0.001 |
| Gender, Female (%) | 0 (0.0) | 528 (39.9) | 527 (39.9) | 598 (45.2) | 655 (49.5) | <0.001 |
| Smoking | 0 (0) |  |  |  |  | <0.001 |
| Never, n (%) |  | 549 (41.5) | 554 (41.9) | 593 (44.9) | 677 (51.2) |  |
| Current, n (%) |  | 89 (6.7) | 89 (6.7) | 128 (9.7) | 141 (10.7) |  |
| Passive, n (%) |  | 0 (0.1) | 1 (0.1) | 0 (0.0) | 0 (0.0) |  |
| Quit, n (%) |  | 558 (42.2) | 541 (40.9) | 466 (35.2) | 398 (30.1) |  |
| Unknown, n (%) |  | 127 (9.6) | 137 (10.4) | 135 (10.2) | 106 (8.0) |  |
| Race | 0 (0) |  |  |  |  | 0.336 |
| White, n (%) |  | 1,150 (86.9) | 1,130 (85.5) | 1,114 (84.3) | 1,101 (83.3) |  |
| Black, n (%) |  | 61 (4.6) | 72 (5.4) | 70 (5.3) | 78 (5.9) |  |
| Hispanic, n (%) |  | 4 (0.3) | 12 (0.9) | 11 (0.8) | 9 (0.7) |  |
| Asian, n (%) |  | 37 (2.8) | 32 (2.4) | 37 (2.8) | 37 (2.8) |  |
| Others, n (%) |  | 41 (3.1) | 48 (3.6) | 60 (4.5) | 69 (5.2) |  |
| Unknown, n (%) |  | 30 (2.3) | 28 (2.1) | 30 (2.3) | 28 (2.1) |  |
| HR, bpm ± SD | 31 (0.6) | 73.2 ± 10.5 | 73.9 ± 12.0 | 74.9 ± 12.1 | 74.7 ± 12.4 | <0.001 |
| SBP, mmHg ± SD | 27 (0.5) | 128.4 ± 15.0 | 128.4 ± 15.0 | 127.9 ± 14.3 | 124.0 ± 13.1 | <0.001 |
| DBP, mmHg ± SD | 27 (0.5) | 68.6 ± 7.9 | 69.9 ± 7.9 | 70.9 ± 8.0 | 71.7 ± 8.2 | <0.001 |
| BMI, kg/m<sup>2</sup> ± SD | 82 (1.6) | 29.7 ± 7.6 | 29.1 ± 6.5 | 28.9 ± 8.0 | 27.2 ± 7.0 | <0.001 |
| eGFR, ml/min/1.73 m<sup>2</sup> ± SD | 374 (7.1) | 58.2 ± 27.3 | 64.0 ± 27.3 | 72.0 ± 27.8 | 85.3 ± 28.9 | <0.001 |
| EF, % ± SD | 421 (8.0) | 58.4 ± 16.4 | 61.6 ± 13.7 | 64.2 ± 11.0 | 65.7 ± 8.3 | <0.001 |
| **Disease history** |  |  |  |  |  |  |
| Heart failure, n (%) | 0 (0.0) | 702 (53.1) | 567 (42.9) | 383 (29.0) | 151 (11.4) | <0.001 |
| Type 2 DM, n (%) | 0 (0.0) | 479 (36.2) | 367 (27.8) | 325 (24.6) | 181 (13.7) | <0.001 |
| Hypertension, n (%) | 0 (0.0) | 1,129 (85.3) | 1,088 (82.3) | 1,021 (77.2) | 737 (55.7) | <0.001 |
| Atrial fibrillation, n (%) | 0 (0.0) | 666 (50.3) | 587 (44.2) | 446 (33.7) | 219 (16.6) | <0.001 |
| Hyperlipidemia, n (%) | 0 (0.0) | 960 (72.6) | 907 (68.6) | 817 (61.8) | 632 (47.8) | <0.001 |
| CAD, n (%) | 0 (0.0) | 745 (56.3) | 592 (44.8) | 420 (31.8) | 273 (20.7) | <0.001 |
| Stroke, n (%) | 0 (0.0) | 207 (15.6) | 132 (10.0) | 156 (11.8) | 96 (7.3) | <0.001 |
| PAD, n (%) | 0 (0.0) | 337 (25.5) | 283 (21.4) | 275 (20.8) | 204 (15.4) | <0.001 |
| COPD, n (%) | 0 (0.0) | 224 (16.9) | 187 (14.1) | 177 (13.4) | 104 (7.9) | <0.001 |
| Aortic stenosis | 0 (0.0) | 205 (15.5) | 150 (11.3) | 125 (9.5) | 67 (5.1) | <0.001 |
| Aortic regurgitation | 0 (0.0) | 43 (3.3) | 39 (3.0) | 47 (3.6) | 31 (2.3) | 0.307 |
| Mitral stenosis | 0 (0.0) | 18 (1.4) | 18 (1.4) | 10 (0.8) | 7 (0.5) | 0.065 |
| Mitral regurgitation | 0 (0.0) | 53 (4.0) | 50 (3.8) | 39 (3.0) | 32 (2.4) | 0.080 |
| **Medication** |  |  |  |  |  |  |
| ASA, n (%) | 0 (0.0) | 969 (73.2) | 857 (64.8) | 817 (61.8) | 606 (45.8) | <0.001 |
| Statin | 0 (0.0) | 971 (73.4) | 873 (66.0) | 783 (59.2) | 480 (36.3) | <0.001 |
| ACEi, n (%) | 0 (0.0) | 472 (35.7) | 452 (34.2) | 385 (29.1) | 283 (21.4) | <0.001 |
| ARB, n (%) | 0 (0.0) | 275 (20.8) | 249 (18.8) | 223 (16.9) | 153 (11.6) | <0.001 |
| Beta blocker, n (%) | 0 (0.0) | 990 (74.8) | 896 (67.8) | 761 (57.6) | 485 (36.7) | <0.001 |
| Thiazide, n (%) | 0 (0.0) | 225 (17.0) | 233 (17.6) | 183 (13.8) | 135 (10.2) | <0.001 |
| Loop diuretic, n (%) | 0 (0.0) | 692 (52.3) | 534 (40.4) | 403 (30.5) | 175 (13.2) | <0.001 |
| MRA, n (%) | 0 (0.0) | 156 (11.8) | 105 (7.9) | 96 (7.3) | 42 (3.2) | <0.001 |

Abbreviations. MGH: Massachusetts General Hospital, CFR: coronary flow reserve, MBF: myocardial blood flow, A.U.: arbitrary unit, HR: heart rate, SBP: systolic blood pressure, DBP: diastolic blood pressure, BMI: body mass index, eGFR: estimated glomerular filtration rate, EF: ejection fraction, LV: left ventricle, DM: diabetes mellitus, CAD: coronary heart disease, PAD: peripheral artery disease, COPD: chronic obstructive pulmonary disease, ASA: acetyl-salicylic acid, ACEi: angiotensin-converting enzyme inhibitor, ARB: angiotensin II receptor blocker, MRA: mineralocorticoid receptor antagonist

**Table S4.** Baseline demographic for EchoAI-restMBF quartiles for heart failure survival analysis (BWH cohort)

|  | Missing n (%) | Quartile 1 | Quartile 2 | Quartile 3 | Quartile 4 | p |
| --- | --- | --- | --- | --- | --- | --- |
| Number patients |  | 1322 | 1322 | 1322 | 1322 |  |
| **AI model** |  |  |  |  |  |  |
| Echo-AI Rest MBF, A.U. ± SD | 0 (0) | 0.7 ± 0.0 | 0.8 ± 0.0 | 0.9 ± 0.0 | 1.0 ± 0.1 | <0.001 |
| Echo-AI Stress MBF, A.U. ± SD | 0 (0) | 0.9 ± 0.4 | 1.2 ± 0.4 | 1.4 ± 0.4 | 1.6 ± 0.4 | <0.001 |
| Echo-AI CFR, A.U. ± SD | 0 (0) | 1.9 ± 0.4 | 2.0 ± 0.4 | 2.0 ± 0.4 | 2.0 ± 0.4 | <0.001 |
| **Demographics** |  |  |  |  |  |  |
| Age, years ± SD | 11 (0.2) | 61.9 ± 16.3 | 59.9 ± 16.5 | 60.6 ± 16.0 | 58.1 ± 16.3 | <0.001 |
| Gender, Female (%) | 11 (0.2) | 401(30.4) | 668 (50.6) | 792 (60.0) | 948 (71.8) | <0.001 |
| Smoking | 0 (0) |  |  |  |  | <0.001 |
| Never, n (%) |  | 611 (46.2) | 674 (51.0) | 703 (53.2) | 731 (55.3) |  |
| Current, n (%) |  | 77 (5.8) | 80 (6.1) | 81 (6.1) | 75 (5.7) |  |
| Passive, n (%) |  | 1 (0.1) | 0 (0.0) | 3 (0.2) | 6 (0.5) |  |
| Quit, n (%) |  | 501 (37.9) | 437 (33.1) | 437 (33.1) | 408 (30.9) |  |
| Unknown, n (%) |  | 132 (10.0) | 131 (9.9) | 98 (7.4) | 102 (7.7) |  |
| Race | 0 (0) |  |  |  |  | 0.186 |
| White, n (%) |  | 1080 (81.7) | 1066 (80.6) | 1079 (81.6) | 1041 (78.7) |  |
| Black, n (%) |  | 104 (7.9) | 89 (6.7) | 92 (7.0) | 109 (8.2) |  |
| Hispanic, n (%) |  | 41 (3.1) | 33 (2.5) | 31 (2.3) | 42 (3.2) |  |
| Asian, n (%) |  | 22 (1.7) | 39 (3.0) | 27 (2.0) | 38 (2.9) |  |
| Others, n (%) |  | 28 (2.1) | 38 (2.9) | 46 (3.5) | 45 (3.4) |  |
| Unknown, n (%) |  | 47 (3.6) | 57 (4.3) | 47 (3.6) | 47 (3.6) |  |
| HR, bpm ± SD | 369 (7.0) | 73.9 ± 13.8 | 73.3 ± 12.9 | 75.2 ± 12.0 | 78.6 ± 11.8 | <0.001 |
| SBP, mmHg ± SD | 346 (6.5) | 127.1 ± 16.6 | 128.5 ± 14.9 | 128.8 ± 15.3 | 129.1 ± 15.3 | <0.008 |
| DBP, mmHg ± SD | 346 (6.5) | 73.9 ± 11.6 | 73.6 ± 9.9 | 73.7 ± 9.7 | 73.6 ± 9.6 | <0.860 |
| BMI, kg/m<sup>2</sup> ± SD | 587 (11.1) | 29.1 ± 7.7 | 28.6 ± 7.3 | 28.3 ± 6.9 | 27.6 ± 8.7 | <0.001 |
| eGFR, ml/min/1.73 m<sup>2</sup> ± SD | 561 (10.6) | 70.5 ± 27.2 | 79.5 ± 28.8 | 80.4 ± 28.0 | 87.9 ± 29.0 | <0.001 |
| EF, % ± SD | 400 (7.6) | 53.2 ± 13.4 | 59.6 ± 8.1 | 61.5 ± 6.5 | 62.7 ± 5.6 | <0.001 |
| **Disease history** |  |  |  |  |  |  |
| Heart failure, n (%) | 3 (0.0) | 413 (31.3) | 224 (17.0) | 130 (9.8) | 128 (9.7) | <0.001 |
| Type 2 DM, n (%) | 3 (0.0) | 261 (19.8) | 209 (15.8) | 216 (16.3) | 186 (14.1) | <0.001 |
| Hypertension, n (%) | 3 (0.0) | 709 (53.7) | 702 (53.1) | 713 (53.9) | 651 (49.3) | <0.001 |
| Atrial fibrillation, n (%) | 3 (0.0) | 452 (34.2) | 205 (15.5) | 118 (8.9) | 86 (6.5) | <0.001 |
| Hyperlipidemia, n (%) | 3 (0.0) | 489 (37.0) | 479 (36.3) | 487 (36.8) | 402 (30.4) | <0.001 |
| CAD, n (%) | 3 (0.0) | 345 (26.1) | 269 (20.4) | 181 (13.7) | 132 (10.0) | <0.001 |
| Stroke, n (%) | 3 (0.0) | 108 (8.2) | 88 (6.7) | 76 (5.7) | 69 (5.2) | <0.001 |
| PAD, n (%) | 3 (0.0) | 128 (9.7) | 123 (9.3) | 117 (8.9) | 97 (7.3) | <0.001 |
| COPD, n (%) | 3 (0.0) | 74 (5.6) | 85 (6.4) | 94 (7.1) | 77 (5.8) | <0.001 |
| Aortic stenosis | 3 (0.0) | 73 (5.5) | 69 (5.2) | 74 (5.6) | 55 (4.2) | 0.310 |
| Aortic regurgitation | 3 (0.0) | 23 (1.7) | 31 (2.3) | 27 (2.0) | 18 (1.4) | 0.282 |
| Mitral stenosis | 3 (0.0) | 13 (1.0) | 5 (0.4) | 5 (0.4) | 4 (0.3) | 0.049 |
| Mitral regurgitation | 3 (0.0) | 98 (7.4) | 67 (5.1) | 60 (4.5) | 48 (3.6) | <0.001 |
| **Medication** |  |  |  |  |  |  |
| ASA, n (%) | 3 (0.0) | 663 (50.2) | 617 (46.7) | 556 (42.1) | 495 (37.5) | <0.001 |
| Statin | 3 (0.0) | 680 (51.5) | 584 (44.2) | 564 (42.7) | 436 (33.0) | <0.001 |
| ACEi, n (%) | 3 (0.0) | 439 (33.2) | 344 (26.0) | 322 (24.4) | 286 (21.7) | <0.001 |
| ARB, n (%) | 3 (0.0) | 216 (16.4) | 185 (14.0) | 197 (14.9) | 172 (13.0) | <0.095 |
| Beta blocker, n (%) | 3 (0.0) | 768 (58.1) | 604 (45.7) | 539 (40.8) | 421 (31.9) | <0.001 |
| Thiazide, n (%) | 3 (0.0) | 175 (13.2) | 149 (11.3) | 165 (12.5) | 159 (12.0) | <0.475 |
| Loop diuretic, n (%) | 3 (0.0) | 468 (35.4) | 265 (20.1) | 194 (14.7) | 153 (11.6) | <0.001 |
| MRA, n (%) | 3 (0.0) | 150 (11.4) | 83 (6.3) | 50 (3.8) | 45 (3.4) | <0.001 |

Abbreviations. BWH: Brigham and Women's Hospital, CFR: coronary flow reserve, MBF: myocardial blood flow, A.U.: arbitrary unit, HR: heart rate, SBP: systolic blood pressure, DBP: diastolic blood pressure, BMI: body mass index, eGFR: estimated glomerular filtration rate, EF: ejection fraction, LV: left ventricle, DM: diabetes mellitus, CAD: coronary heart disease, PAD: peripheral artery disease, COPD: chronic obstructive pulmonary disease, ASA: acetyl-salicylic acid, ACEi: angiotensin-converting enzyme inhibitor, ARB: angiotensin II receptor blocker, MRA: mineralocorticoid receptor antagonist

**Table S5.** Baseline demographic for EchoAI-stressMBF quartiles for heart failure survival analysis (BWH cohort)

|  | Missing n (%) | Quartile 1 | Quartile 2 | Quartile 3 | Quartile 4 | p |
| --- | --- | --- | --- | --- | --- | --- |
| Number patients |  | 1322 | 1322 | 1322 | 1322 |  |
| **AI model** |  |  |  |  |  |  |
| Echo-AI Rest MBF, A.U. ± SD | 0 (0) | 0.7 ± 0.1 | 0.8 ± 0.1 | 0.9 ± 0.1 | 0.9 ± 0.1 | <0.001 |
| Echo-AI Stress MBF, A.U. ± SD | 0 (0) | 0.7 ± 0.2 | 1.1 ± 0.1 | 1.4 ± 0.1 | 1.9 ± 0.2 | <0.001 |
| Echo-AI CFR, A.U. ± SD | 0 (0) | 1.7 ± 0.2 | 1.9 ± 0.3 | 2.1 ± 0.3 | 2.3 ± 0.4 | <0.001 |
| **Demographics** |  |  |  |  |  |  |
| Age, years ± SD | 11 (0.2) | 68.2 ± 14.0 | 62.9 ± 15.0 | 58.7 ± 15.0 | 50.8 ± 16.0 | <0.001 |
| Gender, Female (%) | 11 (0.2) | 412(31.3) | 618 (46.9) | 771 (58.3) | 1008(76.5) | <0.001 |
| Smoking | 0 (0) |  |  |  |  | <0.001 |
| Never, n (%) |  | 565(42.7) | 655 (49.5) | 695 (52.6) | 804(60.8) |  |
| Current, n (%) |  | 62 (4.7) | 83 (6.3) | 85 (6.4) | 83 (6.3) |  |
| Passive, n (%) |  | 2 (0.2) | 1 (0.1) | 3 (0.2) | 4(0.3) |  |
| Quit, n (%) |  | 572 (43.3) | 465 (35.2) | 437 (33.1) | 309 (23.4) |  |
| Unknown, n (%) |  | 121 (9.2) | 118 (8.9) | 102 (7.7) | 122 (9.2) |  |
| Race | 0 (0) |  |  |  |  | 0.007 |
| White, n (%) |  | 1080 (81.7) | 1074 (81.2) | 1081 (81.8) | 1031 (78.0) |  |
| Black, n (%) |  | 108 (8.2) | 97 (7.3) | 90(6.8) | 99 (7.5) |  |
| Hispanic, n (%) |  | 23 (1.7) | 40 (3.0) | 41(3.1) | 43 (3.3) |  |
| Asian, n (%) |  | 23 (1.7) | 28 (2.1) | 25 (1.9) | 50 (3.8) |  |
| Others, n (%) |  | 34 (2.6) | 32 (2.4) | 40 (3.0) | 51 (3.9) |  |
| Unknown, n (%) |  | 54 (4.1) | 51 (3.9) | 45 (3.4) | 48 (3.6) |  |
| HR, bpm ± SD | 369 (7.0) | 72.4 ± 13.2 | 73.3 ± 12.9 | 76.1 ± 12.0 | 79.4 ± 11.9 | <0.001 |
| SBP, mmHg ± SD | 346 (6.5) | 129.2 ± 17.1 | 130.3 ± 15.4 | 128.8 ± 14.4 | 125.4 ± 14.9 | <0.001 |
| DBP, mmHg ± SD | 346 (6.5) | 72.4 ± 11.7 | 74.0 ± 10.1 | 74.5 ± 9.6 | 73.9 ± 9.4 | <0.001 |
| BMI, kg/m<sup>2</sup> ± SD | 587 (11.1) | 29.7 ± 8.3 | 29.3 ± 8.9 | 28.2 ± 6.3 | 26.4 ± 6.6 | <0.001 |
| eGFR, ml/min/1.73 m<sup>2</sup> ± SD | 561 (10.6) | 64.1 ± 25.4 | 75.4 ± 26.5 | 83.4 ± 27.1 | 95.8 ± 27.0 | <0.001 |
| EF, % ± SD | 400 (7.6) | 52.6 ± 13.9 | 60.1 ± 7.7 | 61.7 ± 5.6 | 62.5 ± 5.2 | <0.001 |
| **Disease history** |  |  |  |  |  |  |
| Heart failure, n (%) | 3 (0.0) | 492(37.2) | 211 (16.0) | 137 (10.4) | 55 (4.2) | <0.001 |
| Type 2 DM, n (%) | 3 (0.0) | 337 (25.5) | 256 (19.4) | 174 (13.2) | 105 (7.9) | <0.001 |
| Hypertension, n (%) | 3 (0.0) | 857 (64.9) | 796 (60.3) | 662 (50.1) | 460 (34.8) | <0.001 |
| Atrial fibrillation, n (%) | 3 (0.0) | 511 (38.7) | 220 (16.7) | 77 (5.8) | 53(4.0) | <0.001 |
| Hyperlipidemia, n (%) | 3 (0.0) | 577 (43.7) | 539 (40.8) | 441 (33.4) | 300 (22.7) | <0.001 |
| CAD, n (%) | 3 (0.0) | 478 (36.2) | 246 (18.6) | 153 (11.6) | 50 (3.8) | <0.001 |
| Stroke, n (%) | 3 (0.0) | 129 (9.8) | 88 (6.7) | 81(6.1) | 43(3.3) | <0.001 |
| PAD, n (%) | 3 (0.0) | 159 (12.0) | 134 (10.2) | 116 (8.8) | 56 (4.2) | <0.001 |
| COPD, n (%) | 3 (0.0) | 104 (7.9) | 96 (7.3) | 75 (5.7) | 55 (4.2) | <0.001 |
| Aortic stenosis | 3 (0.0) | 123 (9.3) | 97 (7.3) | 36 (2.7) | 15 (1.1) | <0.001 |
| Aortic regurgitation | 3 (0.0) | 31 (2.3) | 30 (2.3) | 25 (1.9) | 12 (1.0) | 0.037 |
| Mitral stenosis | 3 (0.0) | 15 (1.1) | 9 (0.7) | 3 (0.2) | 0 (0.0) | <0.001 |
| Mitral regurgitation | 3 (0.0) | 129 (9.8) | 85 (6.4) | 37 (2.8) | 22 (1.7) | <0.001 |
| **Medication** |  |  |  |  |  |  |
| ASA, n (%) | 3 (0.0) | 801 (60.6) | 674 (51.1) | 528 (39.9) | 328(24.8) | <0.001 |
| Statin | 3 (0.0) | 828 (62.7) | 663 (50.2) | 499 (37.7) | 274 (20.7) | <0.001 |
| ACEi, n (%) | 3 (0.0) | 488 (36.9) | 367 (27.8) | 318 (24.1) | 218 (16.5) | <0.001 |
| ARB, n (%) | 3 (0.0) | 269 (20.4) | 222 (16.8) | 178 (13.5) | 101 (7.6) | <0.001 |
| Beta blocker, n (%) | 3 (0.0) | 936 (70.9) | 693 (52.5) | 460 (34.8) | 243 (18.4) | <0.001 |
| Thiazide, n (%) | 3 (0.0) | 195 (14.8) | 173 (13.1) | 169 (12.8) | 111 (8.4) | <0.001 |
| Loop diuretic, n (%) | 3 (0.0) | 601 (45.5) | 242 (18.3) | 153 (11.6) | 84 (6.4) | <0.001 |
| MRA, n (%) | 3 (0.0) | 181 (13.7) | 74 (5.6) | 38 (2.9) | 35 (2.6) | <0.001 |

Abbreviations. BWH: Brigham and Women's Hospital, CFR: coronary flow reserve, MBF: myocardial blood flow, A.U.: arbitrary unit, HR: heart rate, SBP: systolic blood pressure, DBP: diastolic blood pressure, BMI: body mass index, eGFR: estimated glomerular filtration rate, EF: ejection fraction, LV: left ventricle, DM: diabetes mellitus, CAD: coronary heart disease, PAD: peripheral artery disease, COPD: chronic obstructive pulmonary disease, ASA: acetyl-salicylic acid, ACEi: angiotensin-converting enzyme inhibitor, ARB: angiotensin II receptor blocker, MRA: mineralocorticoid receptor antagonist

**Table S6.** Baseline demographic for EchoAI-CFR quartiles for heart failure survival analysis (BWH cohort)

|  | Missing n (%) | Quartile 1 | Quartile 2 | Quartile 3 | Quartile 4 | p |
| --- | --- | --- | --- | --- | --- | --- |
| Number patients |  | 1322 | 1322 | 1322 | 1322 |  |
| **AI model** |  |  |  |  |  |  |
| Echo-AI Rest MBF, A.U. ± SD | 0 (0) | 0.8 ± 0.1 | 0.8 ± 0.1 | 0.9 ± 0.1 | 0.9 ± 0.1 | <0.001 |
| Echo-AI Stress MBF, A.U. ± SD | 0 (0) | 0.9 ± 0.4 | 1.2 ± 0.4 | 1.4 ± 0.4 | 1.7 ± 0.4 | <0.001 |
| Echo-AI CFR, A.U. ± SD | 0 (0) | 1.6 ± 0.1 | 1.8 ± 0.1 | 2.1 ± 0.1 | 2.5 ± 0.3 | <0.001 |
| **Demographics** |  |  |  |  |  |  |
| Age, years ± SD | 11 (0.2) | 69.0 ± 14.0 | 64.2 ± 14.8 | 58.2 ± 14.5 | 49.1 ± 14.8 | <0.001 |
| Gender, Female (%) | 11 (0.2) | 582 (44.1) | 618 (46.9) | 725 (55.0) | 884 (67.0) | <0.001 |
| Smoking | 0 (0) |  |  |  |  | <0.001 |
| Never, n (%) |  | 602 (45.5) | 633 (47.9) | 694 (52.5) | 790 (59.8) |  |
| Current, n (%) |  | 60 (4.5) | 85 (6.4) | 87 (6.6) | 81 (6.1) |  |
| Passive, n (%) |  | 4(0.3) | 1 (0.1) | 3 (0.2) | 2 (0.2) |  |
| Quit, n (%) |  | 547 (41.4) | 490 (37.1) | 440 (33.3) | 306 (23.1) |  |
| Unknown, n (%) |  | 109 (8.2) | 113 (8.5) | 98 (7.4) | 143 (10.8) |  |
| Race | 0 (0) |  |  |  |  | 0.112 |
| White, n (%) |  | 1068 (80.8) | 1078 (81.5) | 1035 (78.3) | 1085 (82.1) |  |
| Black, n (%) |  | 100 (7.6) | 101 (7.6) | 114 (8.6) | 79 (6.0) |  |
| Hispanic, n (%) |  | 38 (2.9) | 31 (2.3) | 44 (3.3) | 34 (2.6) |  |
| Asian, n (%) |  | 28 (2.1) | 33 (2.5) | 25 (1.9) | 40 (3.0) |  |
| Others, n (%) |  | 37 (2.8) | 35 (2.6) | 41 (3.1) | 44 (3.3) |  |
| Unknown, n (%) |  | 51 (3.9) | 44 (3.3) | 63 (4.8) | 40 (3.0) |  |
| HR, bpm ± SD | 369 (7.0) | 73.8 ± 12.7 | 75.1 ± 13.2 | 76.0 ± 12.9 | 76.3 ± 12.3 | <0.001 |
| SBP, mmHg ± SD | 346 (6.5) | 130.2 ± 17.1 | 130.5 ± 15.6 | 128.4 ± 15.2 | 124.5 ± 13.4 | <0.001 |
| DBP, mmHg ± SD | 346 (6.5) | 72.2 ± 10.9 | 73.8 ± 10.9 | 74.6 ± 9.9 | 74.2 ± 9.0 | <0.001 |
| BMI, kg/m<sup>2</sup> ± SD | 587 (11.1) | 29.1 ± 9.8 | 29.2 ± 7.6 | 28.6 ± 6.2 | 26.6 ± 6.4 | <0.001 |
| eGFR, ml/min/1.73 m<sup>2</sup> ± SD | 561 (10.6) | 66.9 ± 27.2 | 74.9 ± 28.2 | 82.6 ± 27.1 | 94.9 ± 25.7 | <0.001 |
| EF, % ± SD | 400 (7.6) | 55.9 ± 13.2 | 59.0 ± 9.9 | 60.6 ± 7.3 | 61.6 ± 5.2 | <0.001 |
| **Disease history** |  |  |  |  |  |  |
| Heart failure, n (%) | 3 (0.0) | 410 (31.0) | 259 (19.6) | 156 (11.8) | 70 (5.3) | <0.001 |
| Type 2 DM, n (%) | 3 (0.0) | 351 (26.6) | 257 (19.5) | 192 (14.5) | 72 (5.4) | <0.001 |
| Hypertension, n (%) | 3 (0.0) | 886 (67.1) | 813 (61.6) | 675 (51.1) | 401 (30.3) | <0.001 |
| Atrial fibrillation, n (%) | 3 (0.0) | 373 (28.2) | 280 (21.2) | 139 (10.5) | 69 (5.2) | <0.001 |
| Hyperlipidemia, n (%) | 3 (0.0) | 581 (44.0) | 548 (41.5) | 449 (34.0) | 279 (21.1) | <0.001 |
| CAD, n (%) | 3 (0.0) | 429 (32.5) | 286 (21.7) | 153 (11.6) | 59 (4.5) | <0.001 |
| Stroke, n (%) | 3 (0.0) | 149 (11.3) | 88 (6.7) | 65 (4.9) | 39 (3.0) | <0.001 |
| PAD, n (%) | 3 (0.0) | 184 (13.9) | 131 (9.9) | 82 (6.2) | 68 (5.1) | <0.001 |
| COPD, n (%) | 3 (0.0) | 115 (8.7) | 105 (8.0) | 65 (4.9) | 45 (3.4) | <0.001 |
| Aortic stenosis | 3 (0.0) | 143 (10.8) | 80 (6.1) | 39 (3.0) | 9 (0.7) | <0.001 |
| Aortic regurgitation | 3 (0.0) | 26 (2.0) | 36 (2.7) | 19 (1.4) | 18 (1.4) | 0.036 |
| Mitral stenosis | 3 (0.0) | 16 (1.2) | 7 (0.5) | 4 (0.3) | 0 (0.0) | <0.001 |
| Mitral regurgitation | 3 (0.0) | 120 (9.1) | 88 (6.7) | 38 (2.9) | 27 (2.0) | <0.001 |
| **Medication** |  |  |  |  |  |  |
| ASA, n (%) | 3 (0.0) | 803 (60.8) | 683 (51.7) | 510 (25.4) | 132 (13.3) | <0.001 |
| Statin | 3 (0.0) | 797 (60.3) | 693 (52.5) | 511 (38.7) | 263 (19.9) | <0.001 |
| ACEi, n (%) | 3 (0.0) | 448 (33.9) | 411 (31.1) | 332 (25.1) | 200 (15.1) | <0.001 |
| ARB, n (%) | 3 (0.0) | 272 (20.6) | 218 (16.5) | 183 (13.8) | 97 (7.3) | <0.001 |
| Beta blocker, n (%) | 3 (0.0) | 861 (65.2) | 722 (54.7) | 469 (35.5) | 280 (21.2) | <0.001 |
| Thiazide, n (%) | 3 (0.0) | 189 (14.3) | 196 (14.8) | 168 (12.7) | 95 (7.2) | <0.001 |
| Loop diuretic, n (%) | 3 (0.0) | 494 (37.4) | 343 (26.0) | 182 (13.8) | 61 (4.6) | <0.001 |
| MRA, n (%) | 3 (0.0) | 146 (11.1) | 105 (8.0) | 50 (3.8) | 27 (2.0) | <0.001 |

Abbreviations. BWH: Brigham and Women's Hospital, CFR: coronary flow reserve, MBF: myocardial blood flow, A.U.: arbitrary unit, HR: heart rate, SBP: systolic blood pressure, DBP: diastolic blood pressure, BMI: body mass index, eGFR: estimated glomerular filtration rate, EF: ejection fraction, LV: left ventricle, DM: diabetes mellitus, CAD: coronary heart disease, PAD: peripheral artery disease, COPD: chronic obstructive pulmonary disease, ASA: acetyl-salicylic acid, ACEi: angiotensin-converting enzyme inhibitor, ARB: angiotensin II receptor blocker, MRA: mineralocorticoid receptor antagonist

**Table S7.**
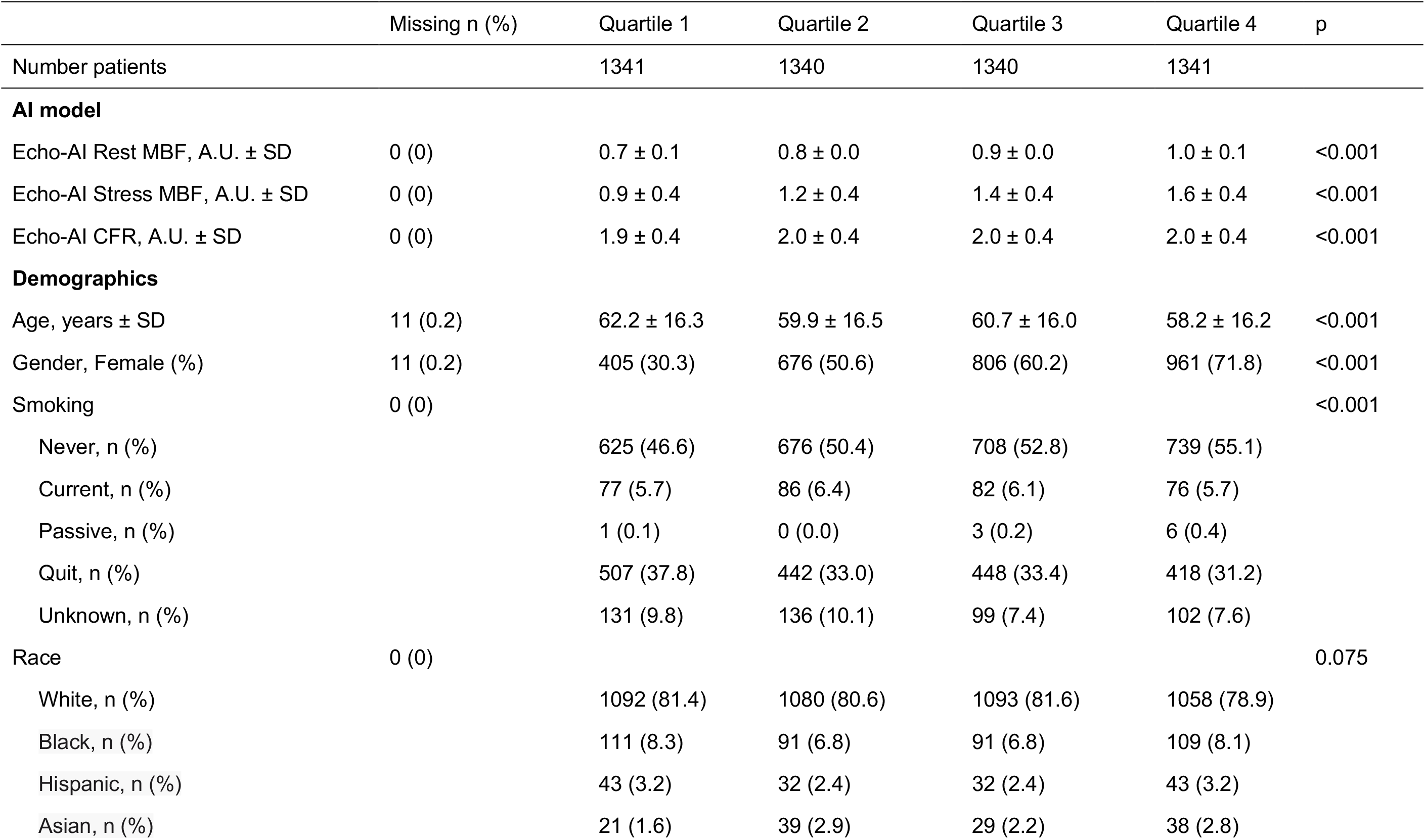
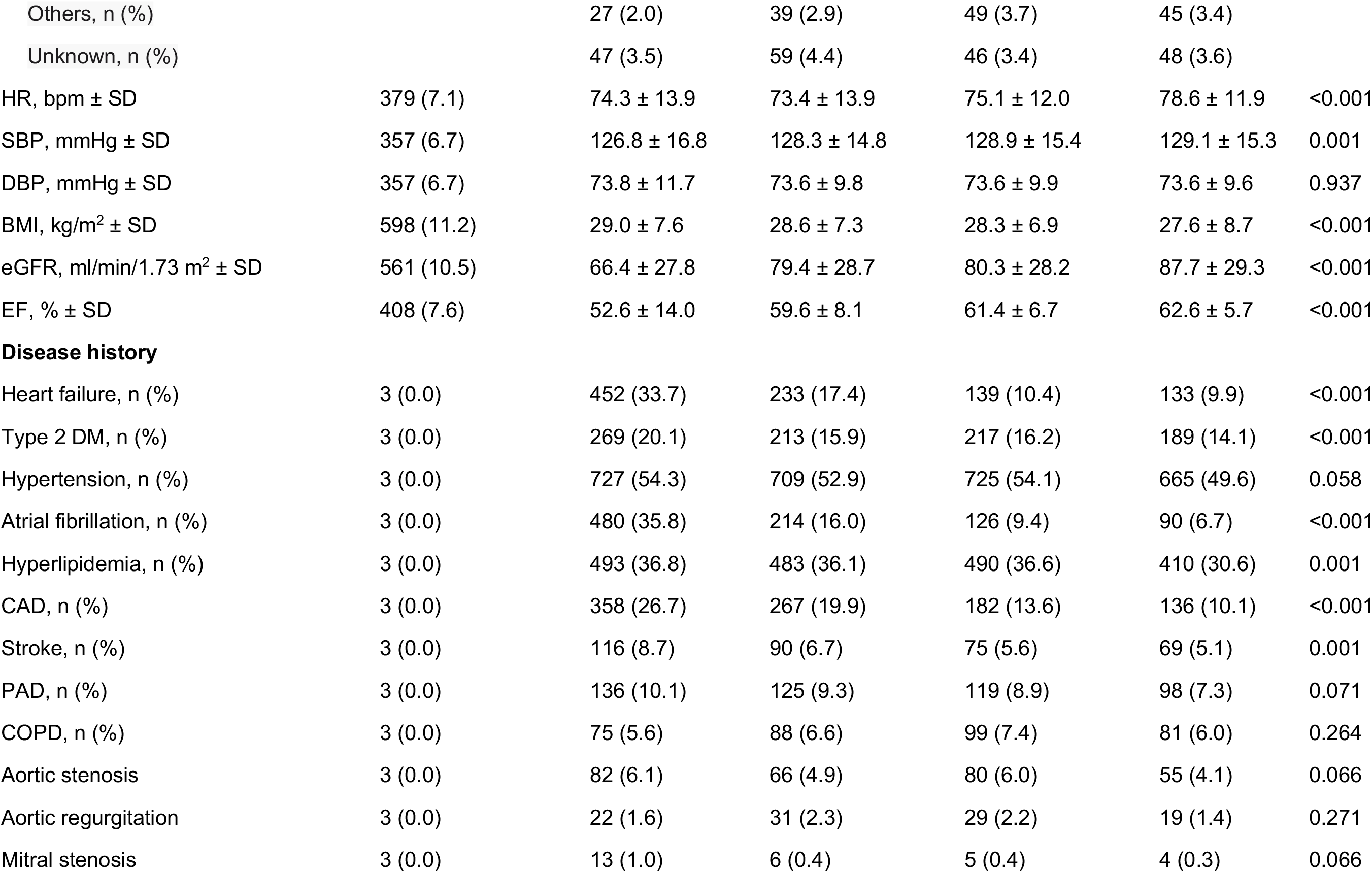
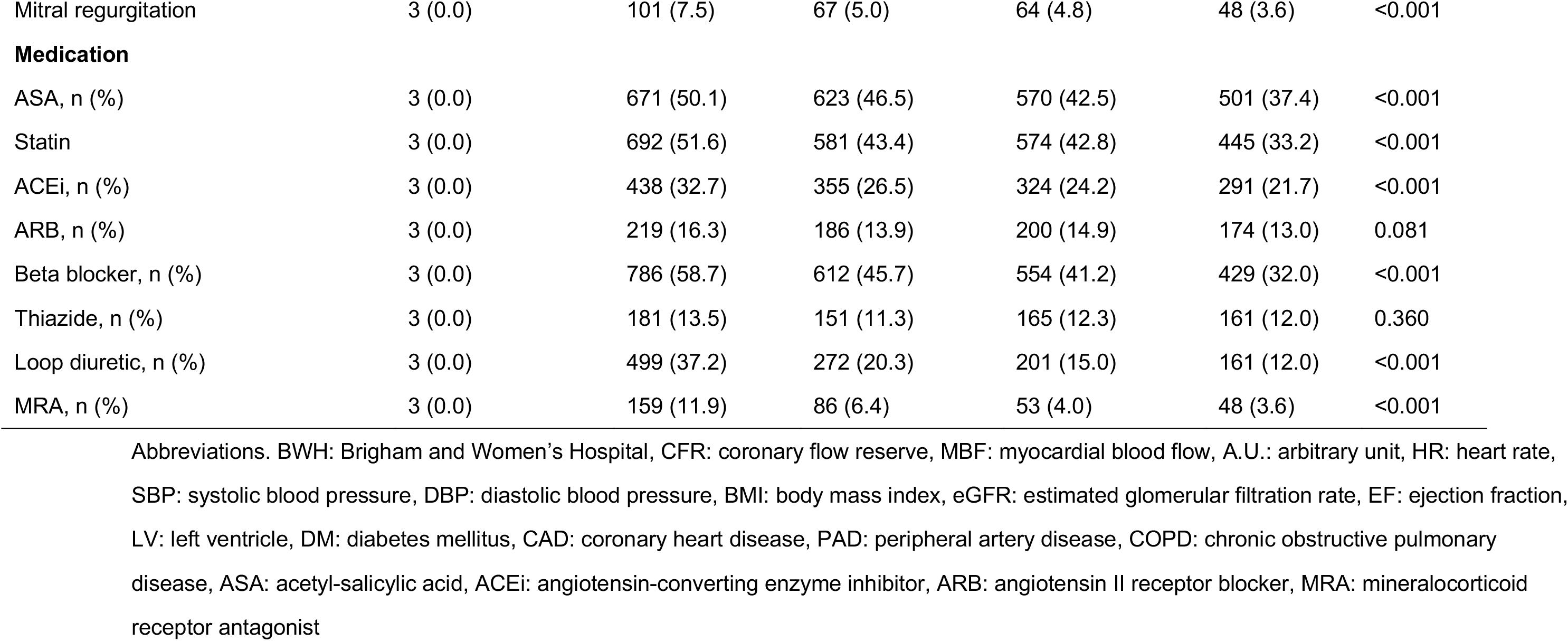
Baseline demographic for EchoAI-restMBF quartiles for ACS survival analysis (BWH cohort)

|  | Missing n (%) | Quartile 1 | Quartile 2 | Quartile 3 | Quartile 4 | p |
| --- | --- | --- | --- | --- | --- | --- |
| Number patients |  | 1341 | 1340 | 1340 | 1341 |  |
| **AI model** |  |  |  |  |  |  |
| Echo-AI Rest MBF, A.U. ± SD | 0 (0) | 0.7 ± 0.1 | 0.8 ± 0.0 | 0.9 ± 0.0 | 1.0 ± 0.1 | <0.001 |
| Echo-AI Stress MBF, A.U. ± SD | 0 (0) | 0.9 ± 0.4 | 1.2 ± 0.4 | 1.4 ± 0.4 | 1.6 ± 0.4 | <0.001 |
| Echo-AI CFR, A.U. ± SD | 0 (0) | 1.9 ± 0.4 | 2.0 ± 0.4 | 2.0 ± 0.4 | 2.0 ± 0.4 | <0.001 |
| **Demographics** |  |  |  |  |  |  |
| Age, years ± SD | 11 (0.2) | 62.2 ± 16.3 | 59.9 ± 16.5 | 60.7 ± 16.0 | 58.2 ± 16.2 | <0.001 |
| Gender, Female (%) | 11 (0.2) | 405 (30.3) | 676 (50.6) | 806 (60.2) | 961 (71.8) | <0.001 |
| Smoking | 0 (0) |  |  |  |  | <0.001 |
| Never, n (%) |  | 625 (46.6) | 676 (50.4) | 708 (52.8) | 739 (55.1) |  |
| Current, n (%) |  | 77 (5.7) | 86 (6.4) | 82 (6.1) | 76 (5.7) |  |
| Passive, n (%) |  | 1 (0.1) | 0 (0.0) | 3 (0.2) | 6 (0.4) |  |
| Quit, n (%) |  | 507 (37.8) | 442 (33.0) | 448 (33.4) | 418 (31.2) |  |
| Unknown, n (%) |  | 131 (9.8) | 136 (10.1) | 99 (7.4) | 102 (7.6) |  |
| Race | 0 (0) |  |  |  |  | 0.075 |
| White, n (%) |  | 1092 (81.4) | 1080 (80.6) | 1093 (81.6) | 1058 (78.9) |  |
| Black, n (%) |  | 111 (8.3) | 91 (6.8) | 91 (6.8) | 109 (8.1) |  |
| Hispanic, n (%) |  | 43 (3.2) | 32 (2.4) | 32 (2.4) | 43 (3.2) |  |
| Asian, n (%) |  | 21 (1.6) | 39 (2.9) | 29 (2.2) | 38 (2.8) |  |
| Others, n (%) |  | 27 (2.0) | 39 (2.9) | 49 (3.7) | 45 (3.4) |  |
| Unknown, n (%) |  | 47 (3.5) | 59 (4.4) | 46 (3.4) | 48 (3.6) |  |
| HR, bpm ± SD | 379 (7.1) | 74.3 ± 13.9 | 73.4 ± 13.9 | 75.1 ± 12.0 | 78.6 ± 11.9 | <0.001 |
| SBP, mmHg ± SD | 357 (6.7) | 126.8 ± 16.8 | 128.3 ± 14.8 | 128.9 ± 15.4 | 129.1 ± 15.3 | 0.001 |
| DBP, mmHg ± SD | 357 (6.7) | 73.8 ± 11.7 | 73.6 ± 9.8 | 73.6 ± 9.9 | 73.6 ± 9.6 | 0.937 |
| BMI, kg/m<sup>2</sup> ± SD | 598 (11.2) | 29.0 ± 7.6 | 28.6 ± 7.3 | 28.3 ± 6.9 | 27.6 ± 8.7 | <0.001 |
| eGFR, ml/min/1.73 m<sup>2</sup> ± SD | 561 (10.5) | 66.4 ± 27.8 | 79.4 ± 28.7 | 80.3 ± 28.2 | 87.7 ± 29.3 | <0.001 |
| EF, % ± SD | 408 (7.6) | 52.6 ± 14.0 | 59.6 ± 8.1 | 61.4 ± 6.7 | 62.6 ± 5.7 | <0.001 |
| **Disease history** |  |  |  |  |  |  |
| Heart failure, n (%) | 3 (0.0) | 452 (33.7) | 233 (17.4) | 139 (10.4) | 133 (9.9) | <0.001 |
| Type 2 DM, n (%) | 3 (0.0) | 269 (20.1) | 213 (15.9) | 217 (16.2) | 189 (14.1) | <0.001 |
| Hypertension, n (%) | 3 (0.0) | 727 (54.3) | 709 (52.9) | 725 (54.1) | 665 (49.6) | 0.058 |
| Atrial fibrillation, n (%) | 3 (0.0) | 480 (35.8) | 214 (16.0) | 126 (9.4) | 90 (6.7) | <0.001 |
| Hyperlipidemia, n (%) | 3 (0.0) | 493 (36.8) | 483 (36.1) | 490 (36.6) | 410 (30.6) | 0.001 |
| CAD, n (%) | 3 (0.0) | 358 (26.7) | 267 (19.9) | 182 (13.6) | 136 (10.1) | <0.001 |
| Stroke, n (%) | 3 (0.0) | 116 (8.7) | 90 (6.7) | 75 (5.6) | 69 (5.1) | 0.001 |
| PAD, n (%) | 3 (0.0) | 136 (10.1) | 125 (9.3) | 119 (8.9) | 98 (7.3) | 0.071 |
| COPD, n (%) | 3 (0.0) | 75 (5.6) | 88 (6.6) | 99 (7.4) | 81 (6.0) | 0.264 |
| Aortic stenosis | 3 (0.0) | 82 (6.1) | 66 (4.9) | 80 (6.0) | 55 (4.1) | 0.066 |
| Aortic regurgitation | 3 (0.0) | 22 (1.6) | 31 (2.3) | 29 (2.2) | 19 (1.4) | 0.271 |
| Mitral stenosis | 3 (0.0) | 13 (1.0) | 6 (0.4) | 5 (0.4) | 4 (0.3) | 0.066 |
| Mitral regurgitation | 3 (0.0) | 101 (7.5) | 67 (5.0) | 64 (4.8) | 48 (3.6) | <0.001 |
| **Medication** |  |  |  |  |  |  |
| ASA, n (%) | 3 (0.0) | 671 (50.1) | 623 (46.5) | 570 (42.5) | 501 (37.4) | <0.001 |
| Statin | 3 (0.0) | 692 (51.6) | 581 (43.4) | 574 (42.8) | 445 (33.2) | <0.001 |
| ACEi, n (%) | 3 (0.0) | 438 (32.7) | 355 (26.5) | 324 (24.2) | 291 (21.7) | <0.001 |
| ARB, n (%) | 3 (0.0) | 219 (16.3) | 186 (13.9) | 200 (14.9) | 174 (13.0) | 0.081 |
| Beta blocker, n (%) | 3 (0.0) | 786 (58.7) | 612 (45.7) | 554 (41.2) | 429 (32.0) | <0.001 |
| Thiazide, n (%) | 3 (0.0) | 181 (13.5) | 151 (11.3) | 165 (12.3) | 161 (12.0) | 0.360 |
| Loop diuretic, n (%) | 3 (0.0) | 499 (37.2) | 272 (20.3) | 201 (15.0) | 161 (12.0) | <0.001 |
| MRA, n (%) | 3 (0.0) | 159 (11.9) | 86 (6.4) | 53 (4.0) | 48 (3.6) | <0.001 |

Abbreviations. BWH: Brigham and Women's Hospital, CFR: coronary flow reserve, MBF: myocardial blood flow, A.U.: arbitrary unit, HR: heart rate, SBP: systolic blood pressure, DBP: diastolic blood pressure, BMI: body mass index, eGFR: estimated glomerular filtration rate, EF: ejection fraction, LV: left ventricle, DM: diabetes mellitus, CAD: coronary heart disease, PAD: peripheral artery disease, COPD: chronic obstructive pulmonary disease, ASA: acetyl-salicylic acid, ACEi: angiotensin-converting enzyme inhibitor, ARB: angiotensin II receptor blocker, MRA: mineralocorticoid receptor antagonist

**Table S8.**
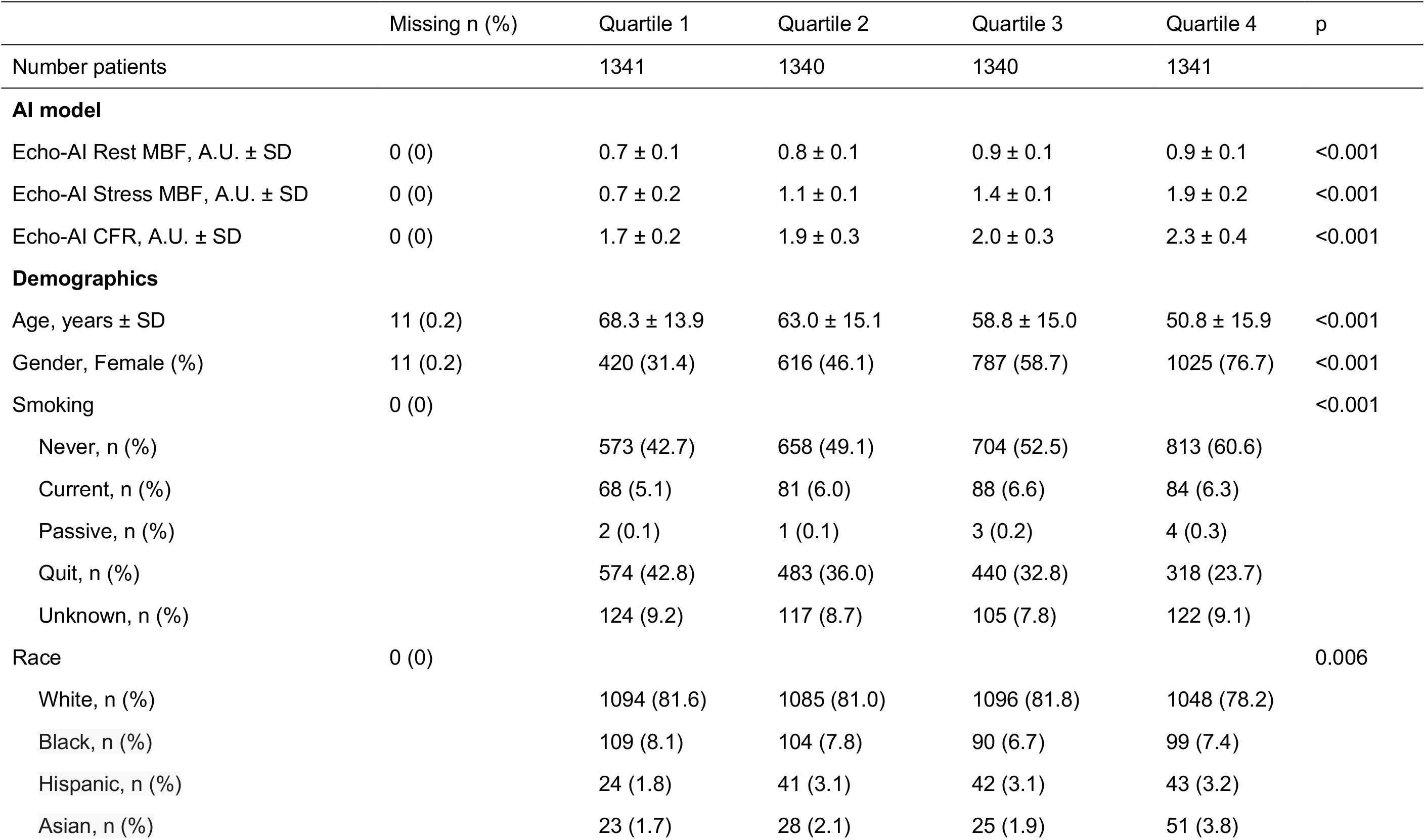
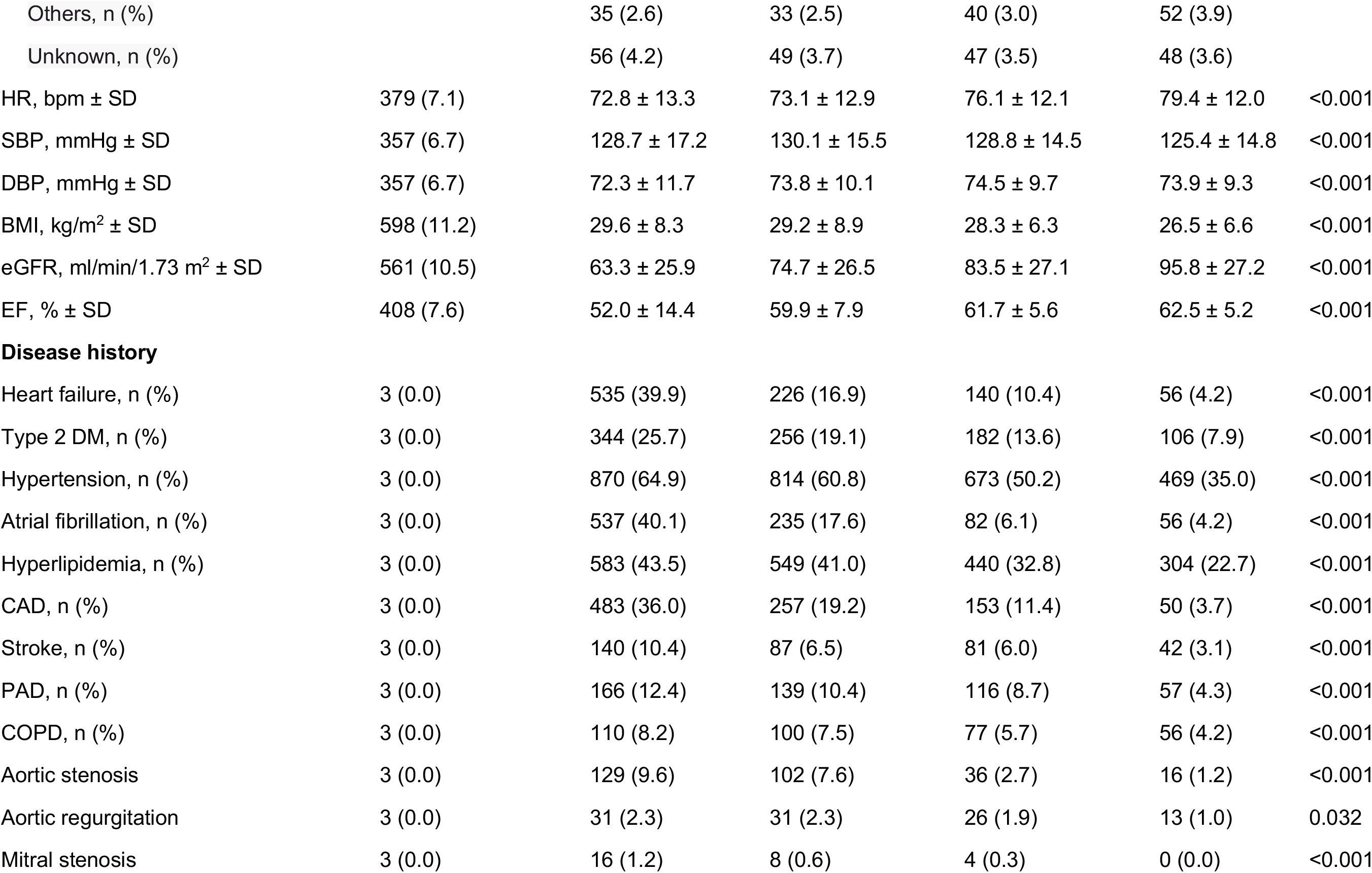
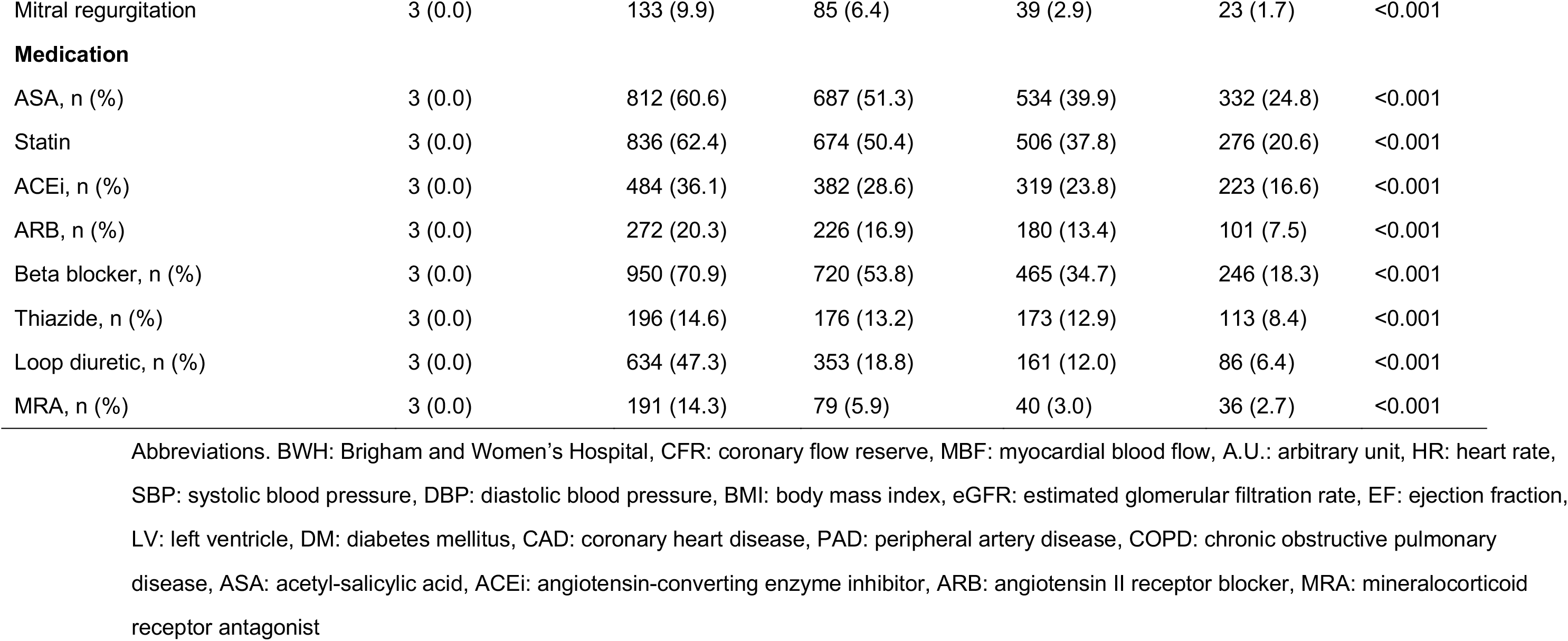
Baseline demographic for EchoAI-stressMBF quartiles for ACS survival analysis (BWH cohort)

**Table S9.** Baseline demographic for EchoAI-CFR quartiles for ACS survival analysis (BWH cohort)

|  | Missing n (%) | Quartile 1 | Quartile 2 | Quartile 3 | Quartile 4 | p |
| --- | --- | --- | --- | --- | --- | --- |
| Number patients |  | 1341 | 1340 | 1340 | 1341 |  |
| **AI model** |  |  |  |  |  |  |
| Echo-AI Rest MBF, A.U. ± SD | 0 (0) | 0.8 ± 0.1 | 0.8 ± 0.1 | 0.9 ± 0.1 | 0.9 ± 0.1 | <0.001 |
| Echo-AI Stress MBF, A.U. ± SD | 0 (0) | 0.9 ± 0.4 | 1.2 ± 0.4 | 1.4 ± 0.4 | 1.7 ± 0.4 | <0.001 |
| Echo-AI CFR, A.U. ± SD | 0 (0) | 1.6 ± 0.1 | 1.8 ± 0.1 | 2.0 ± 0.1 | 2.5 ± 0.3 | <0.001 |
| **Demographics** |  |  |  |  |  |  |
| Age, years ± SD | 11 (0.2) | 69.1 ± 14.0 | 64.3 ± 14.7 | 58.3 ± 14.6 | 49.2 ± 14.8 | <0.001 |
| Gender, Female (%) | 11 (0.2) | 590 (44.1) | 624 (46.7) | 736 (55.0) | 898 (67.1) | <0.001 |
| Smoking | 0 (0) |  |  |  |  | <0.001 |
| Never, n (%) |  | 614 (45.8) | 634 (47.3) | 699 (52.2) | 801 (59.7) |  |
| Current, n (%) |  | 61 (4.5) | 87 (6.5) | 87 (6.5) | 86 (6.4) |  |
| Passive, n (%) |  | 4 (0.3) | 1 (0.1) | 3 (0.2) | 2 (0.1) |  |
| Quit, n (%) |  | 550 (41.0) | 505 (37.7) | 452 (33.7) | 308 (23.0) |  |
| Unknown, n (%) |  | 112 (8.7) | 112 (8.4) | 113 (8.4) | 99 (7.4) |  |
| Race | 0 (0) |  |  |  |  | 0.024 |
| White, n (%) |  | 1076 (80.2) | 1102 (82.2) | 1045 (78.0) | 1100 (82.0) |  |
| Black, n (%) |  | 105 (7.8) | 102 (7.6) | 115 (8.6) | 80 (6.0) |  |
| Hispanic, n (%) |  | 40 (3.0) | 30 (2.2) | 45 (3.4) | 35 (2.6) |  |
| Asian, n (%) |  | 28 (2.1) | 33 (2.5) | 25 (1.9) | 41 (3.1) |  |
| Others, n (%) |  | 39 (2.9) | 31 (2.3) | 45 (3.4) | 45 (3.4) |  |
| Unknown, n (%) |  | 52 (4.0) | 42 (3.1) | 65 (4.9) | 40 (3.0) |  |
| HR, bpm ± SD | 379 (7.1) | 74.0 ± 12.7 | 75.1 ± 13.1 | 76.1 ± 13.1 | 76.4 ± 12.4 | <0.001 |
| SBP, mmHg ± SD | 357 (6.7) | 129.8 ± 17.1 | 130.5 ± 15.7 | 128.3 ± 15.3 | 124.5 ± 13.4 | <0.001 |
| DBP, mmHg ± SD | 357 (6.7) | 72.0 ± 10.9 | 73.8 ± 10.8 | 74.6 ± 10.0 | 74.2 ± 9.0 | <0.001 |
| BMI, kg/m<sup>2</sup> ± SD | 598 (11.2) | 29.1 ± 9.7 | 29.2 ± 7.6 | 28.6 ± 6.3 | 26.6 ± 6.4 | <0.001 |
| eGFR, ml/min/1.73 m<sup>2</sup> ± SD | 561 (10.5) | 66.2 ± 27.5 | 74.0 ± 28.1 | 82.6 ± 27.4 | 95.0 ± 25.7 | <0.001 |
| EF, % ± SD | 408 (7.6) | 55.4 ± 13.7 | 58.7 ± 10.2 | 60.6 ± 7.3 | 61.7 ± 5.2 | <0.001 |
| **Disease history** |  |  |  |  |  |  |
| Heart failure, n (%) | 3 (0.0) | 444 (33.1) | 278 (20.8) | 165 (12.3) | 70 (5.2) | <0.001 |
| Type 2 DM, n (%) | 3 (0.0) | 356 (26.6) | 262 (19.6) | 194 (14.5) | 76 (5.7) | <0.001 |
| Hypertension, n (%) | 3 (0.0) | 901 (67.2) | 827 (61.8) | 691 (51.6) | 407 (30.4) | <0.001 |
| Atrial fibrillation, n (%) | 3 (0.0) | 391 (29.2) | 302 (22.6) | 145 (10.8) | 72 (5.4) | <0.001 |
| Hyperlipidemia, n (%) | 3 (0.0) | 590 (44.0) | 548 (41.0) | 452 (33.7) | 286 (21.3) | <0.001 |
| CAD, n (%) | 3 (0.0) | 441 (32.9) | 289 (21.6) | 155 (11.6) | 58 (4.3) | <0.001 |
| Stroke, n (%) | 3 (0.0) | 154 (11.5) | 92 (6.9) | 62 (4.6) | 42 (3.1) | <0.001 |
| PAD, n (%) | 3 (0.0) | 189 (14.1) | 137 (10.2) | 84 (6.3) | 68 (5.1) | <0.001 |
| COPD, n (%) | 3 (0.0) | 124 (9.3) | 106 (7.9) | 66 (4.9) | 47 (3.5) | <0.001 |
| Aortic stenosis | 3 (0.0) | 149 (11.1) | 85 (6.4) | 38 (2.8) | 11 (0.8) | <0.001 |
| Aortic regurgitation | 3 (0.0) | 27 (2.0) | 36 (2.7) | 20 (1.5) | 18 (1.3) | 0.045 |
| Mitral stenosis | 3 (0.0) | 16 (1.2) | 7 (0.5) | 5 (0.4) | 0 (0.0) | <0.001 |
| Mitral regurgitation | 3 (0.0) | 121 (9.0) | 89 (6.7) | 41 (3.1) | 29 (2.2) | <0.001 |
| **Medication** |  |  |  |  |  |  |
| ASA, n (%) | 3 (0.0) | 812 (60.6) | 699 (52.2) | 513 (38.3) | 341 (25.4) | <0.001 |
| Statin | 3 (0.0) | 803 (59.9) | 700 (52.3) | 520 (38.8) | 269 (20.1) | <0.001 |
| ACEi, n (%) | 3 (0.0) | 448 (33.4) | 416 (31.1) | 343 (25.6) | 201 (15.0) | <0.001 |
| ARB, n (%) | 3 (0.0) | 269 (20.1) | 227 (17.0) | 187 (14.0) | 96 (7.2) | <0.001 |
| Beta blocker, n (%) | 3 (0.0) | 873 (65.1) | 743 (55.5) | 482 (36.0) | 283 (21.1) | <0.001 |
| Thiazide, n (%) | 3 (0.0) | 192 (14.3) | 197 (14.7) | 174 (13.0) | 95 (7.1) | <0.001 |
| Loop diuretic, n (%) | 3 (0.0) | 521 (38.9) | 358 (26.8) | 192 (14.3) | 62 (4.6) | <0.001 |
| MRA, n (%) | 3 (0.0) | 156 (11.6) | 109 (8.1) | 53 (4.0) | 28 (2.1) | <0.001 |

Abbreviations. BWH: Brigham and Women's Hospital, CFR: coronary flow reserve, MBF: myocardial blood flow, A.U.: arbitrary unit, HR: heart rate, SBP: systolic blood pressure, DBP: diastolic blood pressure, BMI: body mass index, eGFR: estimated glomerular filtration rate, EF: ejection fraction, LV: left ventricle, DM: diabetes mellitus, CAD: coronary heart disease, PAD: peripheral artery disease, COPD: chronic obstructive pulmonary disease, ASA: acetyl-salicylic acid, ACEi: angiotensin-converting enzyme inhibitor, ARB: angiotensin II receptor blocker, MRA: mineralocorticoid receptor antagonist

**Table S10.** Baseline demographic for EchoAI-restMBF quartiles for heart failure survival analysis (MGH cohort)

|  | Missing n (%) | Quartile 1 | Quartile 2 | Quartile 3 | Quartile 4 | p |
| --- | --- | --- | --- | --- | --- | --- |
| Number patients |  | 1,285 | 1,284 | 1,284 | 1,284 |  |
| **AI model** |  |  |  |  |  |  |
| Echo-AI Rest MBF, A.U. ± SD | 0 (0) | 0.7 ± 0.0 | 0.8 ± 0.0 | 0.9 ± 0.0 | 1.0 ± 0.1 | <0.001 |
| Echo-AI Stress MBF, A.U. ± SD | 0 (0) | 0.8 ± 0.3 | 1.1 ± 0.3 | 1.2 ± 0.3 | 1.4 ± 0.4 | <0.001 |
| Echo-AI CFR, A.U. ± SD | 0 (0) | 1.9 ± 0.3 | 1.9 ± 0.3 | 1.9 ± 0.3 | 1.9 ± 0.3 | <0.001 |
| **Demographics** |  |  |  |  |  |  |
| Age, years ± SD | 0 (0.0) | 68.1 ± 15.6 | 65.6 ± 15.9 | 66.2 ± 15.6 | 64.5 ± 16.5 | <0.001 |
| Gender, Female (%) | 0 (0.0) | 309 (24.0) | 527 (41.0) | 641 (49.9) | 772 (60.1) | <0.001 |
| Smoking | 0 (0) |  |  |  |  | 0.142 |
| Never, n (%) |  | 534 (41.6) | 566 (44.1) | 598 (46.6) | 617 (48.1) |  |
| Current, n (%) |  | 116 (9.0) | 112 (8.7) | 103 (8.0) | 106 (8.3) |  |
| Passive, n (%) |  | 1 (0.1) | 0 (0.0) | 0 (0.0) | 0 (0.0) |  |
| Quit, n (%) |  | 496 (38.6) | 490 (38.2) | 463 (36.1) | 443 (34.5) |  |
| Unknown, n (%) |  | 138 (10.7) | 116 (9.0) | 120 (9.3) | 118 (9.2) |  |
| Race | 0 (0) |  |  |  |  | 0.253 |
| White, n (%) |  | 1,123 (87.4) | 1,084 (84.4) | 1,095 (85.3) | 1,075 (83.7) |  |
| Black, n (%) |  | 58 (4.5) | 64 (5.0) | 66 (5.1) | 77 (6.0) |  |
| Hispanic, n (%) |  | 9 (0.7) | 11 (0.9) | 9 (0.7) | 5 (0.4) |  |
| Asian, n (%) |  | 21 (1.6) | 35 (2.7) | 38 (3.0) | 43 (3.3) |  |
| Others, n (%) |  | 43 (3.3) | 59 (4.6) | 52 (4.0) | 57 (4.4) |  |
| Unknown, n (%) |  | 31 (2.4) | 31 (2.4) | 24 (1.9) | 27 (2.1) |  |
| HR, bpm ± SD | 31 (0.6) | 73.7 ± 13.0 | 72.5 ± 11.5 | 73.5 ± 11.3 | 76.5 ± 10.7 | <0.001 |
| SBP, mmHg ± SD | 27 (0.5) | 125.1 ± 14.7 | 127.2 ± 14.1 | 127.6 ± 13.4 | 129.3 ± 14.7 | <0.001 |
| DBP, mmHg ± SD | 27 (0.5) | 70.1 ± 8.1 | 70.5 ± 8.2 | 70.3 ± 7.6 | 70.7 ± 8.3 | 0.229 |
| BMI, kg/m<sup>2</sup> ± SD | 82 (1.6) | 29.6 ± 8.0 | 28.8 ± 6.5 | 28.6 ± 8.1 | 27.8 ± 6.5 | <0.001 |
| eGFR, ml/min/1.73 m<sup>2</sup> ± SD | 374(7.3) | 61.9 ± 26.8 | 70.9 ± 27.2 | 72.5 ± 29.0 | 75.5 ± 32.9 | <0.001 |
| EF, % ± SD | 409 (8.0) | 54.0 ± 16.4 | 63.2 ± 10.9 | 65.7 ± 9.1 | 67.9 ± 8.2 | <0.001 |
| **Disease history** |  |  |  |  |  |  |
| Heart failure, n (%) | 0 (0.0) | 685 (53.3) | 428 (33.3) | 311 (24.2) | 265 (20.6) | <0.001 |
| Type 2 DM, n (%) | 0 (0.0) | 366 (28.5) | 320 (24.9) | 298 (23.2) | 313 (24.4) | 0.014 |
| Hypertension, n (%) | 0 (0.0) | 1,016 (79.1) | 963 (75.0) | 945 (73.6) | 925 (72.0) | <0.001 |
| Atrial fibrillation, n (%) | 0 (0.0) | 760 (59.1) | 466 (36.3) | 361 (28.1) | 237 (18.5) | <0.001 |
| Hyperlipidemia, n (%) | 0 (0.0) | 857 (66.7) | 811 (63.2) | 794 (61.8) | 750 (58.4) | <0.001 |
| CAD, n (%) | 0 (0.0) | 652 (50.7) | 498 (38.8) | 446 (34.7) | 353 (27.5) | <0.001 |
| Stroke, n (%) | 0 (0.0) | 154 (12.0) | 154 (12.0) | 143 (11.1) | 117 (9.1) | 0.065 |
| PAD, n (%) | 0 (0.0) | 283 (22.0) | 276 (21.5) | 257 (20.0) | 245 (19.1) | 0.232 |
| COPD, n (%) | 0 (0.0) | 186 (14.5) | 147 (11.4) | 161 (12.5) | 166 (12.9) | 0.145 |
| Aortic stenosis | 0 (0.0) | 110 (8.6) | 151 (11.8) | 142 (11.1) | 124 (9.7) | 0.036 |
| Aortic regurgitation | 0 (0.0) | 34 (2.6) | 46 (3.6) | 36 (2.8) | 41 (3.2) | 0.515 |
| Mitral stenosis | 0 (0.0) | 11 (0.9) | 14 (1.1) | 14 (1.1) | 11 (0.9) | 0.866 |
| Mitral regurgitation | 0 (0.0) | 47 (3.7) | 49 (3.8) | 33 (2.6) | 41 (3.2) | 0.288 |
| **Medication** |  |  |  |  |  |  |
| ASA, n (%) | 0 (0.0) | 823 (64.0) | 804 (62.6) | 789 (61.4) | 732 (57.0) | 0.002 |
| Statin | 0 (0.0) | 846 (65.8) | 755 (58.8) | 718 (55.9) | 686 (53.4) | <0.001 |
| ACEi, n (%) | 0 (0.0) | 466 (36.3) | 385 (30.0) | 358 (27.9) | 336 (26.2) | <0.001 |
| ARB, n (%) | 0 (0.0) | 263 (20.5) | 194 (15.1) | 212 (16.5) | 208 (16.3) | 0.002 |
| Beta blocker, n (%) | 0 (0.0) | 923 (71.8) | 757 (59.0) | 696 (54.2) | 644 (50.2) | <0.001 |
| Thiazide, n (%) | 0 (0.0) | 185 (14.4) | 192 (15.0) | 184 (14.3) | 185 (14.4) | 0.967 |
| Loop diuretic, n (%) | 0 (0.0) | 646 (50.3) | 389 (30.3) | 334 (26.0) | 316 (24.6) | <0.001 |
| MRA, n (%) | 0 (0.0) | 169 (13.2) | 73 (5.7) | 63 (4.9) | 64 (5.0) | <0.001 |

Abbreviations. MGH: Massachusetts General Hospital, CFR: coronary flow reserve, MBF: myocardial blood flow, A.U.: arbitrary unit, HR: heart rate, SBP: systolic blood pressure, DBP: diastolic blood pressure, BMI: body mass index, eGFR: estimated glomerular filtration rate, EF: ejection fraction, LV: left ventricle, DM: diabetes mellitus, CAD: coronary heart disease, PAD: peripheral artery disease, COPD: chronic obstructive pulmonary disease, ASA: acetyl-salicylic acid, ACEi: angiotensin-converting enzyme inhibitor, ARB: angiotensin II receptor blocker, MRA: mineralocorticoid receptor antagonist

**Table S11.** Baseline demographic for EchoAI-stressMBF quartiles for heart failure survival analysis (MGH cohort)

|  | Missing n (%) | Quartile 1 | Quartile 2 | Quartile 3 | Quartile 4 | p |
| --- | --- | --- | --- | --- | --- | --- |
| Number patients |  | 1,285 | 1,284 | 1,284 | 1,284 |  |
| **AI model** |  |  |  |  |  |  |
| Echo-AI Rest MBF, A.U. ± SD | 0 (0) | 0.7 ± 0.1 | 0.8 ± 0.1 | 0.9 ± 0.1 | 0.9 ± 0.1 | <0.001 |
| Echo-AI Stress MBF, A.U. ± SD | 0 (0) | 0.6 ± 0.1 | 0.9 ± 0.1 | 1.2 ± 0.1 | 1.6 ± 0.2 | <0.001 |
| Echo-AI CFR, A.U. ± SD | 0 (0) | 1.7 ± 0.2 | 1.8 ± 0.2 | 2.0 ± 0.2 | 2.2 ± 0.3 | <0.001 |
| **Demographics** |  |  |  |  |  |  |
| Age, years ± SD | 0 (0.0) | 71.8 ± 13.2 | 69.5 ± 14.2 | 65.5 ± 15.4 | 57.6 ± 17.0 | <0.001 |
| Gender, Female (%) | 0 (0.0) | 347 (27.0) | 523 (40.7) | 587 (45.7) | 792 (61.7) | <0.001 |
| Smoking | 0 (0) |  |  |  |  | <0.001 |
| Never, n (%) |  | 514 (40.0) | 571 (44.5) | 579 (45.1) | 651 (50.7) |  |
| Current, n (%) |  | 86 (6.7) | 98 (7.6) | 117 (9.1) | 136 (10.6) |  |
| Passive, n (%) |  | 0 (0.0) | 1 (0.1) | 0 (0.0) | 0 (0.0) |  |
| Quit, n (%) |  | 545 (42.4) | 491 (38.2) | 435 (36.2) | 391 (30.5) |  |
| Unknown, n (%) |  | 140 (10.9) | 123 (9.6) | 123 (9.6) | 106 (8.3) |  |
| Race | 0 (0) |  |  |  |  | 0.015 |
| White, n (%) |  | 1,128 (87.8) | 1,115 (86.8) | 1,079 (84.0) | 1,055 (82.2) |  |
| Black, n (%) |  | 60 (4.7) | 55 (4.3) | 73 (5.7) | 77 (6.0) |  |
| Hispanic, n (%) |  | 7 (0.5) | 7 (0.5) | 8 (0.6) | 12 (0.9) |  |
| Asian, n (%) |  | 22 (1.7) | 30 (2.3) | 34 (2.6) | 51 (4.0) |  |
| Others, n (%) |  | 43 (3.3) | 46 (3.6) | 60 (4.7) | 62 (4.8) |  |
| Unknown, n (%) |  | 25 (1.9) | 31 (2.4) | 30 (2.3) | 27 (2.1) |  |
| HR, bpm ± SD | 31 (0.6) | 72.5 ± 11.8 | 72.1 ± 11.5 | 73.3 ± 11.5 | 78.3 ± 11.3 | <0.001 |
| SBP, mmHg ± SD | 27 (0.5) | 127.2 ± 15.3 | 128.4 ± 14.2 | 127.7 ± 13.7 | 125.8 ± 13.8 | <0.001 |
| DBP, mmHg ± SD | 27 (0.5) | 69.1 ± 8.1 | 70.1 ± 7.7 | 71.0 ± 8.0 | 71.4 ± 8.2 | <0.001 |
| BMI, kg/m<sup>2</sup> ± SD | 82 (1.6) | 29.8 ± 8.3 | 29.2 ± 7.1 | 28.6 ± 6.4 | 27.2 ± 7.1 | <0.001 |
| eGFR, ml/min/1.73 m<sup>2</sup> ± SD | 374(7.3) | 56.8 ± 25.7 | 64.8 ± 25.5 | 73.4 ± 28.8 | 85.8 ± 29.7 | <0.001 |
| EF, % ± SD | 409 (8.0) | 53.9 ± 16.7 | 63.1 ± 11.5 | 66.1 ± 8.8 | 67.5 ± 7.0 | <0.001 |
| **Disease history** |  |  |  |  |  |  |
| Heart failure, n (%) | 0 (0.0) | 780 (60.7) | 505 (39.3) | 290 (22.6) | 114 (8.9) | <0.001 |
| Type 2 DM, n (%) | 0 (0.0) | 480 (37.4) | 327 (25.5) | 284 (22.1) | 206 (16.0) | <0.001 |
| Hypertension, n (%) | 0 (0.0) | 1,109 (86.3) | 1,032 (80.4) | 950 (74.0) | 758 (59.0) | <0.001 |
| Atrial fibrillation, n (%) | 0 (0.0) | 774 (60.2) | 565 (44.0) | 338 (26.3) | 147 (11.4) | <0.001 |
| Hyperlipidemia, n (%) | 0 (0.0) | 939 (73.1) | 882 (68.7) | 802 (62.5) | 589 (45.9) | <0.001 |
| CAD, n (%) | 0 (0.0) | 779 (60.6) | 557 (43.4) | 421 (32.8) | 192 (15.0) | <0.001 |
| Stroke, n (%) | 0 (0.0) | 186 (14.5) | 144 (11.2) | 134 (10.4) | 104 (8.1) | <0.001 |
| PAD, n (%) | 0 (0.0) | 330 (25.7) | 319 (24.8) | 230 (17.9) | 182 (14.2) | <0.001 |
| COPD, n (%) | 0 (0.0) | 204 (15.9) | 160 (12.5) | 155 (12.1) | 141 (11.0) | 0.002 |
| Aortic stenosis | 0 (0.0) | 172 (13.4) | 158 (12.3) | 131 (10.2) | 66 (5.1) | <0.001 |
| Aortic regurgitation | 0 (0.0) | 46 (3.7) | 50 (3.9) | 35 (2.7) | 24 (1.9) | 0.009 |
| Mitral stenosis | 0 (0.0) | 14 (1.1) | 19 (1.5) | 8 (0.6) | 9 (0.7) | 0.101 |
| Mitral regurgitation | 0 (0.0) | 59 (4.6) | 51 (4.0) | 40 (3.1) | 20 (1.6) | <0.001 |
| **Medication** |  |  |  |  |  |  |
| ASA, n (%) | 0 (0.0) | 938 (73.0) | 861 (67.1) | 808 (62.9) | 541 (42.1) | <0.001 |
| Statin | 0 (0.0) | 966 (75.2) | 825 (64.3) | 736 (57.3) | 478 (37.2) | <0.001 |
| ACEi, n (%) | 0 (0.0) | 504 (39.2) | 397 (30.9) | 361 (28.1) | 283 (22.0) | <0.001 |
| ARB, n (%) | 0 (0.0) | 279 (21.7) | 235 (18.3) | 218 (17.0) | 145 (11.3) | <0.001 |
| Beta blocker, n (%) | 0 (0.0) | 1,012 (78.8) | 881 (68.6) | 687 (53.5) | 440 (34.3) | <0.001 |
| Thiazide, n (%) | 0 (0.0) | 213 (16.6) | 200 (15.6) | 194 (15.1) | 139 (10.8) | <0.001 |
| Loop diuretic, n (%) | 0 (0.0) | 728 (56.7) | 481 (37.5) | 291 (22.7) | 185 (14.4) | <0.001 |
| MRA, n (%) | 0 (0.0) | 171 (13.3) | 83 (6.5) | 65 (5.1) | 50 (3.9) | <0.001 |

Abbreviations. MGH: Massachusetts General Hospital, CFR: coronary flow reserve, MBF: myocardial blood flow, A.U.: arbitrary unit, HR: heart rate, SBP: systolic blood pressure, DBP: diastolic blood pressure, BMI: body mass index, eGFR: estimated glomerular filtration rate, EF: ejection fraction, LV: left ventricle, DM: diabetes mellitus, CAD: coronary heart disease, PAD: peripheral artery disease, COPD: chronic obstructive pulmonary disease, ASA: acetyl-salicylic acid, ACEi: angiotensin-converting enzyme inhibitor, ARB: angiotensin II receptor blocker, MRA: mineralocorticoid receptor antagonist

**Table S12.** Baseline demographic for EchoAI-CFR quartiles for heart failure survival analysis (MGH cohort)

|  | Missing n (%) | Quartile 1 | Quartile 2 | Quartile 3 | Quartile 4 | p |
| --- | --- | --- | --- | --- | --- | --- |
| Number patients |  | 1,285 | 1,284 | 1,284 | 1,284 |  |
| **AI model** |  |  |  |  |  |  |
| Echo-AI Rest MBF, A.U. ± SD | 0 (0) | 0.8 ± 0.1 | 0.8 ± 0.1 | 0.8 ± 0.1 | 0.8 ± 0.1 | <0.001 |
| Echo-AI Stress MBF, A.U. ± SD | 0 (0) | 0.8 ± 0.3 | 1.0 ± 0.3 | 1.1 ± 0.3 | 1.4 ± 0.3 | <0.001 |
| Echo-AI CFR, A.U. ± SD | 0 (0) | 1.6 ± 0.1 | 1.8 ± 0.0 | 2.0 ± 0.1 | 2.3 ± 0.2 | <0.001 |
| **Demographics** |  |  |  |  |  |  |
| Age, years ± SD | 0 (0.0) | 73.0 ± 13.0 | 70.1 ± 13.6 | 65.8 ± 14.5 | 55.6 ± 16.7 | <0.001 |
| Gender, Female (%) | 0 (0.0) | 518 (40.3) | 508 (39.6) | 588 (45.8) | 635 (49.5) | <0.001 |
| Smoking | 0 (0) |  |  |  |  | <0.001 |
| Never, n (%) |  | 533 (41.5) | 543 (42.3) | 579 (45.1) | 660 (51.4) |  |
| Current, n (%) |  | 87 (6.8) | 88 (6.9) | 125 (9.7) | 137 (10.7) |  |
| Passive, n (%) |  | 0 (0.0) | 1 (0.1) | 0 (0.0) | 0 (0.0) |  |
| Quit, n (%) |  | 545 (42.4) | 517 (40.3) | 446 (34.7) | 384 (29.9) |  |
| Unknown, n (%) |  | 120 (9.3) | 135 (10.5) | 134 (10.4) | 103 (8.0) |  |
| Race | 0 (0) |  |  |  |  | 0.144 |
| White, n (%) |  | 1,127 (87.7) | 1,096 (85.4) | 1,087 (84.7) | 1,067 (83.1) |  |
| Black, n (%) |  | 54 (4.2) | 68 (5.3) | 67 (5.2) | 76 (5.9) |  |
| Hispanic, n (%) |  | 3 (0.2) | 11 (0.9) | 11 (0.9) | 9 (0.7) |  |
| Asian, n (%) |  | 34 (2.6) | 32 (2.5) | 35 (2.7) | 36 (2.8) |  |
| Others, n (%) |  | 38 (3.0) | 47 (3.7) | 58 (4.5) | 68 (5.3) |  |
| Unknown, n (%) |  | 29 (2.3) | 30 (2.3) | 26 (2.0) | 28 (2.2) |  |
| HR, bpm ± SD | 31 (0.6) | 73.0 ± 10.5 | 73.8 ± 12.0 | 74.8 ± 12.0 | 74.6 ± 12.4 | 0.001 |
| SBP, mmHg ± SD | 27 (0.5) | 128.7 ± 14.8 | 128.7 ± 14.5 | 127.9 ± 14.2 | 123.9 ± 13.0 | <0.001 |
| DBP, mmHg ± SD | 27 (0.5) | 68.8 ± 7.9 | 70.2 ± 7.9 | 71.0 ± 8.0 | 71.7 ± 8.1 | <0.001 |
| BMI, kg/m<sup>2</sup> ± SD | 82 (1.6) | 29.6 ± 7.6 | 29.0 ± 6.4 | 28.9 ± 8.0 | 27.2 ± 7.1 | <0.001 |
| eGFR, ml/min/1.73 m<sup>2</sup> ± SD | 374(7.3) | 58.9 ± 27.2 | 64.3 ± 27.3 | 72.4 ± 27.9 | 85.5 ± 28.6 | <0.001 |
| EF, % ± SD | 409 (8.0) | 58.8 ± 16.1 | 61.8 ± 13.5 | 64.4 ± 10.8 | 65.7 ± 8.1 | <0.001 |
| **Disease history** |  |  |  |  |  |  |
| Heart failure, n (%) | 0 (0.0) | 658 (51.2) | 531 (41.4) | 359 (28.0) | 141 (11.0) | <0.001 |
| Type 2 DM, n (%) | 0 (0.0) | 453 (35.3) | 353 (27.5) | 315 (24.5) | 176 (13.7) | <0.001 |
| Hypertension, n (%) | 0 (0.0) | 1,091 (84.9) | 1,056 (82.2) | 993 (77.3) | 709 (55.2) | <0.001 |
| Atrial fibrillation, n (%) | 0 (0.0) | 636 (49.5) | 561 (43.7) | 425 (33.1) | 202 (15.7) | <0.001 |
| Hyperlipidemia, n (%) | 0 (0.0) | 931 (72.5) | 882 (68.7) | 788 (61.4) | 611 (47.6) | <0.001 |
| CAD, n (%) | 0 (0.0) | 714 (55.6) | 573 (44.6) | 399 (31.1) | 263 (20.5) | <0.001 |
| Stroke, n (%) | 0 (0.0) | 194 (15.1) | 129 (10.0) | 153 (11.9) | 92 (7.2) | <0.001 |
| PAD, n (%) | 0 (0.0) | 320 (24.9) | 284 (22.1) | 261 (20.3) | 196 (15.3) | <0.001 |
| COPD, n (%) | 0 (0.0) | 213 (16.6) | 178 (13.9) | 168 (13.1) | 101 (7.9) | <0.001 |
| Aortic stenosis | 0 (0.0) | 201 (15.6) | 143 (11.1) | 119 (9.3) | 64 (5.0) | <0.001 |
| Aortic regurgitation | 0 (0.0) | 41 (3.2) | 41 (3.2) | 45 (3.5) | 30 (2.3) | 0.351 |
| Mitral stenosis | 0 (0.0) | 16 (1.2) | 17 (1.3) | 10 (0.8) | 7 (0.5) | 0.135 |
| Mitral regurgitation | 0 (0.0) | 52 (4.0) | 49 (3.8) | 37 (2.9) | 32 (2.5) | 0.085 |
| **Medication** |  |  |  |  |  |  |
| ASA, n (%) | 0 (0.0) | 940 (73.2) | 834 (65.0) | 786 (61.2) | 588 (45.8) | <0.001 |
| Statin | 0 (0.0) | 934 (73.0) | 850 (66.2) | 754 (58.7) | 463 (36.1) | <0.001 |
| ACEi, n (%) | 0 (0.0) | 460 (35.8) | 435 (33.9) | 377 (29.4) | 273 (21.3) | <0.001 |
| ARB, n (%) | 0 (0.0) | 270 (21.0) | 246 (19.2) | 215 (16.7) | 146 (11.4) | <0.001 |
| Beta blocker, n (%) | 0 (0.0) | 958 (74.6) | 864 (67.3) | 736 (57.3) | 462 (36.0) | <0.001 |
| Thiazide, n (%) | 0 (0.0) | 217 (16.9) | 220 (17.1) | 183 (14.3) | 126 (9.8) | <0.001 |
| Loop diuretic, n (%) | 0 (0.0) | 650 (50.6) | 501 (39.0) | 371 (28.9) | 163 (12.7) | <0.001 |
| MRA, n (%) | 0 (0.0) | 143 (11.1) | 98 (7.6) | 90 (7.0) | 38 (3.0) | <0.001 |

Abbreviations. MGH: Massachusetts General Hospital, CFR: coronary flow reserve, MBF: myocardial blood flow, A.U.: arbitrary unit, HR: heart rate, SBP: systolic blood pressure, DBP: diastolic blood pressure, BMI: body mass index, eGFR: estimated glomerular filtration rate, EF: ejection fraction, LV: left ventricle, DM: diabetes mellitus, CAD: coronary heart disease, PAD: peripheral artery disease, COPD: chronic obstructive pulmonary disease, ASA: acetyl-salicylic acid, ACEi: angiotensin-converting enzyme inhibitor, ARB: angiotensin II receptor blocker, MRA: mineralocorticoid receptor antagonist

**Table S13.**
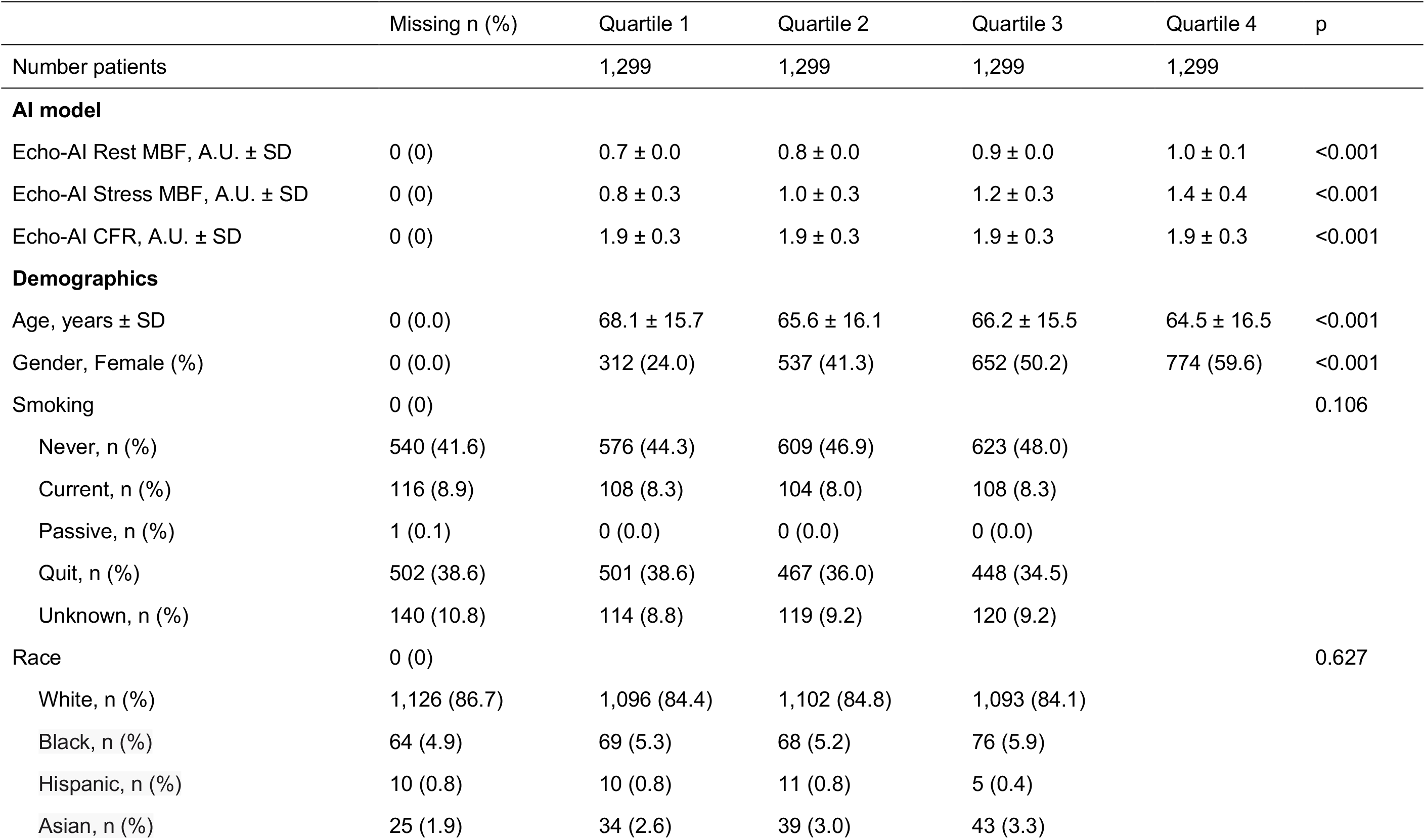
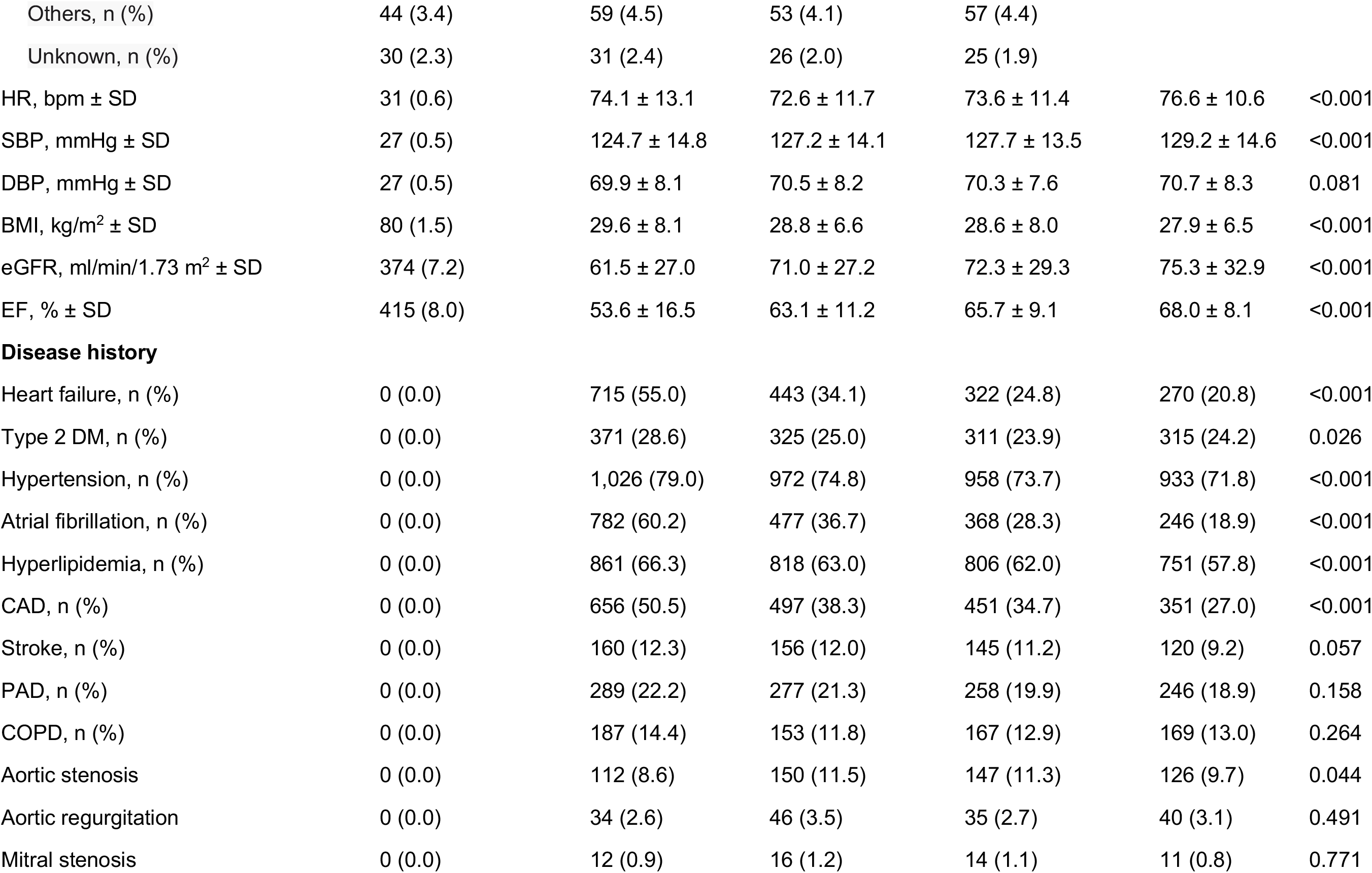
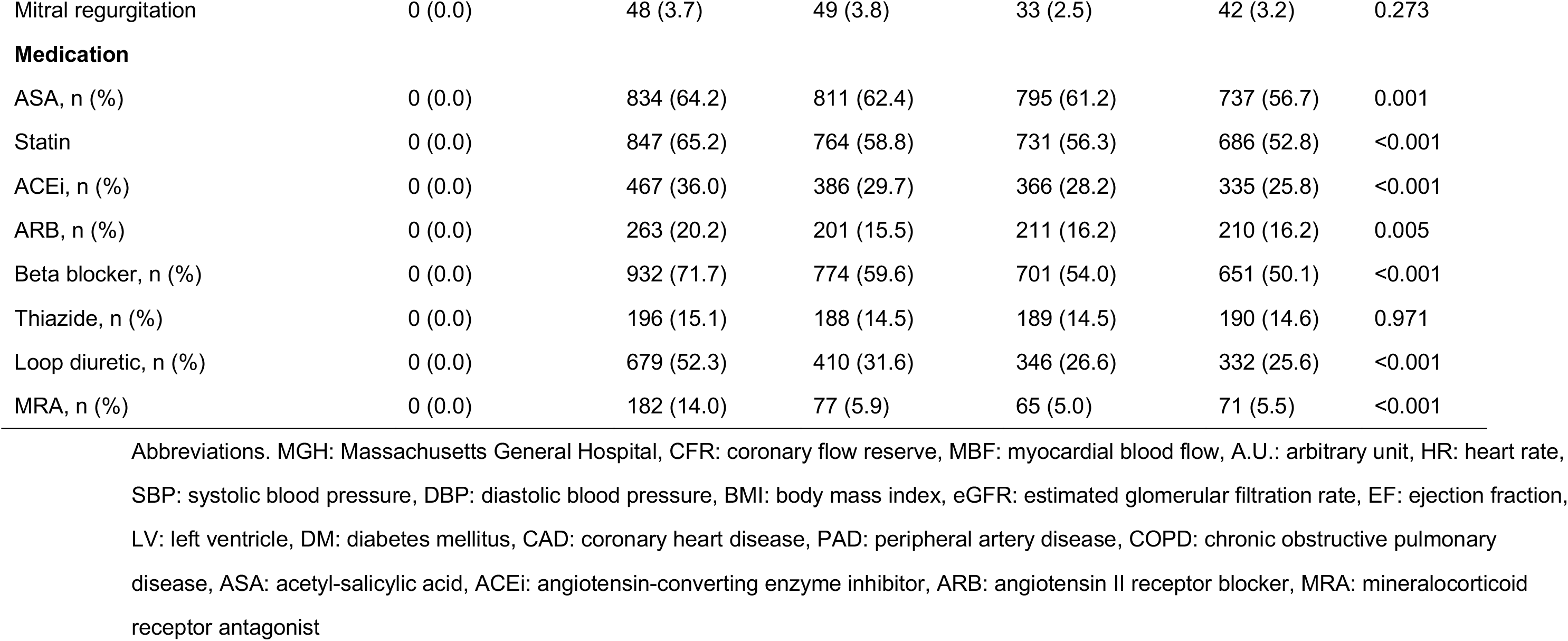
Baseline demographic for EchoAI-restMBF quartiles for ACS survival analysis (MGH cohort)

**Table S14.**
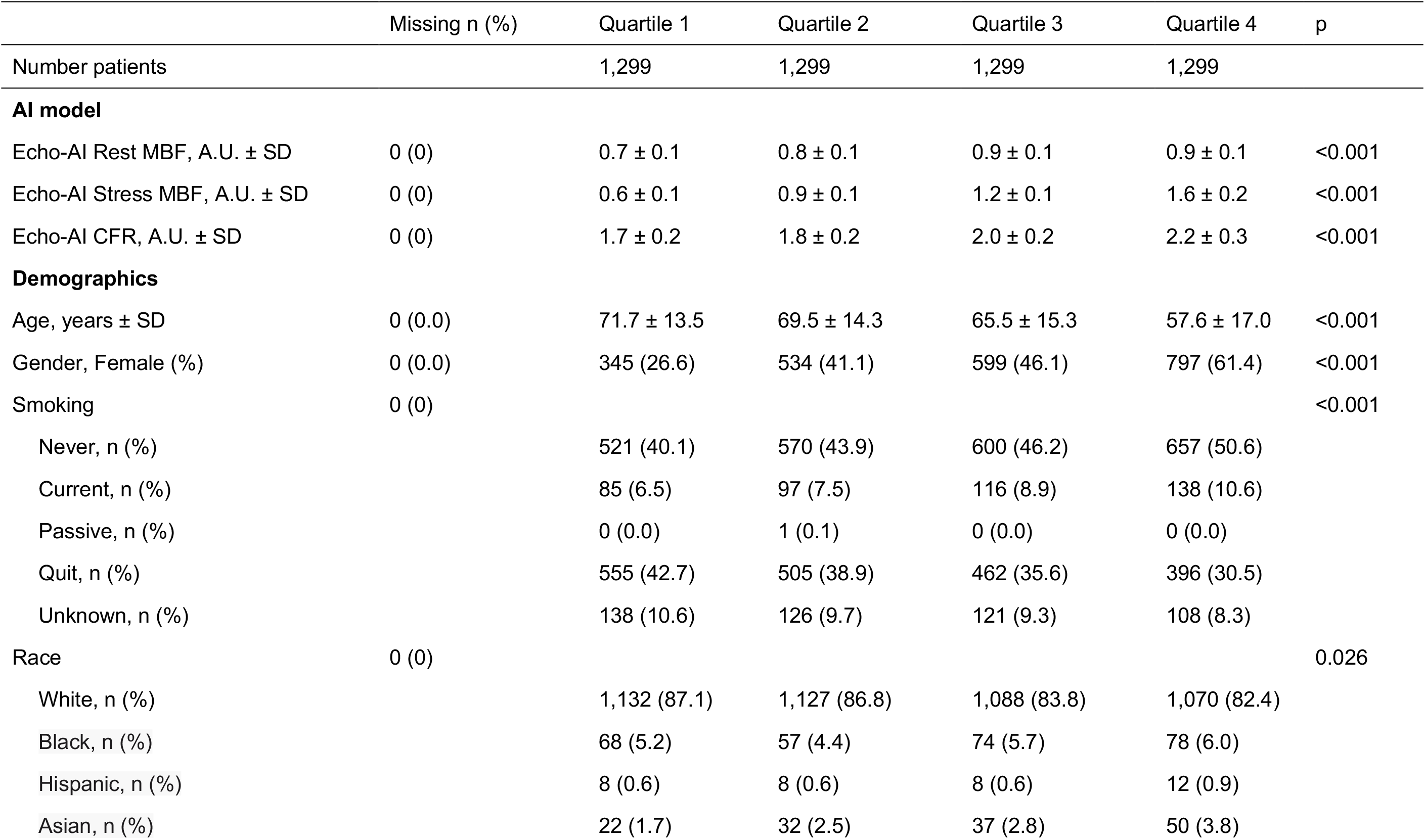
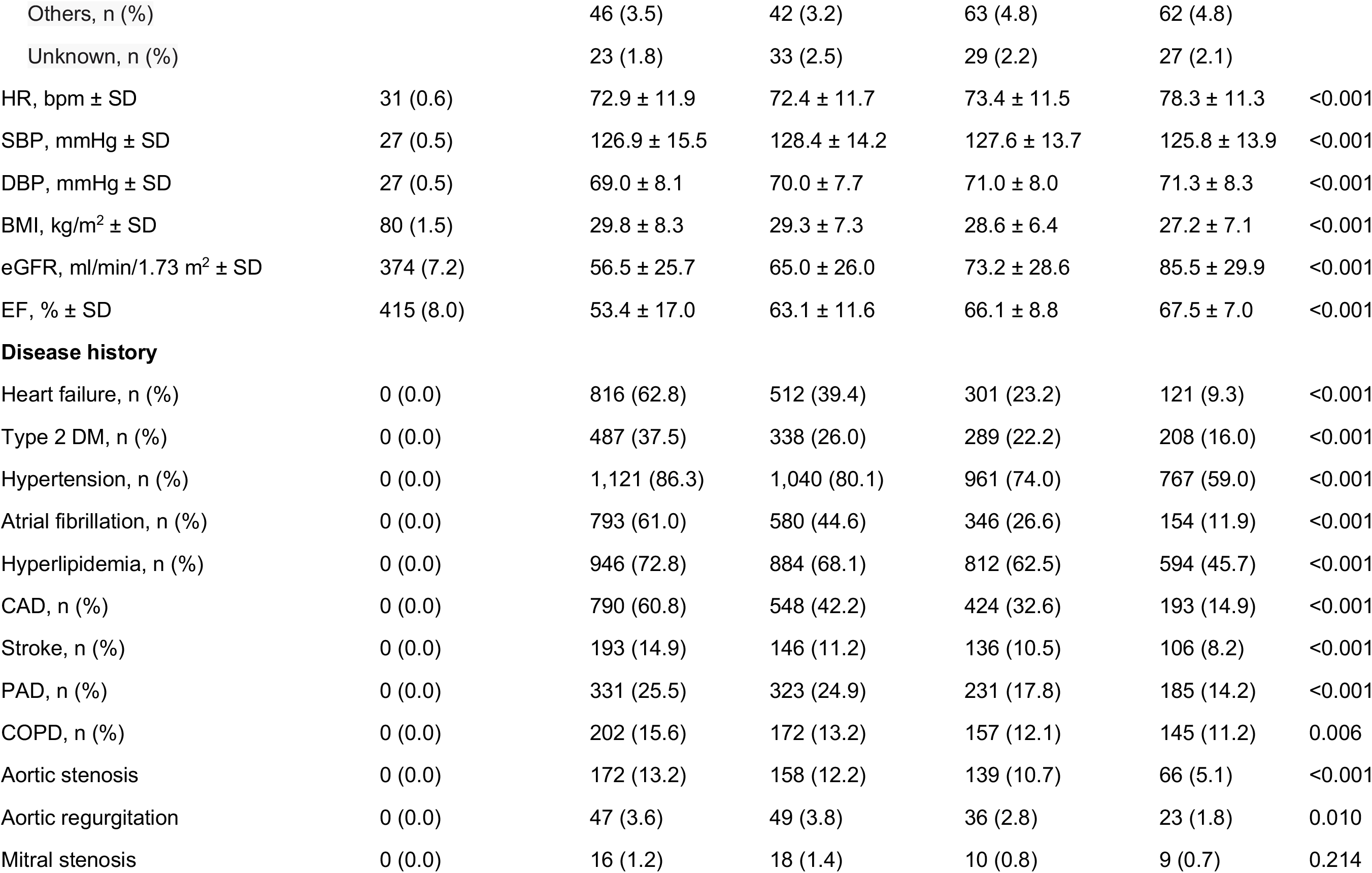
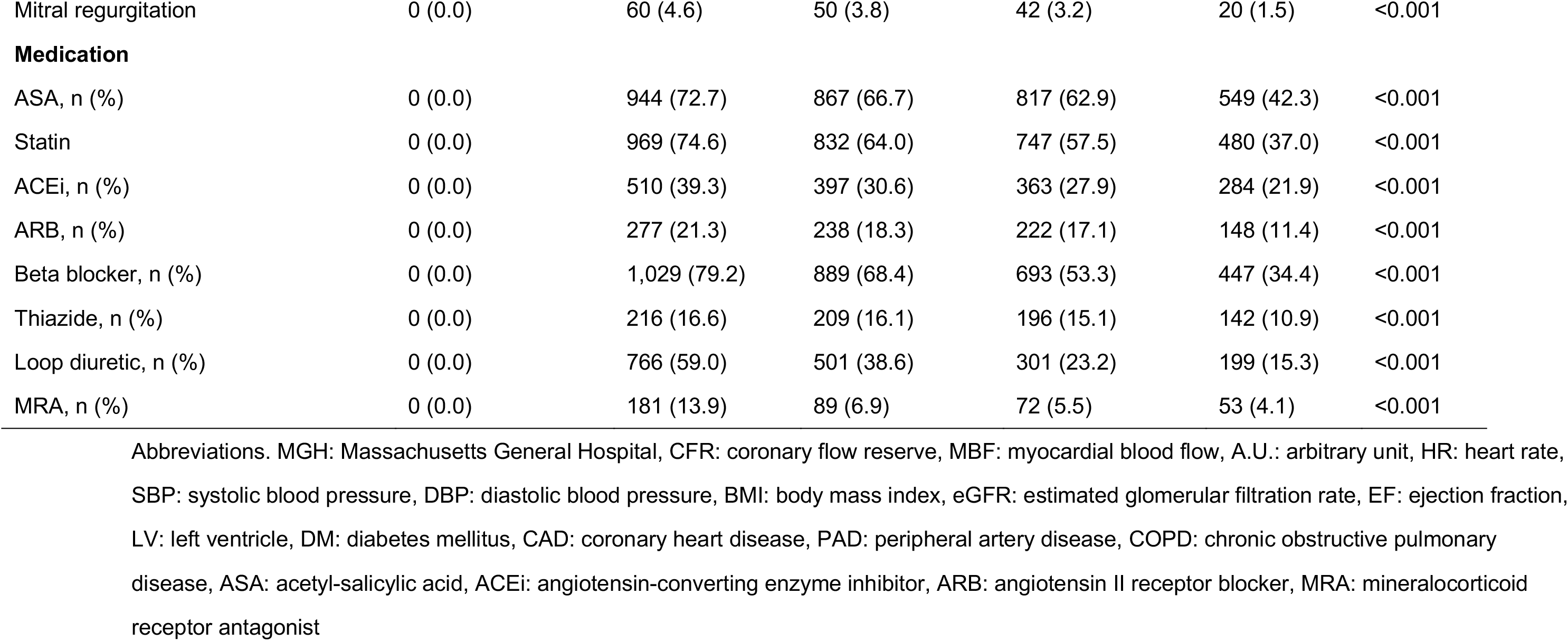
Baseline demographic for EchoAI-stressMBF quartiles for ACS survival analysis (MGH cohort)

|  | Missing n (%) | Quartile 1 | Quartile 2 | Quartile 3 | Quartile 4 | p |
| --- | --- | --- | --- | --- | --- | --- |
| Number patients |  | 1,299 | 1,299 | 1,299 | 1,299 |  |
| **AI model** |  |  |  |  |  |  |
| Echo-AI Rest MBF, A.U. ± SD | 0 (0) | 0.7 ± 0.1 | 0.8 ± 0.1 | 0.9 ± 0.1 | 0.9 ± 0.1 | <0.001 |
| Echo-AI Stress MBF, A.U. ± SD | 0 (0) | 0.6 ± 0.1 | 0.9 ± 0.1 | 1.2 ± 0.1 | 1.6 ± 0.2 | <0.001 |
| Echo-AI CFR, A.U. ± SD | 0 (0) | 1.7 ± 0.2 | 1.8 ± 0.2 | 2.0 ± 0.2 | 2.2 ± 0.3 | <0.001 |
| **Demographics** |  |  |  |  |  |  |
| Age, years ± SD | 0 (0.0) | 71.7 ± 13.5 | 69.5 ± 14.3 | 65.5 ± 15.3 | 57.6 ± 17.0 | <0.001 |
| Gender, Female (%) | 0 (0.0) | 345 (26.6) | 534 (41.1) | 599 (46.1) | 797 (61.4) | <0.001 |
| Smoking | 0 (0) |  |  |  |  | <0.001 |
| Never, n (%) |  | 521 (40.1) | 570 (43.9) | 600 (46.2) | 657 (50.6) |  |
| Current, n (%) |  | 85 (6.5) | 97 (7.5) | 116 (8.9) | 138 (10.6) |  |
| Passive, n (%) |  | 0 (0.0) | 1 (0.1) | 0 (0.0) | 0 (0.0) |  |
| Quit, n (%) |  | 555 (42.7) | 505 (38.9) | 462 (35.6) | 396 (30.5) |  |
| Unknown, n (%) |  | 138 (10.6) | 126 (9.7) | 121 (9.3) | 108 (8.3) |  |
| Race | 0 (0) |  |  |  |  | 0.026 |
| White, n (%) |  | 1,132 (87.1) | 1,127 (86.8) | 1,088 (83.8) | 1,070 (82.4) |  |
| Black, n (%) |  | 68 (5.2) | 57 (4.4) | 74 (5.7) | 78 (6.0) |  |
| Hispanic, n (%) |  | 8 (0.6) | 8 (0.6) | 8 (0.6) | 12 (0.9) |  |
| Asian, n (%) |  | 22 (1.7) | 32 (2.5) | 37 (2.8) | 50 (3.8) |  |
| Others, n (%) |  | 46 (3.5) | 42 (3.2) | 63 (4.8) | 62 (4.8) |  |
| Unknown, n (%) |  | 23 (1.8) | 33 (2.5) | 29 (2.2) | 27 (2.1) |  |
| HR, bpm ± SD | 31 (0.6) | 72.9 ± 11.9 | 72.4 ± 11.7 | 73.4 ± 11.5 | 78.3 ± 11.3 | <0.001 |
| SBP, mmHg ± SD | 27 (0.5) | 126.9 ± 15.5 | 128.4 ± 14.2 | 127.6 ± 13.7 | 125.8 ± 13.9 | <0.001 |
| DBP, mmHg ± SD | 27 (0.5) | 69.0 ± 8.1 | 70.0 ± 7.7 | 71.0 ± 8.0 | 71.3 ± 8.3 | <0.001 |
| BMI, kg/m<sup>2</sup> ± SD | 80 (1.5) | 29.8 ± 8.3 | 29.3 ± 7.3 | 28.6 ± 6.4 | 27.2 ± 7.1 | <0.001 |
| eGFR, ml/min/1.73 m<sup>2</sup> ± SD | 374 (7.2) | 56.5 ± 25.7 | 65.0 ± 26.0 | 73.2 ± 28.6 | 85.5 ± 29.9 | <0.001 |
| EF, % ± SD | 415 (8.0) | 53.4 ± 17.0 | 63.1 ± 11.6 | 66.1 ± 8.8 | 67.5 ± 7.0 | <0.001 |
| **Disease history** |  |  |  |  |  |  |
| Heart failure, n (%) | 0 (0.0) | 816 (62.8) | 512 (39.4) | 301 (23.2) | 121 (9.3) | <0.001 |
| Type 2 DM, n (%) | 0 (0.0) | 487 (37.5) | 338 (26.0) | 289 (22.2) | 208 (16.0) | <0.001 |
| Hypertension, n (%) | 0 (0.0) | 1,121 (86.3) | 1,040 (80.1) | 961 (74.0) | 767 (59.0) | <0.001 |
| Atrial fibrillation, n (%) | 0 (0.0) | 793 (61.0) | 580 (44.6) | 346 (26.6) | 154 (11.9) | <0.001 |
| Hyperlipidemia, n (%) | 0 (0.0) | 946 (72.8) | 884 (68.1) | 812 (62.5) | 594 (45.7) | <0.001 |
| CAD, n (%) | 0 (0.0) | 790 (60.8) | 548 (42.2) | 424 (32.6) | 193 (14.9) | <0.001 |
| Stroke, n (%) | 0 (0.0) | 193 (14.9) | 146 (11.2) | 136 (10.5) | 106 (8.2) | <0.001 |
| PAD, n (%) | 0 (0.0) | 331 (25.5) | 323 (24.9) | 231 (17.8) | 185 (14.2) | <0.001 |
| COPD, n (%) | 0 (0.0) | 202 (15.6) | 172 (13.2) | 157 (12.1) | 145 (11.2) | 0.006 |
| Aortic stenosis | 0 (0.0) | 172 (13.2) | 158 (12.2) | 139 (10.7) | 66 (5.1) | <0.001 |
| Aortic regurgitation | 0 (0.0) | 47 (3.6) | 49 (3.8) | 36 (2.8) | 23 (1.8) | 0.010 |
| Mitral stenosis | 0 (0.0) | 16 (1.2) | 18 (1.4) | 10 (0.8) | 9 (0.7) | 0.214 |
| Mitral regurgitation | 0 (0.0) | 60 (4.6) | 50 (3.8) | 42 (3.2) | 20 (1.5) | <0.001 |
| **Medication** |  |  |  |  |  |  |
| ASA, n (%) | 0 (0.0) | 944 (72.7) | 867 (66.7) | 817 (62.9) | 549 (42.3) | <0.001 |
| Statin | 0 (0.0) | 969 (74.6) | 832 (64.0) | 747 (57.5) | 480 (37.0) | <0.001 |
| ACEi, n (%) | 0 (0.0) | 510 (39.3) | 397 (30.6) | 363 (27.9) | 284 (21.9) | <0.001 |
| ARB, n (%) | 0 (0.0) | 277 (21.3) | 238 (18.3) | 222 (17.1) | 148 (11.4) | <0.001 |
| Beta blocker, n (%) | 0 (0.0) | 1,029 (79.2) | 889 (68.4) | 693 (53.3) | 447 (34.4) | <0.001 |
| Thiazide, n (%) | 0 (0.0) | 216 (16.6) | 209 (16.1) | 196 (15.1) | 142 (10.9) | <0.001 |
| Loop diuretic, n (%) | 0 (0.0) | 766 (59.0) | 501 (38.6) | 301 (23.2) | 199 (15.3) | <0.001 |
| MRA, n (%) | 0 (0.0) | 181 (13.9) | 89 (6.9) | 72 (5.5) | 53 (4.1) | <0.001 |

Abbreviations. MGH: Massachusetts General Hospital, CFR: coronary flow reserve, MBF: myocardial blood flow, A.U.: arbitrary unit, HR: heart rate, SBP: systolic blood pressure, DBP: diastolic blood pressure, BMI: body mass index, eGFR: estimated glomerular filtration rate, EF: ejection fraction, LV: left ventricle, DM: diabetes mellitus, CAD: coronary heart disease, PAD: peripheral artery disease, COPD: chronic obstructive pulmonary disease, ASA: acetyl-salicylic acid, ACEi: angiotensin-converting enzyme inhibitor, ARB: angiotensin II receptor blocker, MRA: mineralocorticoid receptor antagonist

**Table S15.** Baseline demographic for EchoAI-CFR quartiles for ACS survival analysis (MGH cohort)

|  | Missing n (%) | Quartile 1 | Quartile 2 | Quartile 3 | Quartile 4 | p |
| --- | --- | --- | --- | --- | --- | --- |
| Number patients |  | 1,299 | 1,299 | 1,299 | 1,299 |  |
| **AI model** |  |  |  |  |  |  |
| Echo-AI Rest MBF, A.U. ± SD | 0 (0) | 0.8 ± 0.1 | 0.8 ± 0.1 | 0.8 ± 0.1 | 0.8 ± 0.1 | <0.001 |
| Echo-AI Stress MBF, A.U. ± SD | 0 (0) | 0.8 ± 0.3 | 1.0 ± 0.3 | 1.1 ± 0.3 | 1.4 ± 0.3 | <0.001 |
| Echo-AI CFR, A.U. ± SD | 0 (0) | 1.6 ± 0.1 | 1.8 ± 0.0 | 2.0 ± 0.1 | 2.3 ± 0.2 | <0.001 |
| **Demographics** |  |  |  |  |  |  |
| Age, years ± SD | 0 (0.0) | 72.8 ± 13.1 | 70.1 ± 13.9 | 65.9 ± 14.6 | 55.6 ± 16.7 | <0.001 |
| Gender, Female (%) | 0 (0.0) | 519 (40.0) | 518 (39.9) | 591 (45.5) | 647 (49.8) | <0.001 |
| Smoking | 0 (0) |  |  |  |  | <0.001 |
| Never, n (%) |  | 542 (41.7) | 553 (42.6) | 585 (45.0) | 668 (51.4) |  |
| Current, n (%) |  | 86 (6.6) | 87 (6.7) | 127 (9.8) | 136 (10.5) |  |
| Passive, n (%) |  | 0 (0.0) | 1 (0.1) | 0 (0.0) | 0 (0.0) |  |
| Quit, n (%) |  | 550 (42.3) | 525 (40.4) | 452 (34.8) | 391 (30.1) |  |
| Unknown, n (%) |  | 121 (9.3) | 133 (10.2) | 135 (10.4) | 104 (8.0) |  |
| Race | 0 (0) |  |  |  |  | 0.240 |
| White, n (%) |  | 1,132 (87.1) | 1,111 (85.5) | 1,095 (84.3) | 1,079 (83.1) |  |
| Black, n (%) |  | 59 (4.5) | 72 (5.5) | 68 (5.2) | 78 (6.0) |  |
| Hispanic, n (%) |  | 4 (0.3) | 12 (0.9) | 11 (0.8) | 9 (0.7) |  |
| Asian, n (%) |  | 36 (2.8) | 32 (2.5) | 37 (2.8) | 36 (2.8) |  |
| Others, n (%) |  | 40 (3.1) | 46 (3.5) | 58 (4.5) | 69 (5.3) |  |
| Unknown, n (%) |  | 28 (2.2) | 26 (2.0) | 30 (2.3) | 28 (2.2) |  |
| HR, bpm ± SD | 31 (0.6) | 73.2 ± 10.6 | 74.0 ± 12.0 | 75.0 ± 12.1 | 74.7 ± 12.4 | 0.001 |
| SBP, mmHg ± SD | 27 (0.5) | 128.4 ± 15.0 | 128.6 ± 14.5 | 127.9 ± 14.4 | 123.9 ± 13.1 | <0.001 |
| DBP, mmHg ± SD | 27 (0.5) | 68.7 ± 7.9 | 70.1 ± 7.9 | 70.9 ± 8.0 | 71.7 ± 8.2 | <0.001 |
| BMI, kg/m<sup>2</sup> ± SD | 80 (1.5) | 29.7 ± 7.7 | 29.1 ± 6.5 | 28.9 ± 7.9 | 27.2 ± 7.1 | <0.001 |
| eGFR, ml/min/1.73 m<sup>2</sup> ± SD | 374 (7.2) | 58.6 ± 27.4 | 64.3 ± 27.3 | 72.2 ± 27.8 | 85.4 ± 28.9 | <0.001 |
| EF, % ± SD | 415 (8.0) | 58.5 ± 16.3 | 61.8 ± 13.5 | 64.3 ± 11.0 | 65.7 ± 8.2 | <0.001 |
| **Disease history** |  |  |  |  |  |  |
| Heart failure, n (%) | 0 (0.0) | 683 (52.6) | 549 (42.3) | 369 (28.4) | 149 (11.5) | <0.001 |
| Type 2 DM, n (%) | 0 (0.0) | 470 (36.2) | 354 (27.3) | 319 (24.6) | 179 (13.8) | <0.001 |
| Hypertension, n (%) | 0 (0.0) | 1,104 (85.0) | 1,064 (81.9) | 1,000 (77.0) | 721 (55.5) | <0.001 |
| Atrial fibrillation, n (%) | 0 (0.0) | 651 (50.1) | 573 (44.1) | 437 (33.6) | 212 (16.3) | <0.001 |
| Hyperlipidemia, n (%) | 0 (0.0) | 940 (72.4) | 883 (68.0) | 797 (61.4) | 616 (47.4) | <0.001 |
| CAD, n (%) | 0 (0.0) | 719 (55.4) | 570 (43.9) | 403 (31.0) | 263 (20.2) | <0.001 |
| Stroke, n (%) | 0 (0.0) | 200 (15.4) | 132 (10.2) | 155 (11.9) | 94 (7.2) | <0.001 |
| PAD, n (%) | 0 (0.0) | 321 (24.7) | 282 (21.7) | 268 (20.6) | 199 (15.3) | <0.001 |
| COPD, n (%) | 0 (0.0) | 216 (16.6) | 184 (14.2) | 174 (13.4) | 102 (7.9) | <0.001 |
| Aortic stenosis | 0 (0.0) | 202 (15.6) | 144 (11.1) | 123 (9.5) | 66 (5.1) | <0.001 |
| Aortic regurgitation | 0 (0.0) | 41 (3.2) | 39 (3.0) | 46 (3.5) | 29 (2.2) | 0.255 |
| Mitral stenosis | 0 (0.0) | 18 (1.4) | 18 (1.4) | 10 (0.8) | 7 (0.5) | 0.065 |
| Mitral regurgitation | 0 (0.0) | 53 (4.1) | 49 (3.8) | 38 (2.9) | 32 (2.5) | 0.079 |
| **Medication** |  |  |  |  |  |  |
| ASA, n (%) | 0 (0.0) | 948 (73.0) | 838 (64.5) | 801 (61.7) | 590 (45.4) | <0.001 |
| Statin | 0 (0.0) | 946 (72.8) | 851 (65.5) | 765 (58.9) | 466 (35.9) | <0.001 |
| ACEi, n (%) | 0 (0.0) | 463 (35.6) | 441 (33.9) | 376 (28.9) | 274 (21.1) | <0.001 |
| ARB, n (%) | 0 (0.0) | 269 (20.7) | 246 (18.9) | 219 (16.9) | 151 (11.6) | <0.001 |
| Beta blocker, n (%) | 0 (0.0) | 968 (74.5) | 875 (67.4) | 743 (57.2) | 472 (36.3) | <0.001 |
| Thiazide, n (%) | 0 (0.0) | 219 (16.9) | 229 (17.6) | 184 (14.2) | 131 (10.1) | <0.001 |
| Loop diuretic, n (%) | 0 (0.0) | 683 (52.6) | 521 (40.1) | 391 (30.1) | 172 (13.2) | <0.001 |
| MRA, n (%) | 0 (0.0) | 155 (11.9) | 105 (8.1) | 95 (7.3) | 40 (3.1) | <0.001 |

Abbreviations. MGH: Massachusetts General Hospital, CFR: coronary flow reserve, MBF: myocardial blood flow, A.U.: arbitrary unit, HR: heart rate, SBP: systolic blood pressure, DBP: diastolic blood pressure, BMI: body mass index, eGFR: estimated glomerular filtration rate, EF: ejection fraction, LV: left ventricle, DM: diabetes mellitus, CAD: coronary heart disease, PAD: peripheral artery disease, COPD: chronic obstructive pulmonary disease, ASA: acetyl-salicylic acid, ACEi: angiotensin-converting enzyme inhibitor, ARB: angiotensin II receptor blocker, MRA: mineralocorticoid receptor antagonist

